# HORMONAL THERAPIES FOR ENDOMETRIOSIS: A SYSTEMATIC REVIEW AND META-ANALYSIS OF RANDOMISED HEAD-TO-HEAD TRIALS

**DOI:** 10.64898/2026.08.26.26361442

**Authors:** Veronica Bandini, Lucy HR Whitaker, Katy Vincent, Noemi Salmeri, Rebecca Mawson, Paolo Vercellini, Andrew W Horne

**Author notes:** Corresponding author: Professor Andrew Horne, Centre for Reproductive Health, Institute for Regeneration and Repair, University of Edinburgh, Edinburgh, United Kingdom.

## Abstract

**Background:** Endometriosis is a chronic pain condition in which hormonal therapies form the cornerstone of long-term management. Treatment tolerability is critical for adherence and therapeutic success, but most comparative studies and reviews have focused on their ability to reduce menstrual pain, while their impact on non-menstrual pelvic pain (NMPP), bleeding patterns, adverse events (AEs), treatment discontinuation and quality of life (QoL) remain poorly characterised. This systematic review and meta-analysis evaluate these outcomes across currently available hormonal therapies, providing practical evidence for clinical decision-making.

**Methods:** PubMed/MEDLINE, Scopus, and Embase were searched up to November 2025 for randomised controlled trials comparing at least two active first- or second-line hormonal treatments for endometriosis. Studies without confirmed endometriosis, treatment duration less than three months and comparing therapies to placebo only were excluded. Data were extracted by two reviewers from reports. Pain outcomes were pooled as mean differences (MD, 95% CI), with bleeding patterns, AEs, and discontinuations as proportions. Analyses were performed in R. PROSPERO: CRD420251137785.

**Findings:** Of 1892 records screened, 48 trials (5583 women) met our inclusion criteria. Overall pelvic pain (0-10 scale) was significantly reduced across all treatment categories (*p*<0·001): combined oral contraceptives (COCs) (MD 3·17), oral and long-acting progestogens (MD 3·83; MD 4·29), and GnRH-analogues (MD 3·81). Sensitivity analyses restricted to studies reporting NMPP yielded comparable results. GnRH-agonists showed the most favourable bleeding profile, followed by continuous COCs. However, all regimens reported class-specific AEs, including mood changes, nausea, headache, weight gain, and decreased libido (pooled proportions >10%). Overall discontinuation due to AEs was 7·7%, and vaginal bleeding was the leading cause. Heterogeneity across meta-analyses was high. Risk of bias (RoB2) was moderate to high.

**Interpretation:** Given similar reductions in overall pelvic pain across hormonal therapies, treatment decisions should prioritise differences in bleeding profiles, therapy-specific AEs, and QoL.

**Funding:** None.

**Panel: Research in context:** *Evidence before this study:* Hormonal therapies are widely recommended as first-line treatment for endometriosis-associated pain and several randomised controlled trials (RCTs), systematic reviews, and network meta-analyses have demonstrated their effectiveness. However, the available evidence is limited by small study populations, few direct head-to-head comparisons, heterogeneous outcome measures, and short follow-up. Most original studies and reviews focus primarily on pain outcomes (particularly menstrual pain or “dysmenorrhea”), whereas overall pelvic pain and patient-centred outcomes, such as quality of life, treatment satisfaction, acceptability, and tolerability are infrequently assessed, leaving uncertainty about the comparative safety and tolerability profiles of commonly used medical treatments. We searched PubMed/MEDLINE, Scopus, and Embase up to November 2025 using terms related to endometriosis, adenomyosis, and hormonal treatments. We included 48 RCTs comparing two or more first- or second-line hormonal therapies. Exclusion criteria were non-English articles, endometriosis not confirmed by imaging or surgery, placebo-controlled trials, therapy length less than three months.

*Added value of this study:* This systematic review and meta-analysis provide a comprehensive comparison of the most commonly used hormonal therapies for endometriosis. Unlike most previous syntheses, we evaluated overall pelvic pain and non-menstrual pelvic pain (NMPP) as primary pain outcomes (not only menstrual pain or ‘dysmenorrhea’), as a more clinically relevant measure of endometriosis-associated pain, and conducted the first systematic comparisons of continuous versus cyclic combined oral contraceptive (COC) regimens. We also evaluated safety, tolerability, bleeding patterns, treatment discontinuation, and other patient-centred outcomes. In addition to confirming the effectiveness of hormonal therapies in reducing overall pelvic pain and NMPP, our findings highlight important differences in bleeding profiles and adverse events across regimens.

*Implications of all the available evidence:* The findings of this review reinforce the need for an individualised, patient-centred approach to hormonal management of endometriosis, weighing efficacy alongside safety, tolerability, bleeding profiles, and patient preferences. To better inform treatment decisions, future research should prioritise longer-term head-to-head comparisons of hormonal therapies and include a range of validated patient-reported outcomes among the endpoints assessed.

## INTRODUCTION

Endometriosis affects an estimated 190 million women and those assigned female at birth and is a leading cause of chronic non-menstrual pelvic pain (NMPP), dysmenorrhoea, deep dyspareunia, and impaired quality of life (QoL).^1^ In those not actively seeking pregnancy, hormonal therapy represents the cornerstone of long-term management, aiming to suppress disease activity and control symptoms while avoiding repeated surgery.^2–5^ Over the past decades, the hormonal therapeutic armamentarium has expanded substantially, encompassing newer combined oral contraceptive (COCs) formulations, oral and long-acting progestogens, GnRH-agonists, and, more recently, oral GnRH-antagonists.^6,7^

Several systematic reviews, meta-analyses, and network meta-analyses have evaluated hormonal therapies for endometriosis, consistently confirming their efficacy in reducing dysmenorrhea and preventing disease progression.^8–15^ The key unmet clinical question is therefore no longer to demonstrate that hormonal therapy is effective in reducing dysmenorrhoea, but to determine which treatment should be preferred for each individual patient. Because medical treatment is often continued for years, if not decades, its long-term success depends not only on efficacy, both menstrual and non-menstrual, but also on safety and tolerability.

We therefore conducted a systematic review and meta-analysis of randomised clinical trial (RCTs) directly comparing active hormonal treatments and focusing on currently recommended therapeutic options for endometriosis. The present study aimed to providing clinically relevant data to facilitate treatment decisions across a wide range of outcomes. These outcomes included overall pelvic pain and NMPP, bleeding patterns, adverse events (AEs), treatment discontinuation, QoL and treatment satisfaction.

## METHODS

### Search strategy and selection criteria

This systematic review and meta-analysis were conducted according to a predefined protocol and were prospectively registered on PROSPERO (CRD420251137785).

Electronic searches were performed from inception to November 2025 in PubMed/MEDLINE, Embase, and Scopus using a combination of MeSH terms and free-text keywords related to endometriosis, adenomyosis, and hormonal treatments. The complete search strategy is provided in appendix (p.3). Reference lists of included studies were also screened to identify additional eligible articles. Grey literature sources and unpublished studies were not systematically searched. Only aggregate published data were extracted.

Eligible studies were RCTs comparing two or more first- or second-line hormonal therapies for endometriosis. Included treatments comprised COCs, dienogest (DNG), norethisterone acetate (NETA), drospirenone (DRSP), desogestrel (DSG), oral and depot medroxyprogesterone acetate (MPA and DMPA), levonorgestrel-releasing intrauterine device (LNG-IUD), etonogestrel implant (ENG), and GnRH-agonists and antagonists. Studies were included only when the hormonal compound and dosage regimen were clearly reported. Exclusion criteria were non-English publications, lack of imaging or surgical confirmation of endometriosis, placebo-controlled trials, and treatment duration shorter than three months.

The primary outcome was the reduction in pain symptoms, including any of the following overall pelvic pain, NMPP, dysmenorrhoea, and deep dyspareunia. Secondary outcomes included bleeding patterns (spotting, breakthrough bleeding, and amenorrhoea), AEs, treatment discontinuation, QoL, recurrence of lesions and/or symptoms, patient satisfaction and change in analgesic use during treatment.

Further details regarding study selection, data extraction, and handling of heterogeneous outcome reporting are provided in the appendix (p.4-6).

### Data analysis

All statistical analyses were conducted using R software (R Foundation for Statistical Computing, Vienna, Austria). Meta-analyses were performed using the “meta” and “metafor” packages.

For continuous pain outcomes measured using comparable 0–10 pain scales, the pooled effect sizes were calculated as mean differences (MDs), along with their corresponding 95% confidence intervals (CIs), using the “metagen” function. Random-effects models were applied throughout to account for expected clinical and methodological heterogeneity across studies. For meta-analysis of pain outcomes, data were synthesised at two levels. First, interventions were grouped into four predefined therapeutic categories (COCs, oral progestogens, long-acting progestogens, and GnRH-analogues). Second, subgroup analyses were performed according to specific regimens whenever at least two studies were available. Bleeding patterns, AEs, and treatment discontinuation were evaluated at the individual regimen level only.

For dichotomous outcomes (bleeding patterns, AEs, and treatment discontinuation) pooled proportions were estimated using the “metaprop” function. Bleeding outcomes were evaluated separately for spotting, breakthrough bleeding, and amenorrhoea, whereas AEs were analysed according to event type. For treatment discontinuation, only those attributable to AEs were included in the quantitative synthesis. Proportions were pooled using random-effects models with logit transformation, and between-study variance was estimated using the restricted maximum likelihood (REML) method.

Between-study heterogeneity was quantified using the *I*² statistic. When substantial heterogeneity was observed, potential sources were explored through subgroup and/or sensitivity analyses. Potential small-study effects in the main meta-analyses were evaluated using Egger’s regression asymmetry test.

Given the substantial heterogeneity in QoL instruments and reporting methods across studies, quantitative meta-analysis was not feasible. Therefore, synthesis without meta-analysis (SWiM) using vote counting based on direction of effect was performed according to Cochrane and SWiM guidance.^16^ When available, summary questionnaire scores were used to determine direction of effect; otherwise, domain-specific scores were considered. Treatment arms were classified according to the reported direction of effect on QoL as positive, no effect, or negative. Harvest plots were constructed to visually summarise the distribution of treatment effects across hormonal treatment classes.

Finally, data on dysmenorrhoea, deep dyspareunia, lesion recurrence or reappearance of symptoms, reasons for treatment discontinuation other than AEs, treatment satisfaction and change in analgesic use were summarised qualitatively, primarily because of substantial heterogeneity in outcome definitions and reporting across studies.

Risk of bias was independently assessed by two reviewers (V.B. and N.S.) using the revised Cochrane risk-of-bias tool for RCTs (RoB 2).^17^ Disagreements were resolved by consensus. Trustworthiness of the included RCTs was assessed according to criteria recently proposed by the Obstetrics and Gynecology Editors’ Integrity Group (OGEIG).^18^ The overall certainty of evidence for the main outcomes was assessed using the Grading of Recommendations Assessment, Development and Evaluation (GRADE) framework.^19^

### Role of the funding source

N/A (no funding)

## RESULTS

A total of 2607 records were identified through database searching. After removal of duplicates and non-English publications, 1892 records underwent title and abstract screening, of which 504 were excluded. Ultimately, of the 1388 full-text articles assessed, 48 RCTs involving 5583 women were included in the qualitative synthesis, with 47 eligibles for meta-analysis (one study was excluded as the only outcome reported was lesions recurrence). The reasons for exclusion and the entire study selection process are detailed in Figure 1.

**Figure 1.**
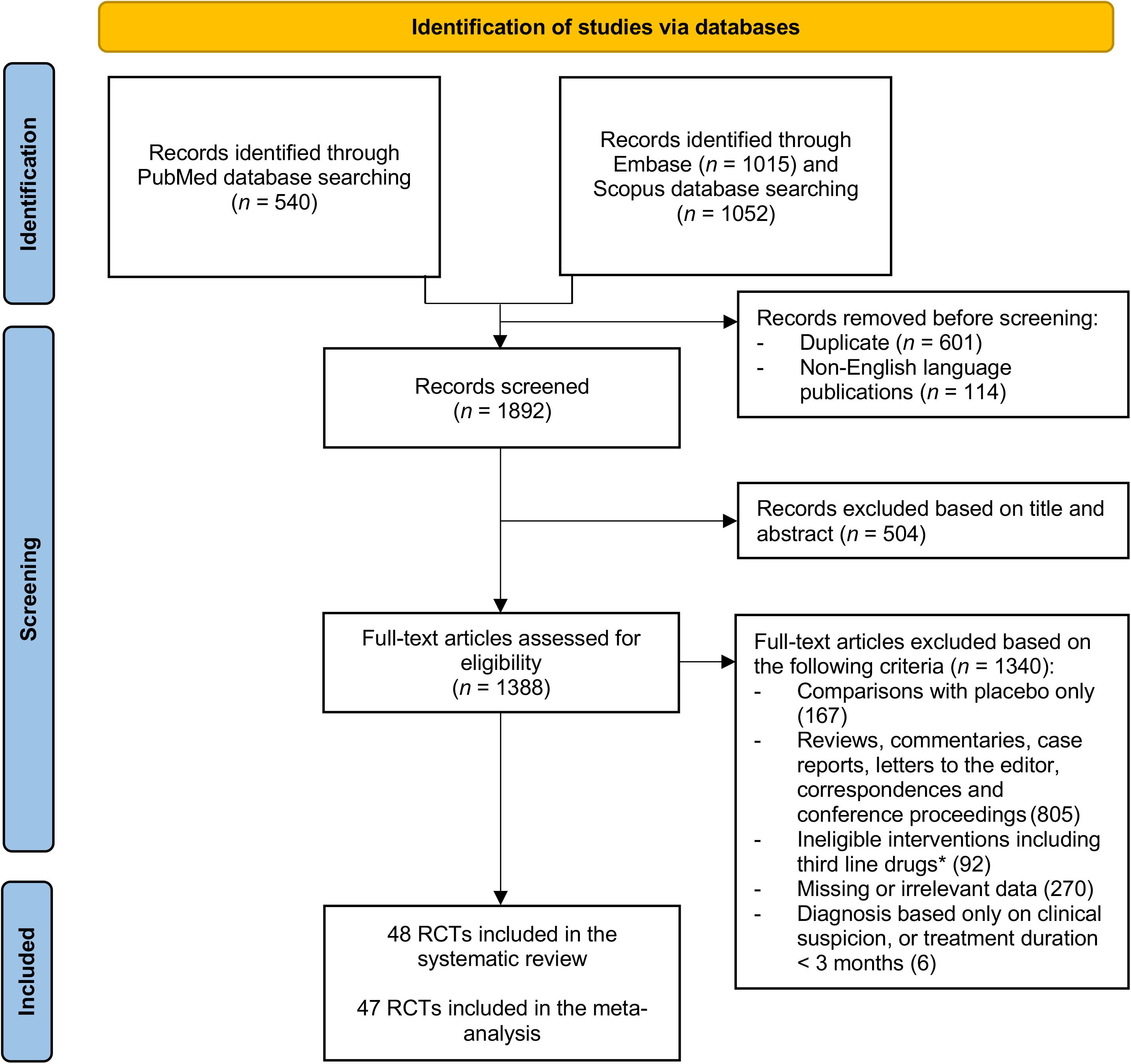
PRISMA flow diagram of study selection. Danazol, aromatase inhibitors (letrozole and anastrozole), gestrinone, and GnRH-agonists administered as nasal sprays (nafarelin and buserelin) were not considered eligible interventions.

The characteristics of the included RCTs are summarised in Tables 1 and 2. The studies were grouped according to the hormonal treatment classes evaluated in the original trial. Studies comparing only COCs and/or oral progestogens were considered separately^20–34^, whereas studies including at least one GnRH-analogue^35–55^ or one long-acting progestogen (LAP)^56–67^ treatment arm were classified within the corresponding category. Table 1 summarises study and patient characteristics, including sample size, demographic features, disease phenotype and stage, presence of adenomyosis, and diagnostic methods. Table 2 provides an overview about treatment characteristics and outcomes, including interventions and comparators, treatment and follow-up duration, outcome measures, and the main findings regarding efficacy and tolerability.

**Table 1.**
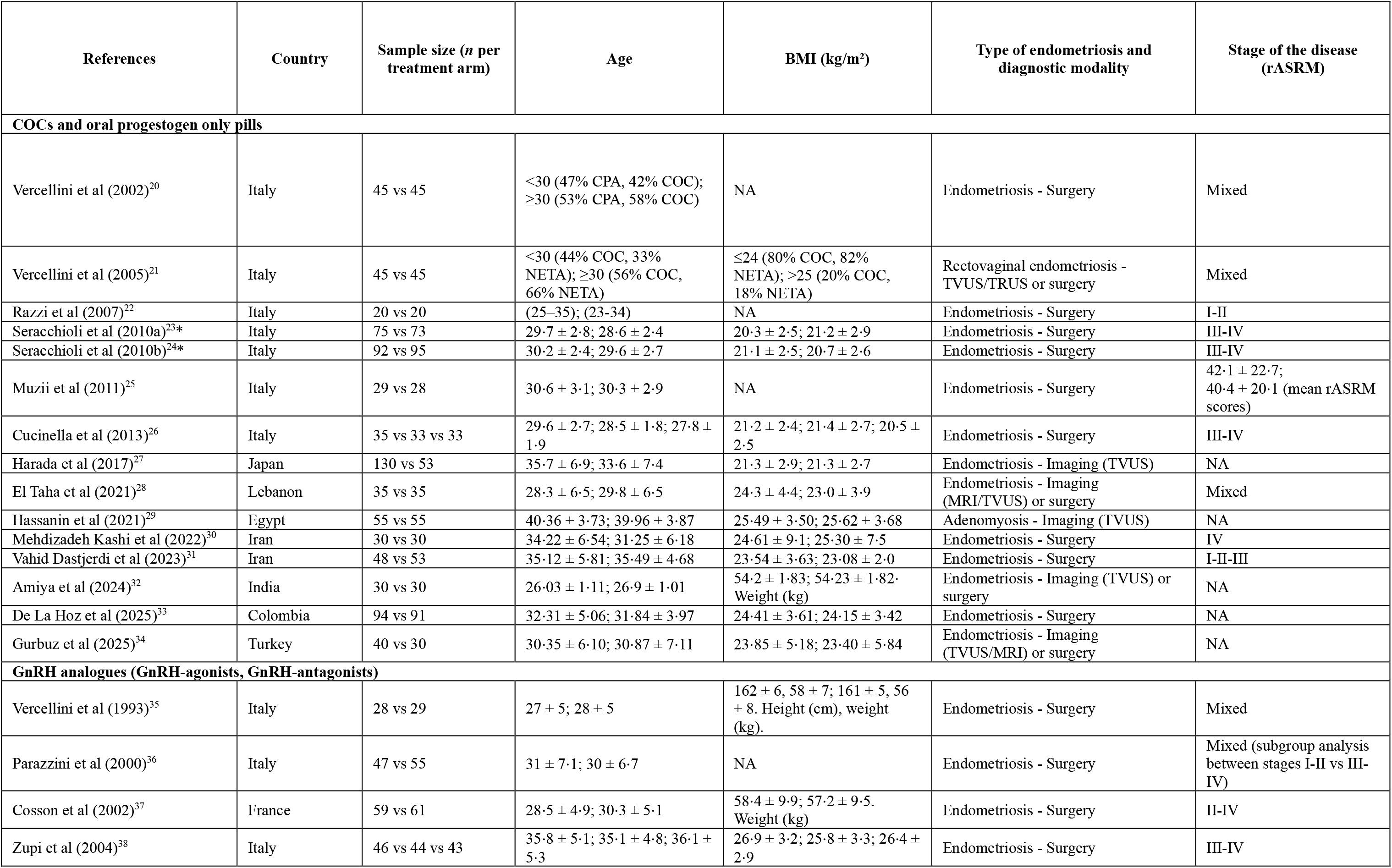
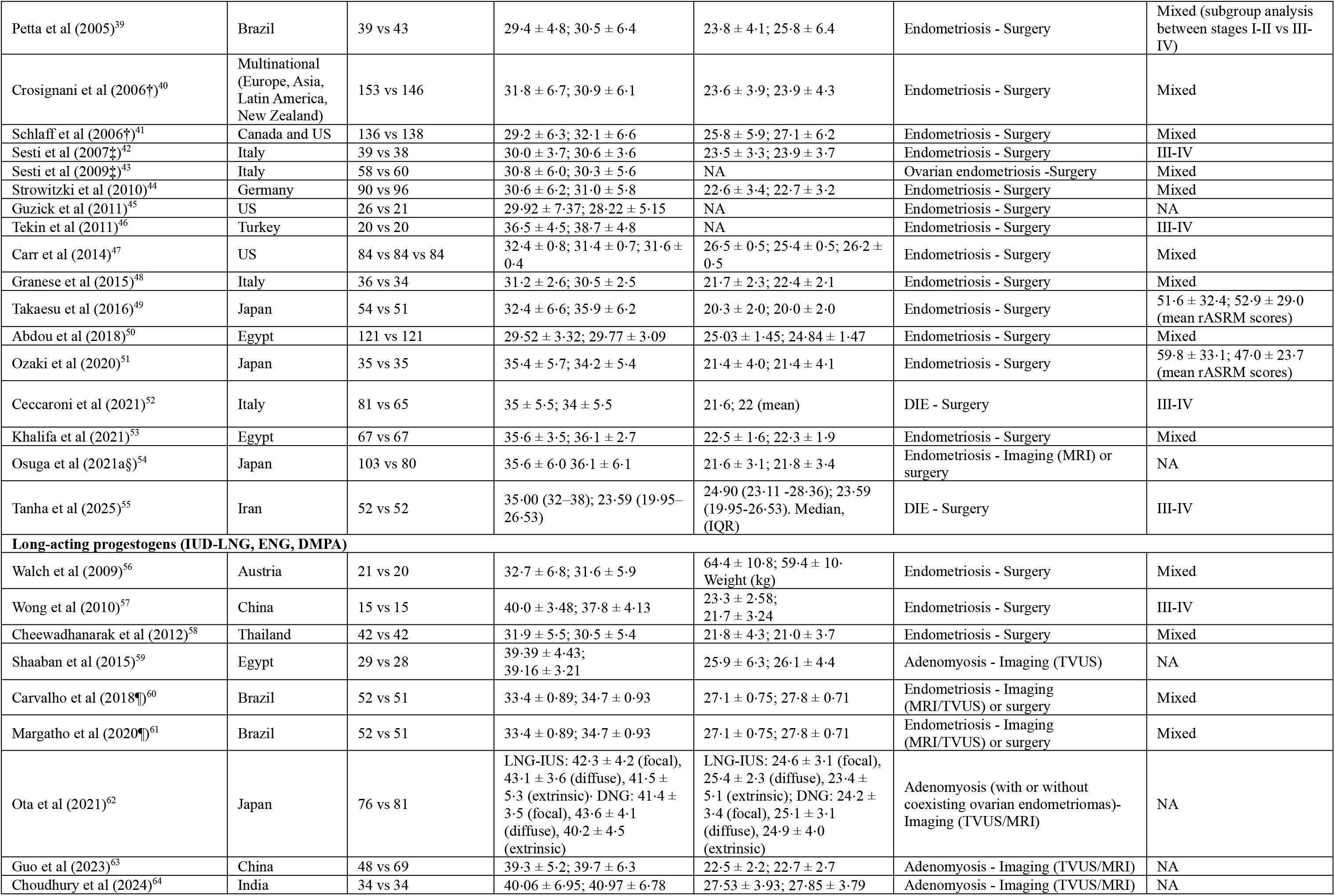
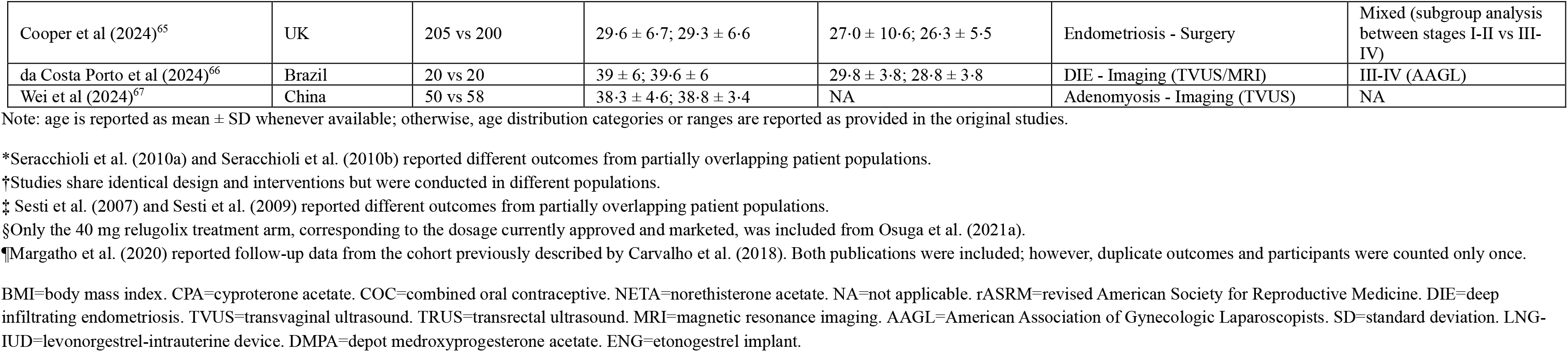
Patient characteristics of RCTs with hormonal therapies and endometriosis.

**Table 2.**
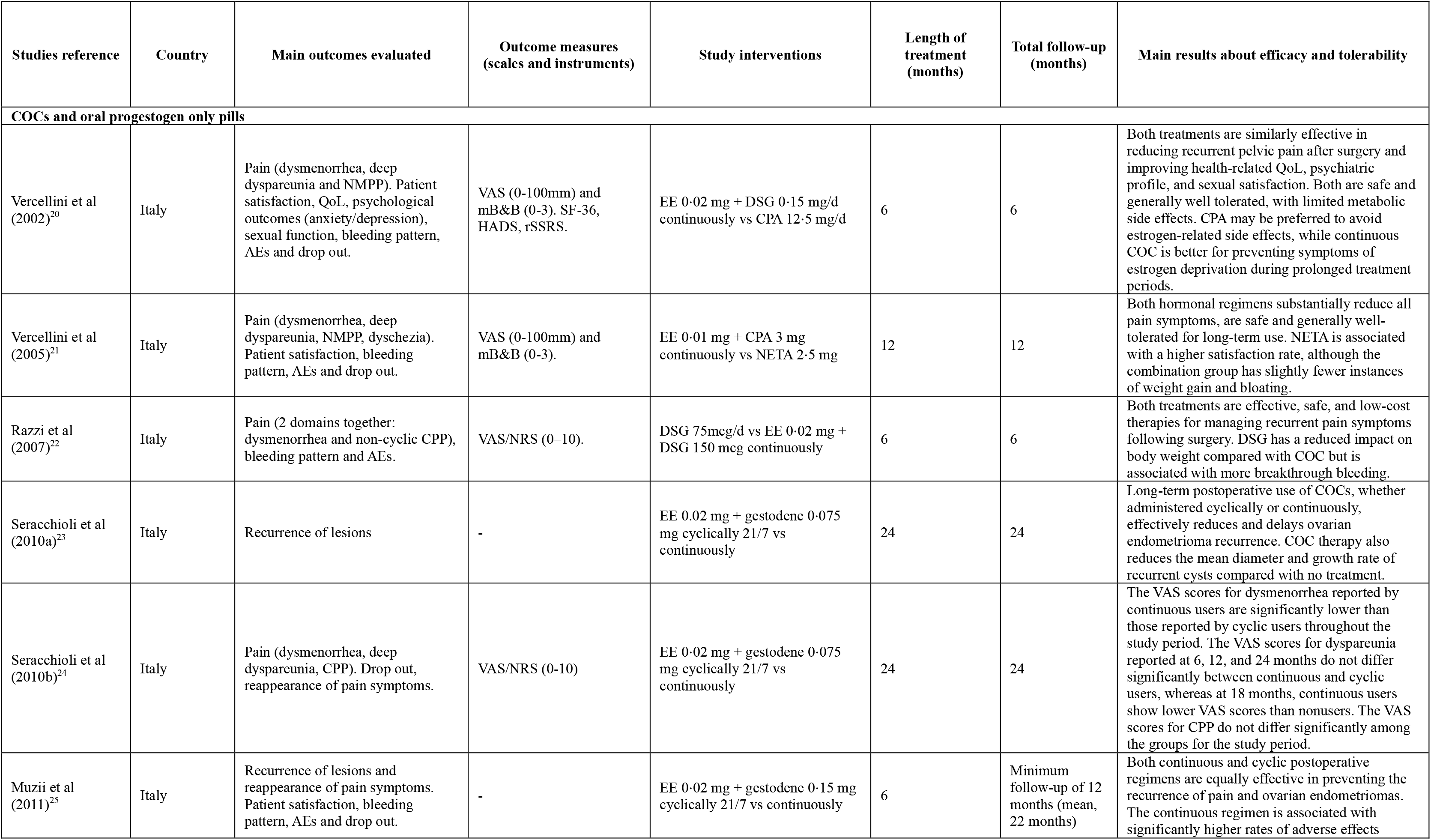
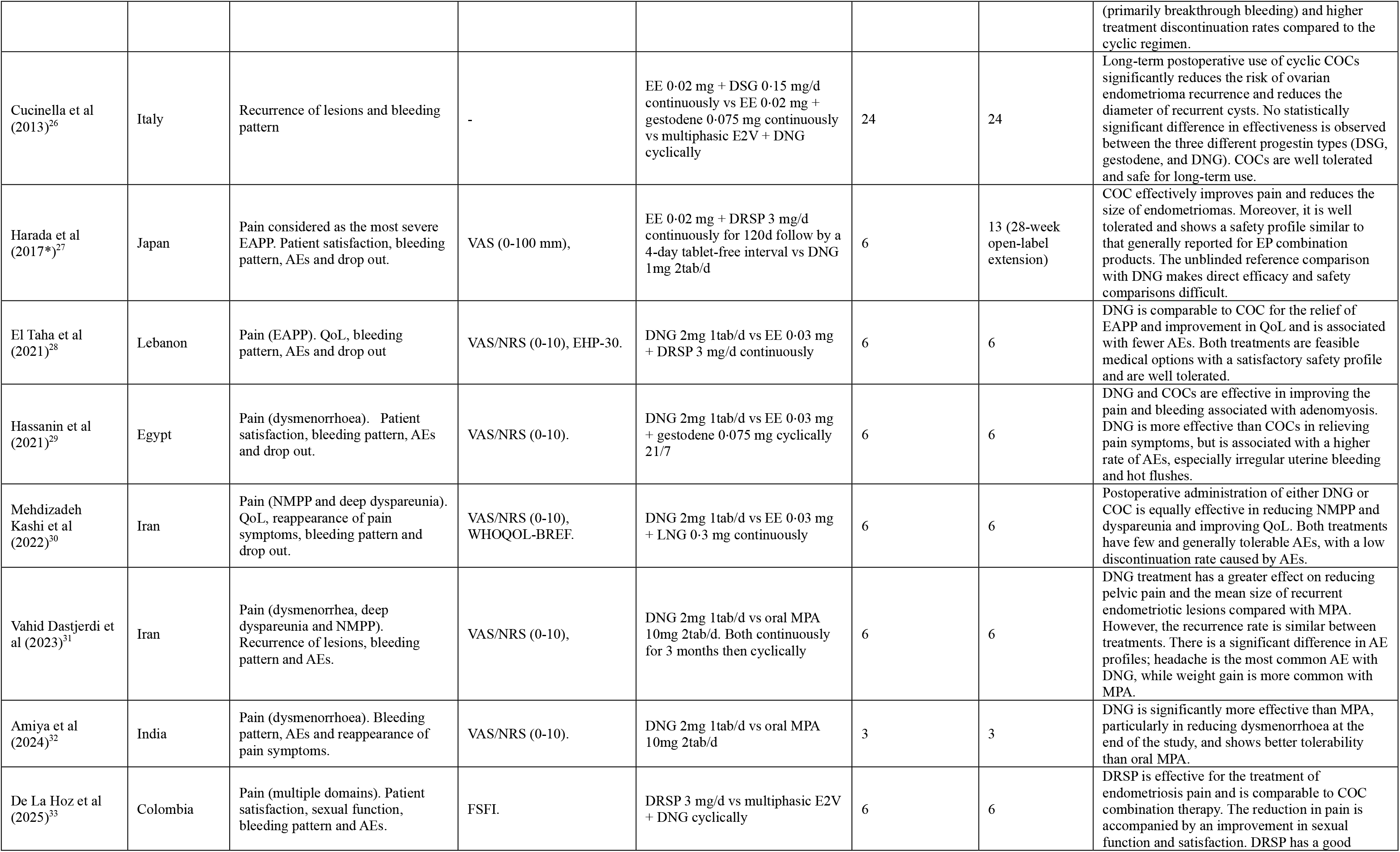
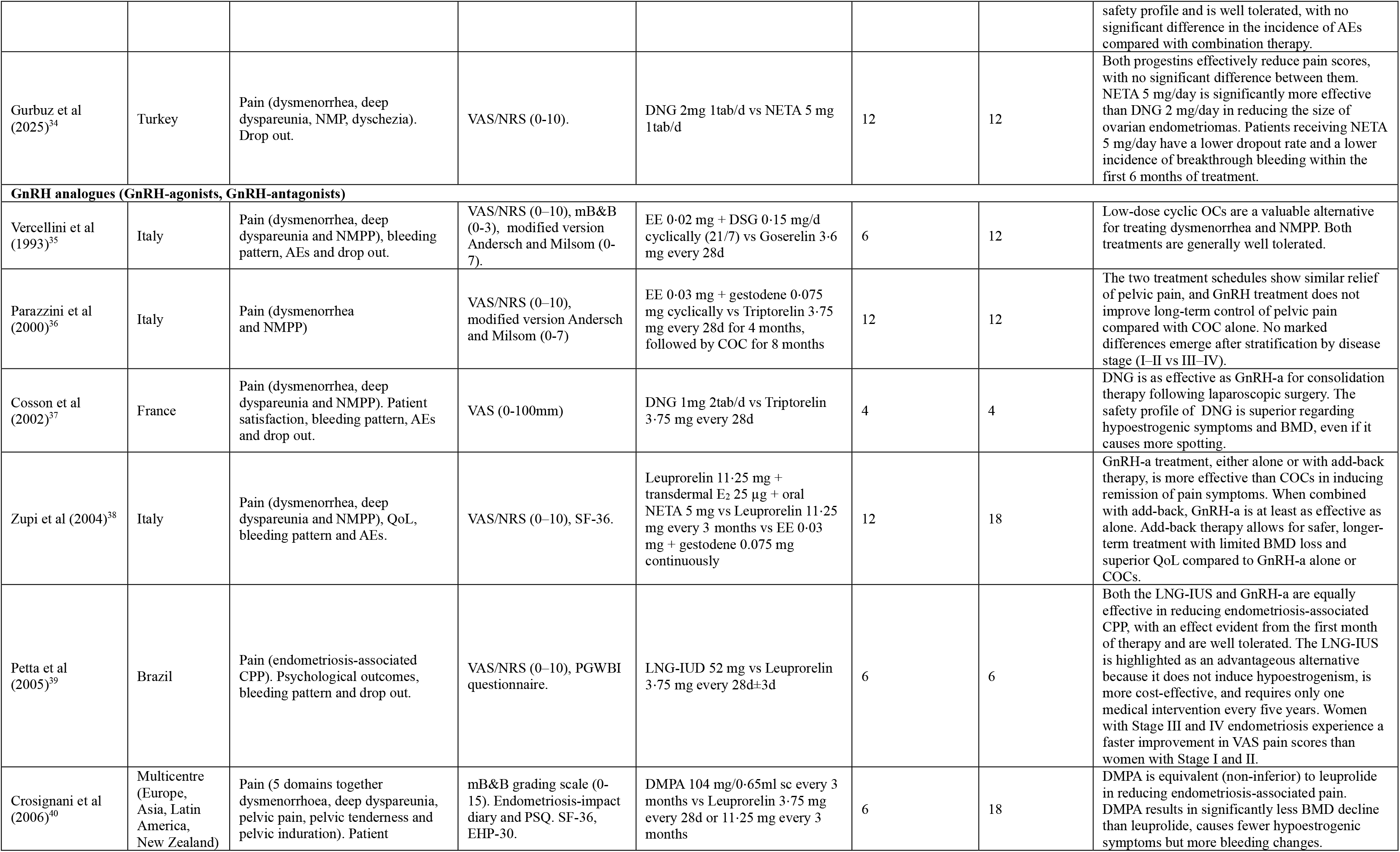
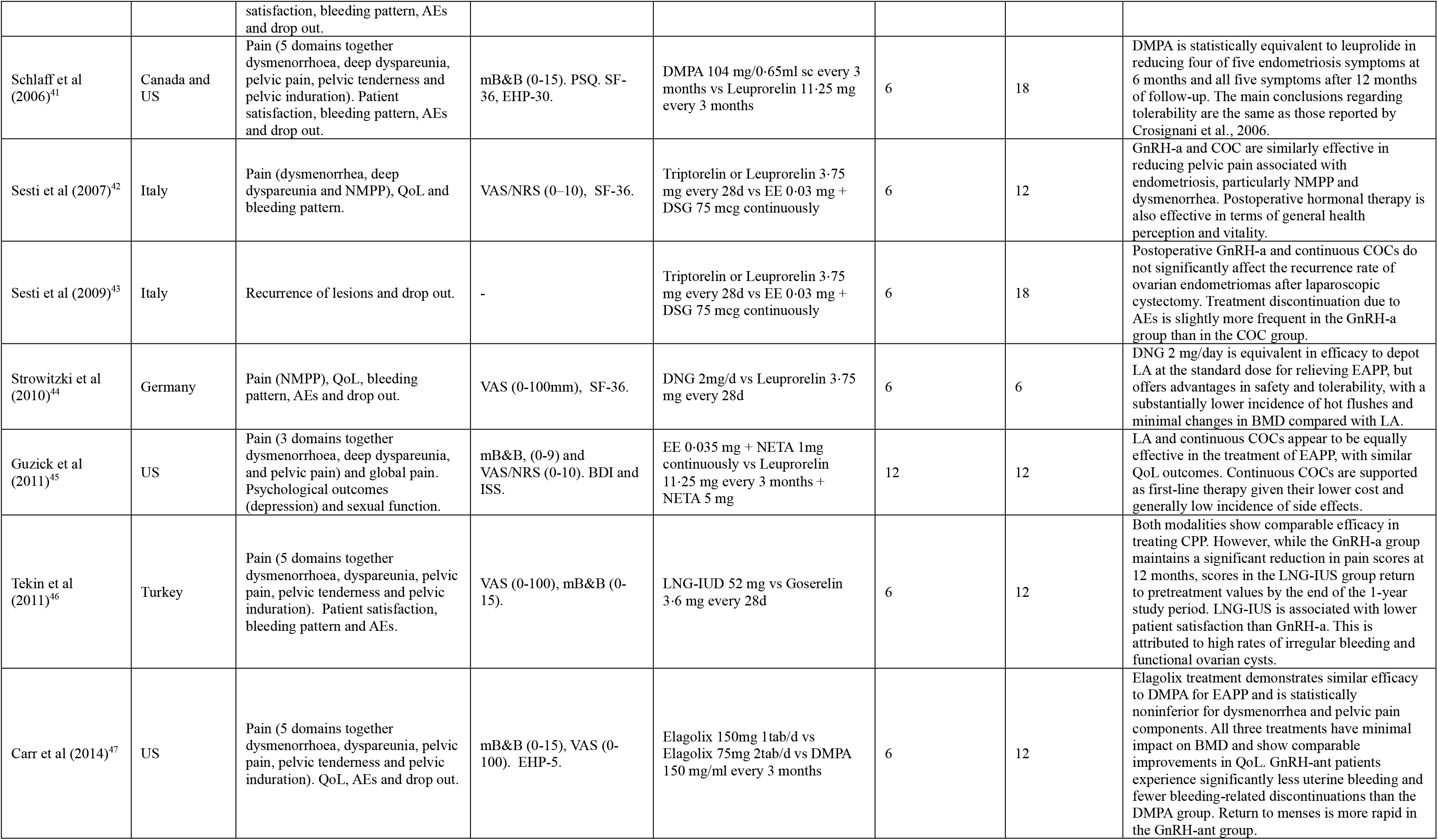
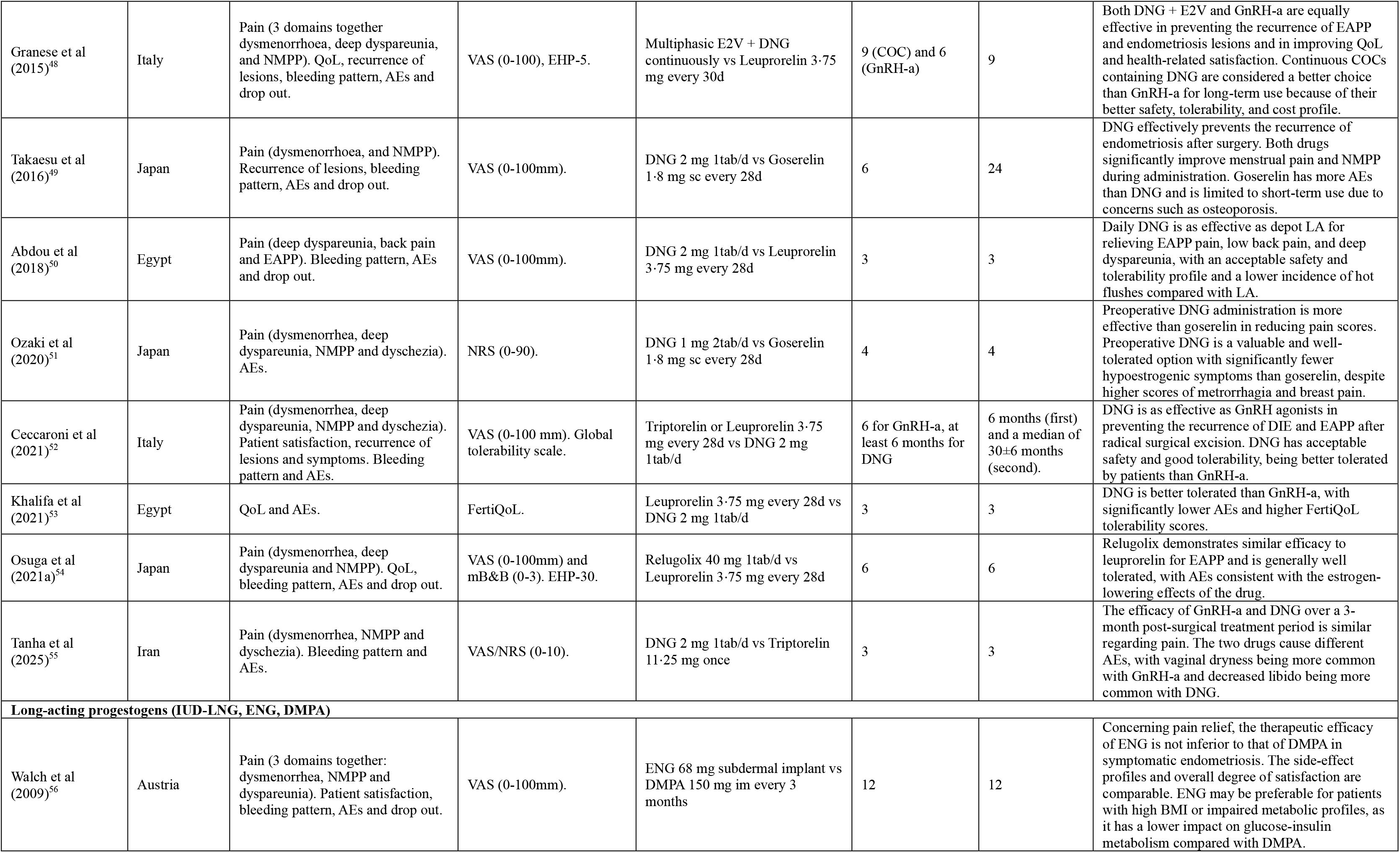
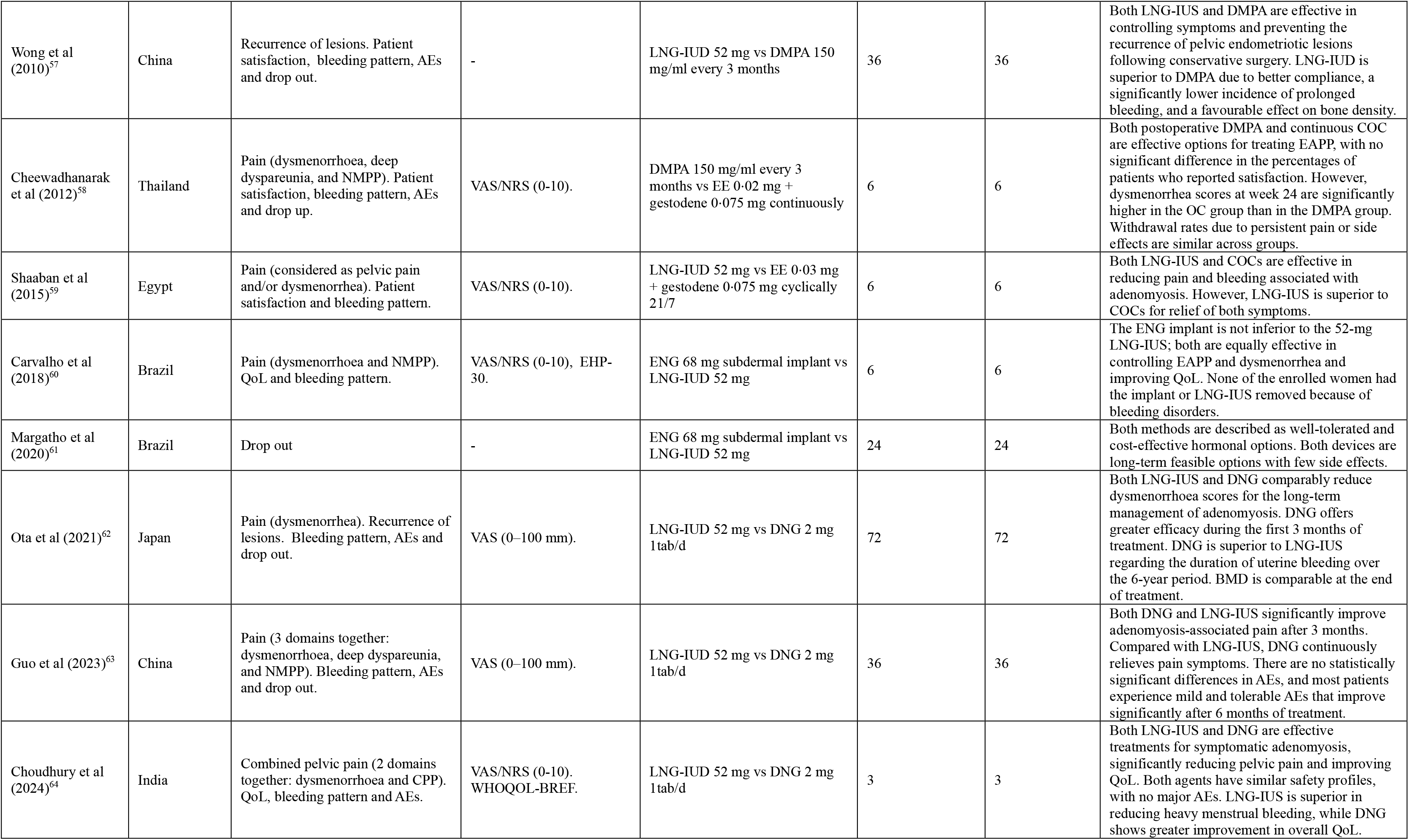
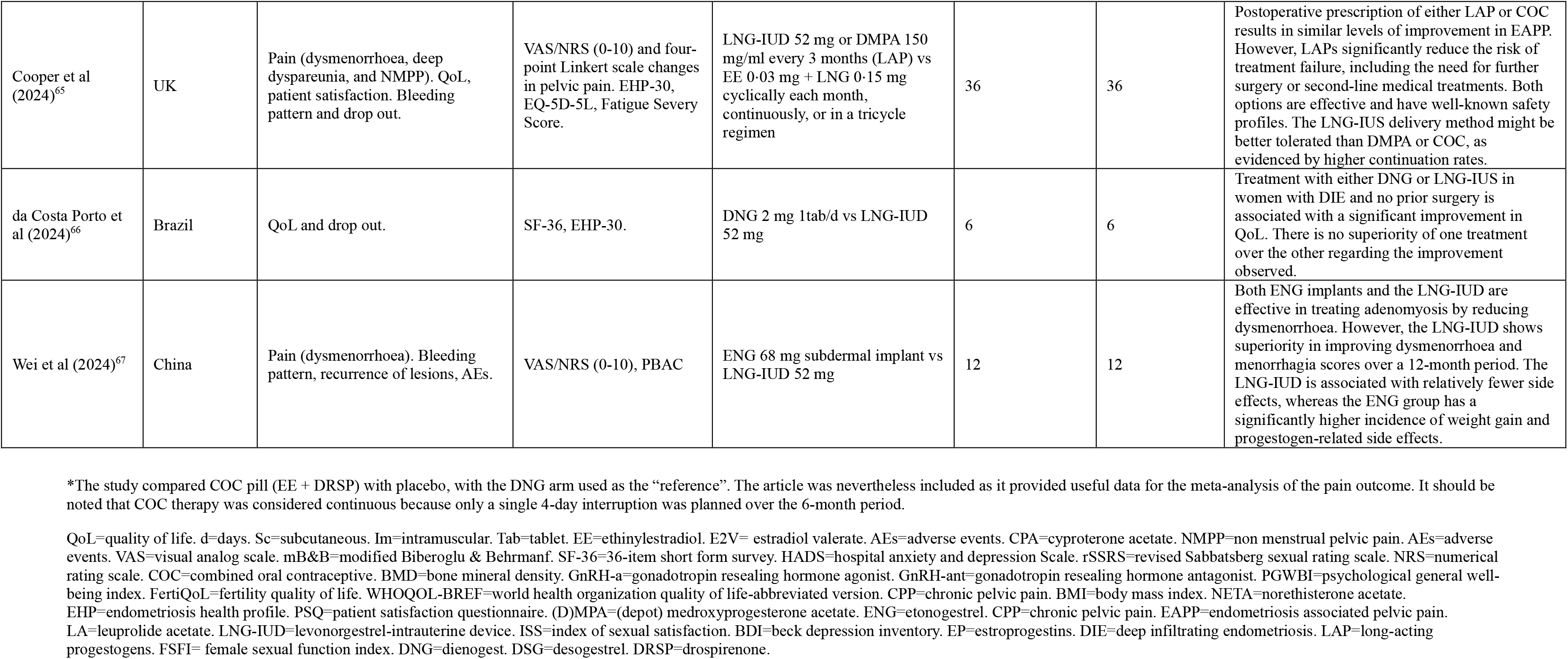
Treatment characteristics and outcomes of RCTs with hormonal therapies and endometriosis.

All medical treatments were consistently associated with significant reductions in overall pelvic pain and NMPP scores (appendix p.7-10 and Figure 2).

**Figure 2.**
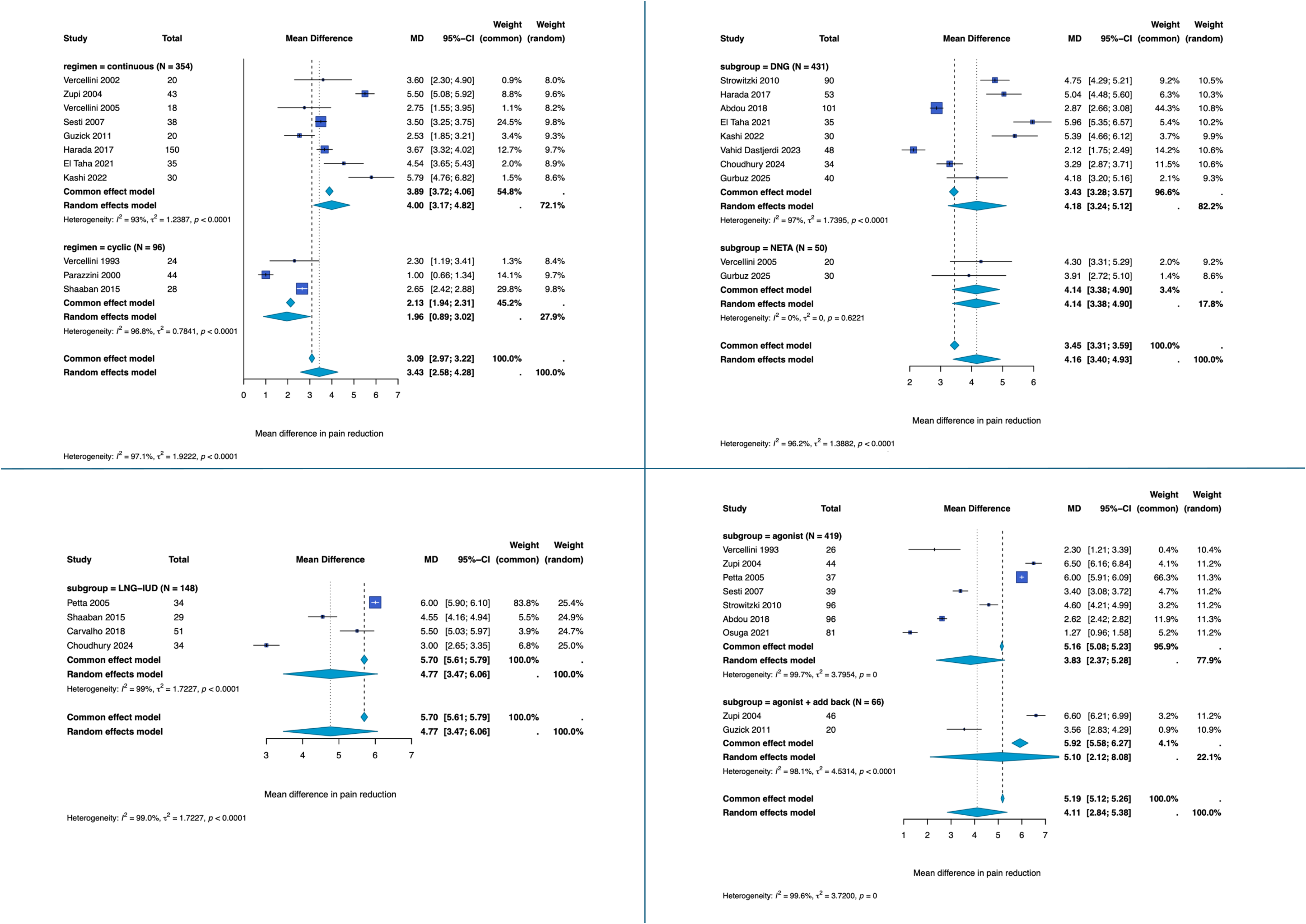
Subgroup analyses of pain outcomes according to hormonal treatment regimen. (A) Combined oral contraceptives, comparing continuous and cyclic regimens; (B) oral progestogens, comparing dienogest (DNG) and norethisterone acetate (NETA); (C) long-acting progestogens, with the subgroup analysis restricted to levonorgestrel intrauterine device (LNG-IUD); and (D) GnRH-agonists, comparing agonist monotherapy with agonist plus add-back therapy. Panels A–C refer to overall pelvic pain, whereas panel D refers to non-menstrual pelvic pain (NMPP). Pooled estimates are presented as mean differences (MDs) with 95% CIs.

Combined oral contraceptives significantly decrease overall pelvic pain among participants, with a MD of 3⋅17 points on a 0-10 scale (95% CI 2⋅24–4⋅09; *p*<0⋅0001; 12 studies^20,21,27,28,30,35,36,38,42,45,59,65^), although substantial heterogeneity was observed (*I*²=98⋅0%; appendix p.31). Subgroup analyses showed that continuous regimens resulted in greater symptom reduction than cyclic ones (MD 4⋅00, 95% CI 3⋅17–4⋅82 *vs* MD 1⋅96, 95% CI 0⋅89–3⋅02; *p*=0⋅003; Figure 2A). No evidence of small-study effects was observed (Egger’s test *p*=0⋅5691). A sensitivity analysis excluding two studies that did not report NMPP as a standalone outcome - one assessing a composite pain outcome including dysmenorrhoea^59^, and one assessing the severest EAPP^27^ - yielded comparable results (MD 3⋅17, 95% CI 2⋅05–4⋅29; 10 studies; *I*^2^=98.3%). The superiority of continuous over cyclic regimens was maintained (MD 4⋅05 vs 1⋅54; *p*=0⋅0018; *data not shown*).

Patients treated with oral progestogens exhibited a substantial decrease in overall pelvic pain scores (MD 3⋅83, 95% CI 3⋅04–4⋅62; *p*<0⋅0001; *I*²=96⋅9%; 10 studies with 12 treatment arms;^20,21,27,28,30,31,34,44,50,64^ appendix p.31). In subgroup analyses, DNG and NETA yielded comparable improvements, with no significant differences between treatments (*p*=0⋅95; Figure 2B). No clear evidence of small-study effects was detected (Egger’s test *p*=0⋅0557). Exclusion of two studies that did not specifically assess NMPP - one reporting the severest EAPP, ^27^ and one using a composite pain outcome including dysmenorrhoea,^64^ had minimal impact on the overall estimate (MD 3⋅76, 95% CI 2⋅85–4⋅67; 10 studies; *I²*=97⋅0%). The pooled effect for DNG was also consistent with the primary analysis (MD 4⋅19, 95% CI 2⋅99–5⋅39; *data not shown*).

Long-acting progestogens (DMPA, LNG-IUD and ENG) reduced overall pelvic pain by 4⋅29 points (95% CI 2⋅77–5⋅81; *p*<0⋅0001; *I*²=99⋅5%; five studies with six treatment arms;^39,59,60,64,65^ appendix p.32), with similar results in the subgroup analysis restricted to LNG-IUDs (MD 4⋅77, 95% CI 3⋅47–6⋅06; 4 studies; *p*<0⋅0001; Figure 2C). Small-study effects were not formally assessed because of the limited number of studies. After excluding the two studies that reported composite pain outcomes^59,64^ the pooled effect estimate for LAPs changed only marginally (MD 4⋅55, 95% CI 2⋅28–6⋅82; 4 studies; *I²*=99⋅6%). Comparable findings were obtained for the LNG-IUD subgroup (MD 5⋅80, 95% CI 5⋅33–6⋅28; *data not shown*).

Similarly, GnRH-analogues showed a mean reduction of 3⋅81 points in NMPP scores from baseline to study end (95% CI 2⋅54–5⋅09; *p*<0⋅0001; *I*²=99⋅7%; eight studies with 10 treatment arms^35,38,39,42,44,45,50,54^) (appendix p.32). In subgroup analyses, both GnRH-agonists alone and GnRH-agonists combined with add-back therapy were effective, without significant differences between regimens (*p*=0⋅45; Figure 2D). In this instance, no study reported composite pain outcomes, therefore no further analyses were carried out. No evidence of small-study effects was identified (Egger’s test *p*=0⋅1698).

The pooled proportion of spotting during hormonal treatment was 22⋅5% (95% CI 16⋅6– 29⋅8), based on 37 study arms including 1712 participants, with considerable heterogeneity across studies (*I*²=85⋅4%; Figure 3 and appendix p.10-13). Spotting was most frequently reported during DNG therapy (41⋅1%, 95% CI 22⋅4–62⋅9) and DMPA (40⋅4%, 95% CI 4⋅2–91⋅4), whereas lower proportions were observed with GnRH-agonists (8⋅1%, 95% CI 3⋅5–17⋅5). Intermediate proportions were reported with continuous COCs, ENG, cyclic COCs, and LNG-IUD. Pairwise comparisons demonstrated a significantly higher spotting rate with DNG than with GnRH-agonists, following multiple-testing correction (*p*=0⋅013).

**Figure 3.**
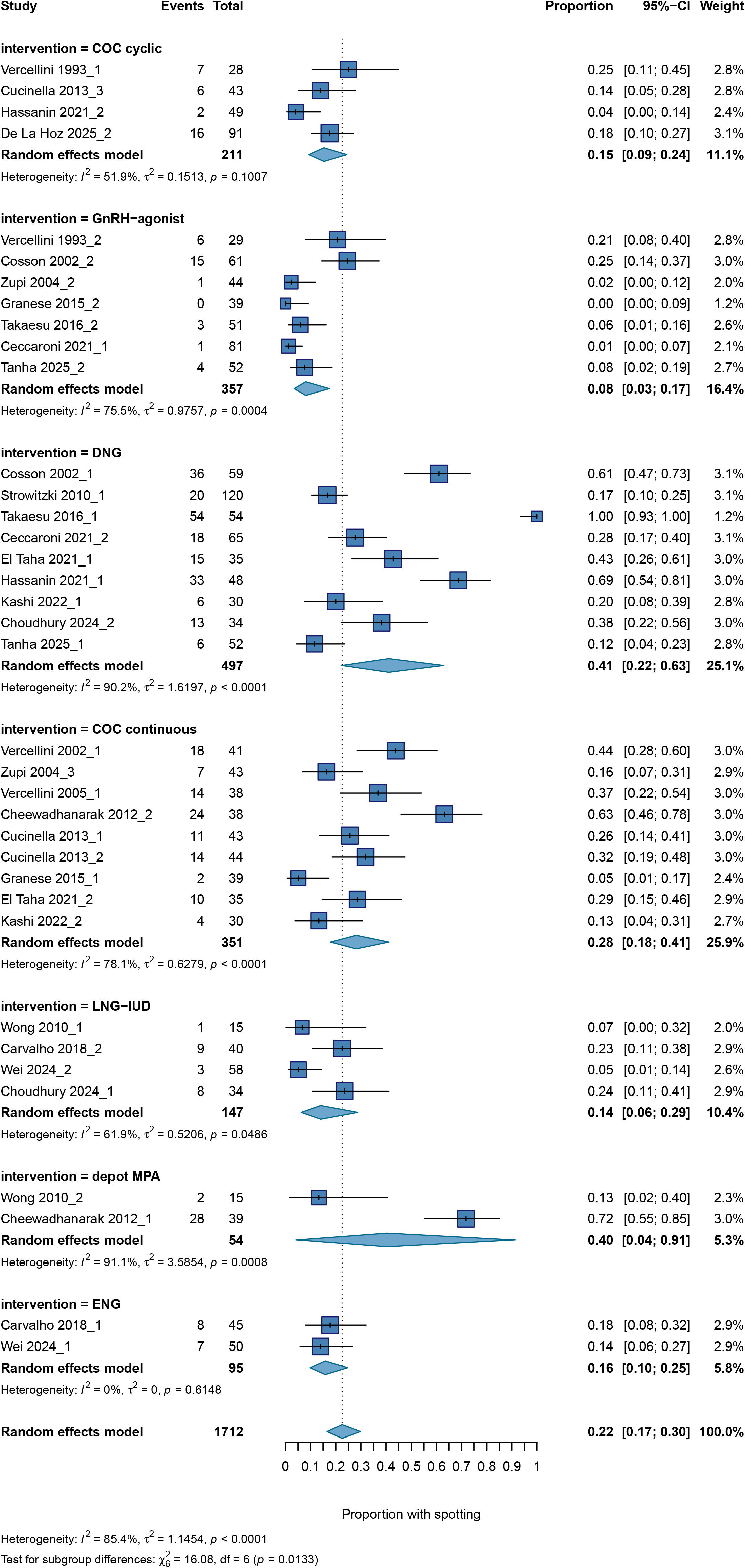
Pooled proportion of spotting across hormonal treatments. Pooled estimates are presented as proportions with 95% CIs.

The pooled proportion of breakthrough bleeding, derived from 31 study arms comprising 2049 participants, was 14⋅5% (95% CI 9⋅1–22⋅4; *I*²=90⋅9%; appendix p.33). Greater proportions were identified in patients treated with LNG-IUD (30⋅4%, 95% CI 9⋅2–65⋅2), continuous COCs (20⋅9%, 95% CI 12⋅2–33⋅6), and DNG (19⋅6%, 95% CI 5⋅5–50⋅4). Lower proportions were observed with DMPA (9⋅1%, 95% CI 6⋅1–13⋅2), GnRH-agonists (6⋅2%, 95% CI 1⋅1–28⋅5), and GnRH-antagonists (3⋅1%, 95% CI 0⋅2–39⋅7). The confidence intervals were wide for several of the treatments. Differences between treatment subgroups were not statistically significant (*p*=0⋅080).

Amenorrhoea was achieved in 51⋅4% of participants overall (95% CI 34⋅6–67⋅8; 29 study arms, 1519 participants), with considerable between-study heterogeneity (*I*²=93⋅8%; appendix p.34). The highest proportions were observed with GnRH-agonists (89⋅8%, 95% CI 70⋅3–97⋅1), and the lowest with DMPA (17⋅5%, 95% CI 12⋅9–23⋅2). Intermediate estimates were reported with ENG, DNG, continuous COCs, and LNG-IUD. Pairwise analyses showed significantly greater amenorrhoea rates with GnRH-agonists compared with DPMA, continuous COCs, DNG, and LNG-IUD after multiple-testing correction (*p*<0⋅0001).

Evidence of small-study effects was observed for spotting (Egger’s test *p*=0⋅0079) and breakthrough bleeding (*p*=0⋅0002), but not for amenorrhoea (*p*=0⋅87).

Mood changes were the most frequently reported AEs across hormonal treatments, with a pooled proportion of 15⋅2% (95% CI 9⋅9–22⋅7; Figure 4), followed by nausea (14⋅9%, 95% CI 9⋅1– 23⋅5), headache (11⋅9%, 95% CI 9⋅0–15⋅5), weight gain, decreased libido, and hot flushes/night sweats. Lower pooled proportions were observed for breast tenderness (9⋅4%, 95% CI 6⋅3–14⋅0), abdominal discomfort/bloating, vaginal dryness, acne/oily skin, insomnia, and hair loss. Heterogeneity was substantial across most analyses (appendix p.13-18 and p.35-43).

**Figure 4.**
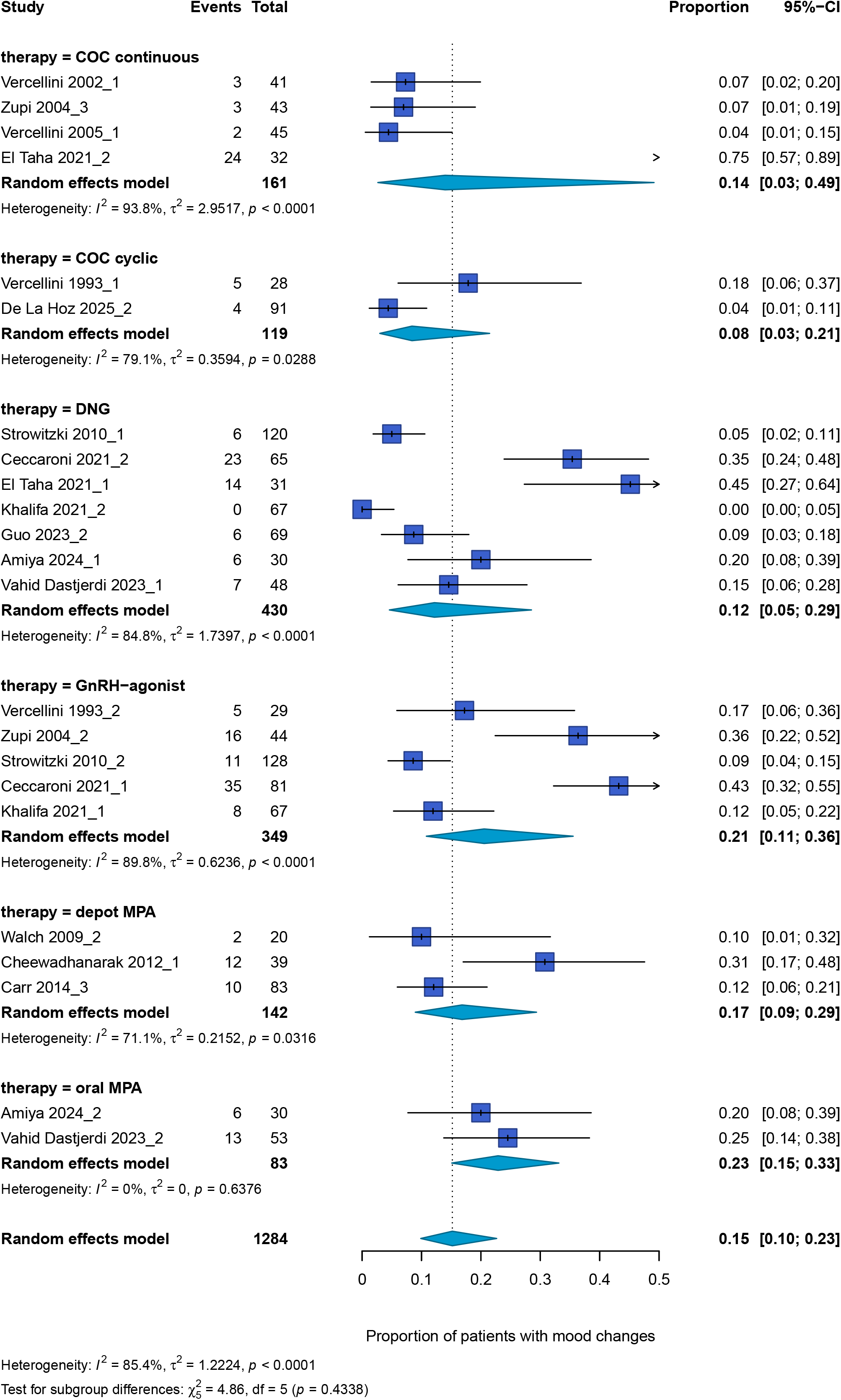
Pooled proportion of mood changes across hormonal treatments. Pooled estimates are presented as proportions with 95% CIs.

Adverse event profiles differed across hormonal treatments. GnRH-agonists were associated with markedly higher proportions of hypoestrogenic symptoms, including hot flushes/night sweats (42⋅2%, 95% CI 22⋅6–64⋅5), mood changes (20⋅6%, 95% CI 10⋅8–35⋅5), decreased libido, vaginal dryness, and insomnia. Overall, continuous COCs and DNG showed comparable AE-profiles, with similar proportions of abdominal discomfort/bloating, breast tenderness, weight gain, and mood changes, although slightly higher proportions of headaches and nausea were observed with continuous COCs. Oral MPA was associated with the highest proportion of weight gain (37⋅4%, 95% CI 27⋅7–48⋅2) and mood changes (22⋅9%, 95% CI 15⋅1–33⋅1). The observed differences between treatment classes reached statistical significance only for weight gain (*p*<0⋅0001), insomnia (*p*=0⋅0280), hot flushes/night sweats (*p*=0⋅0020), and breast tenderness (*p*=0⋅0011) (appendix p.35-43).

Evidence of small-study effects was observed for headache, mood changes, weight gain, vaginal dryness, hot flushes/night sweats, hair loss, decreased libido, breast tenderness, acne/oily skin (all Egger’s test *p*<0⋅05). No evidence of small-study effects was detected for nausea (*p*=0⋅6565) and insomnia (*p*=0⋅0677), whereas the analysis could not be performed for abdominal discomfort/bloating because of the limited number of studies.

The pooled proportion of treatment discontinuation due to any AEs, across all hormonal treatments, was 7⋅7% (95% CI 6⋅0–9⋅9; *p*<0⋅0001), based on 58 study arms including 3640 participants, with high heterogeneity across studies (*I*²=73⋅4%; Figure 5 and appendix p.19-20). Discontinuation rates were broadly comparable across treatments (*p* for subgroup differences = 0⋅23). Slightly higher proportions were observed with LNG-IUD (12⋅4%, 95% CI 8⋅0–18⋅7) and DMPA (10⋅2%, 95% CI 4⋅4-22⋅1), whereas lower proportions were reported with GnRH-agonists (4⋅7%, 95% CI 2⋅7-8⋅0) and GnRH-antagonists (5⋅0%, 95% CI 2⋅3–10⋅4). Evidence of small-study effects was identified (Egger’s test *p*<0⋅0001).

**Figure 5.**
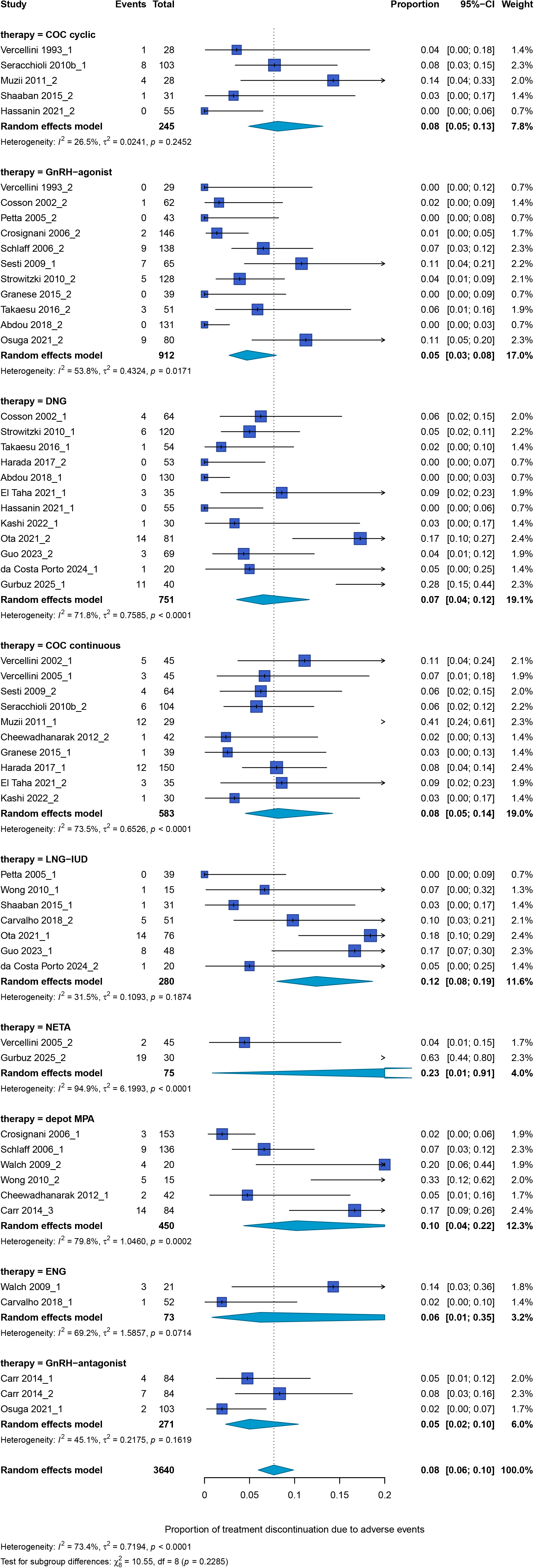
Pooled proportion of treatment discontinuation due to adverse events across hormonal treatments. Pooled estimates are presented as proportions with 95% CIs.

The risk of bias assessment identified 22 studies at high risk of bias, 14 with some concerns, and 12 at low risk of bias (appendix p.44). The evaluation of the trial’s trustworthiness revealed that 28 studies satisfied all the applicable criteria, 11 met the criteria with concerns, and 9 failed to reach the predefined trustworthiness criteria (appendix p.21-23). Overall certainty of evidence assessed using the GRADE approach was low.

In addition to overall pelvic pain and NMPP, which were quantitatively synthesised, dysmenorrhoea and deep dyspareunia were assessed qualitatively. Results for these outcomes, as reported in the original studies are summarised in appendix (p.7-10). Across studies, all hormonal treatment classes were associated with reductions in both pain domains from baseline. Direct comparisons between active treatments showed broadly similar effects, with no consistent advantage of any specific regimen (Table 2 and appendix p.7-10).

Overall, most treatment arms showed improvements in QoL, and none reported a negative effect (appendix p.24-25). Specifically, improved QoL was observed in four of five COC treatment arms, five of six oral progestogens, four of six GnRH-analogues, and all LAPs (6/6). No significant change was described in one COC and two GnRH-analogue treatment arms. The distribution of effect directions across hormonal treatment classes is illustrated in the harvest plots (Figure 6).

**Figure 6.**
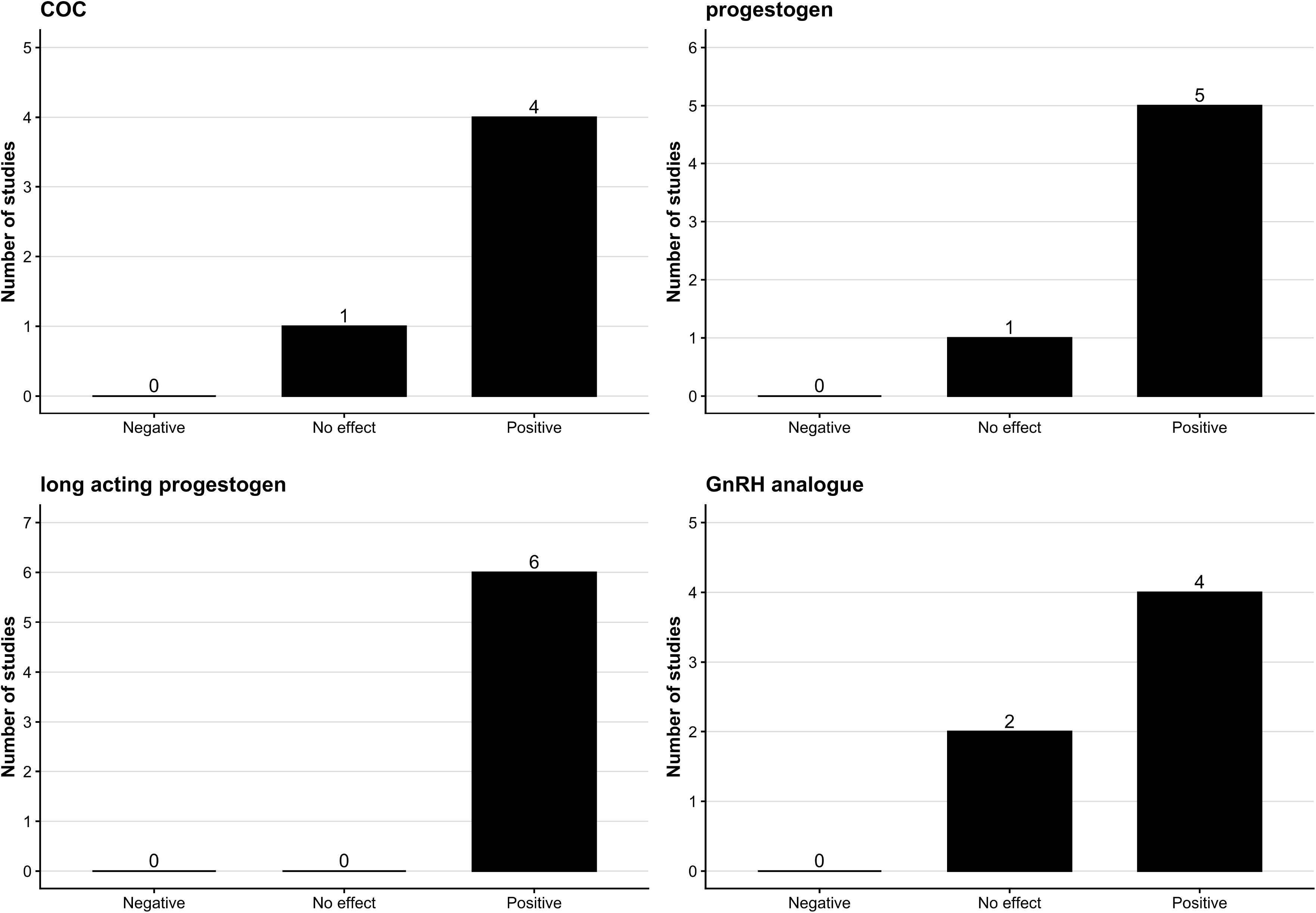
Harvest plot of the direction of effect on quality of life across hormonal treatment classes. Treatment arms were classified according to the reported direction of effect on quality of life as positive, no effect, or negative. The number of treatment arms in each category is shown for combined oral contraceptives, oral progestogens, long-acting progestogens, and GnRH-analogues.

Recurrence or worsening of symptoms and treatment discontinuation due to treatment failure were analysed separately, as clinical deterioration did not necessarily result in treatment withdrawal (appendix p.19-20 and p.26-28).^25,30,32^ Across studies, after excluding reports in which symptom outcomes were assessed during post-treatment follow-up, 96 of 1529 patients (6⋅3%) experienced recurrence or worsening of symptoms while receiving hormonal therapy (appendix p.26-28).

Assessment of imaging-confirmed disease progression or recurrence of lesions during active treatment was uncommon (appendix p.26-28). In most studies, such outcomes were assessed 6–18 months after treatment discontinuation rather than during treatment. ^25,43,49,52^ When reported, these events appeared to occur predominantly in patients with ovarian disease involvement. Only one study reported withdrawal due to disease worsening, in 9/76 LNG-IUD-treated patients with progression of endometriomas and/or adenomyosis.^62^

Reasons for treatment discontinuation are summarised in appendix (p.19-20) and were categorised as discontinuation due to AEs, treatment failure (persistent symptoms, disease progression, need for surgery, or alternative medical treatment), loss to follow-up, and other (e.g. withdrawal of consent). Across studies, discontinuation due to AEs was the most frequently reported reason, whereas discontinuation due to treatment failure was uncommon with one notable exception which reported a relatively high proportion of treatment failures requiring further surgery or second-line medical treatment, although the indications for these interventions were not clearly specified^65^. The AEs most commonly leading to treatment withdrawal, were abnormal vaginal bleeding or spotting, followed by IUD expulsion, mood changes and weight gain. Less frequent causes included headache, decreased libido, and hot flashes. Loss to follow-up and discontinuation for other reasons were reported inconsistently and generally accounted for a small proportion of treatment interruptions (appendix p.19-20).

Treatment satisfaction was reported in only nine of the 48 RCTs included in the review. Across the studies, most women reported satisfaction or very high satisfaction with therapy, with rates generally exceeding 70%. Nevertheless, comparisons between interventions were constrained by substantial heterogeneity in the instruments and criteria utilised to measure treatment satisfaction (appendix p.28-29).

In addition to QoL and treatment satisfaction, changes in analgesic use during hormonal treatment were also evaluated among the patient-reported outcomes. While no study reported the complete discontinuation of analgesic use, all trials involving non-steroidal anti-inflammatory drugs (NSAIDs) or paracetamol showed a reduction in consumption during treatment. The only exception was the study by Carr et al. (2014),^47^ which recorded a slight increase in opioid use (appendix p.29-30).

## DISCUSSION

The clinical challenge in contemporary endometriosis management is no longer establishing whether hormonal therapy is effective, but identifying which regimen offers the best balance between symptom control, tolerability, bleeding profile, and long-term acceptability. By restricting our analyses to head-to-head RCTs comparing active treatments, we found that all currently recommended hormonal therapies substantially improved overall pelvic pain and NMPP, with few clinically meaningful differences in analgesic efficacy. The one exception was that continuous COCs resulted in greater pain reduction than cyclic regimens. Both DNG and NETA, LNG-IUD and GnRH-agonists (with or without add-back therapy) achieved comparable analgesic outcomes. This suggests that, once adequate ovarian suppression is achieved, treatment selection should rely less on expectations of superior pain relief and more on tolerability, bleeding profile, patient preferences, and consequent likely long-term adherence.

Problematic bleeding was a key differentiator between interventions. Amenorrhoea was most frequent with GnRH-agonists, versus lower rates with COCs and progestogens, particularly DMPA. Spotting predominated with DNG, while breakthrough bleeding was more frequent with LNG-IUDs, despite considerable uncertainty in the pooled estimate (95% CI, 9-65%). Although rarely clinically serious, bleeding disturbances were the leading reason for treatment discontinuation.

Adverse events had differing impact on likelihood of treatment persistence. Headache, weight gain, mood-related symptoms, and spotting disturbances were among the most frequent reasons for treatment discontinuation, whereas other commonly reported AEs (i.e. nausea, decreased libido, breast tenderness) rarely resulted in cessation. These findings suggest that the impact of AEs depends not only on their frequency but also on how much they affect patients’ daily lives and treatment acceptability. However, the relatively low overall discontinuation rate due to AEs, indicates that most women perceived the hormonal therapy as acceptable. This is supported by the consistently high levels of treatment satisfaction and improvements in QoL found across hormonal treatments.

Our review’s findings align with those of previous evidence syntheses demonstrating that hormonal therapies reduce menstrual pain.^8–15^ However, our study also addressed a more specific clinical question of the impact of different hormonal treatments and regimens on non-menstrual pain (both overall pelvic pain and NMPP). Whilst no major differences in pain reduction were observed between hormonal treatment groups in our review, we observed a substantial difference between continuous and cyclic COC regimens (MD 4.00 vs 1.96 points). This contrasts with Muzii et al. (2016)^68^, who, after surgery, reported a lower recurrence of dysmenorrhoea with continuous versus cyclic COCs, but no significant difference in chronic pelvic pain. Our broader meta-analysis suggests that the potential benefit of continuous over cyclical COC administration may extend beyond menstrual pain and should be considered when selecting the hormonal regimen. Consequently, one key difference from previous reviews was the decision to use overall pelvic pain and NMPP as the primary quantitative endpoints. Although dysmenorrhoea remains a hallmark symptom of endometriosis, these outcomes may provide more informative endpoints when comparing hormonal regimens aimed at menstrual suppression.^1,69^ The present review also differed in its deliberate focus on active-treatment RCTs and clinically relevant hormonal regimens, allowing direct comparisons between drugs that clinicians would realistically consider.

Our assessment encompassed AEs, treatment discontinuation, QoL and satisfaction, alongside efficacy, recurrence and disease progression, allowing a comprehensive evaluation of hormonal therapies. The included population was clinically representative: all comprised studies enrolled women with surgically and/or imaging confirmed endometriosis, drawn from diverse study groups and geographical settings, supporting the generalisability of our findings. However, very limited data are available on the ethnicity of participants involved. Future clinical trials need to ensure that diverse populations are included and described such that clinicians can understand the relevance to their own clinical population.^70^

Nevertheless, several limitations remain. The included interventions varied in terms of doses and regimens, treatment duration and follow-up, and how outcomes were defined, scaled and measured. The timing of outcome assessment differed considerably across studies, although six months was the most frequently reported treatment duration. Short treatment lengths are particularly relevant when evaluating bleeding and AEs, as their occurrence or severity can change over time, for example rates of spotting and irregular bleeding were lower in studies with LNG-IUD treatment periods over six months. A further limitation was the inability to explore potential effect modification according to disease characteristics and treatment formulation. The available data were too scarce and heterogeneous to allow consistent stratification by endometriosis stage or phenotype, including ovarian, deep endometriosis, or adenomyosis, or by the type of oestrogen contained in COCs. Only three of the 48 RCTs investigated oestradiol valerate-containing COCs^26,33,48^, and all reported improvements in pain symptoms; however, the small number of studies precluded meaningful quantitative comparison with ethinylestradiol (EE)-containing formulations. This aspect is noteworthy given that body-identical oestrogens are generally well tolerated and may offer a more favourable safety profile than EE.^71–74^ Moreover, despite sensitivity and subgroup analyses, substantial statistical heterogeneity remained across several pooled analyses. Funnel plot asymmetry was observed for most AE outcomes, although, in the context of high heterogeneity and single-arm meta-analyses, this may reflect small-study effects rather than publication bias alone. Finally, the methodological quality of the included studies was limited, with several trials at high risk of bias and an overall low certainty of evidence according to GRADE, warranting cautious interpretation of the pooled estimates.

In conclusion, in routine practice, therapeutic decisions should be guided by the overall impact of treatment on patients, not pain improvement alone. Future trials should prioritise adequately powered head-to-head comparisons with standardised outcome definitions and assessment timepoints, together with systematic use of patient-reported outcome measures, to enable more consistent evaluation of efficacy, tolerability and patient experience across hormonal regimens.

## Data Availability

All data produced in the present work are contained in the manuscript and supplementary appendix. Protocol registered on PROSPERO.

## Contributors

AWH, LHRW and KV conceived and designed the study. VB and NS conducted the literature search and study selection, extracted the data, assessed risk of bias, and did the statistical analyses. VB, AWH, KV, and LHRW interpreted the results. VB, AWH, KV and NS drafted the manuscript. LHRW, PV, and RM critically revised the manuscript. All authors contributed to the final approval of the manuscript. VB and NS had full access to all the data in the study, directly accessed and verified the underlying data, and take responsibility for the integrity of the data and the accuracy of the data analysis. All authors had full access to the study data and had final responsibility for the decision to submit for publication.

## Declaration of interests

AWH receives grant funding from the NIHR, UKRI, Chief Scientist’s Office, Wellbeing of Women, European Union, and Roche. AWH’s institution has received honoraria for consultancy from Gedeon Richter and Theramex, and he has received lecture fees from Gedeon Richter and Theramex. AWH’s institution has received grant funding from Roche Diagnostics and honoraria for teaching from Gedeon Richter. LHRW receives grant funding the NIHR, MRC, Chief Scientist’s Office and Wellbeing of Women. LHRW’s institution has received grant funding from Roche Diagnostics, honoraria for consultancy from Thermax, Absci, Hirundo and F-prime and honoraria for teaching from Gedeon Richter and Theramex. KV receives grant funding from the NIHR, MRC, Medical Research Foundation, European Union and NIH. KV’s institution has received honoraria for consultancy and talks and associated travel costs from Gedeon Richter, Bayer Healthcare, Reckitt, Organon and Gesynta. RM previously received an honorarium from Roche Diagnostics for consultancy work. PV is a member of the Editorial Board of Human Reproduction Open, the Journal of Obstetrics and Gynaecology Canada, and the International Editorial Board of Acta Obstetricia et Gynecologica Scandinavica, and has received royalties from Wolters Kluwer for chapters on endometriosis management in the clinical decision support resource UpToDate. NS reports relationships with the World Endometriosis Society (WES), the European Society of Human Reproduction and Embryology (ESHRE), and the Society for Endometriosis and Uterine Disorders (SEUD), including travel reimbursement. VB declare no conflicts of interest.

## Data sharing

All data extracted from the included studies and used for the analyses are available in the main manuscript or appendix. No individual participant data were collected or generated for this study.

## Notes

### Competing Interest Statement

AWH receives grant funding from the NIHR, UKRI, Chief Scientists Office, Wellbeing of Women, European Union, and Roche. AWHs institution has received honoraria for consultancy from Gedeon Richter and Theramex, and he has received lecture fees from Gedeon Richter and Theramex. AWHs institution has received grant funding from Roche Diagnostics and honoraria for teaching from Gedeon Richter. LHRW receives grant funding the NIHR, MRC, Chief Scientist's Office and Wellbeing of Women. LHRWs institution has received grant funding from Roche Diagnostics, honoraria for consultancy from Thermax, Absci, Hirundo and F-prime and honoraria for teaching from Gedeon Richter and Theramex. KV receives grant funding from the NIHR, MRC, Medical Research Foundation, European Union and NIH. KV's institution has received honoraria for consultancy and talks and associated travel costs from Gedeon Richter, Bayer Healthcare, Reckitt, Organon and Gesynta. RM previously received an honorarium from Roche Diagnostics for consultancy work. PV is a member of the Editorial Board of Human Reproduction Open, the Journal of Obstetrics and Gynaecology Canada, and the International Editorial Board of Acta Obstetricia et Gynecologica Scandinavica, and has received royalties from Wolters Kluwer for chapters on endometriosis management in the clinical decision support resource UpToDate. NS reports relationships with the World Endometriosis Society (WES), the European Society of Human Reproduction and Embryology (ESHRE), and the Society for Endometriosis and Uterine Disorders (SEUD), including travel reimbursement. VB declare no conflicts of interest.

## References

1. As-Sanie S, Mackenzie SC, Morrison L, Schrepf A, Zondervan KT, Horne AW, Missmer SA. Endometriosis: A Review. JAMA. 2025;334(1):64–78.

2. Capezzuoli T, Vannuccini S, Mautone D, Sorbi F, Chen H, Reis FM, Ceccaroni M, Petraglia F. Long-term hormonal treatment reduces repetitive surgery for endometriosis recurrence. Reprod Biomed Online. 2021;42(2):451–456.

3. Taylor HS, Kotlyar AM, Flores VA. Endometriosis is a chronic systemic disease: clinical challenges and novel innovations. Lancet. 2021;397(10276):839–852.

4. Piriyev E, Schiermeier S, Römer T. Hormonal Treatment of Endometriosis: A Narrative Review. Pharmaceuticals (Basel). 2025;18(4):588.

5. Vercellini P, Salmeri N, Bandini V, Conca B, Viganò P, Somigliana E, Vignali M. Medical Treatment for Endometriosis: One Size Does Not Fit All. J Clin Med. 2026;15(6):2408.

6. Saunders PTK, Horne AW. Endometriosis: new insights and opportunities for relief of symptoms. Biol Reprod. 2025;113(5):1029–1043.

7. Vercellini P, Vercellini P, Buffo C, Viganò P, Somigliana E. Update on Medical Treatment of Endometriosis: New Drugs or New Therapeutic Approaches? Gynecol Obstet Invest. 2025;90(6):535–559.

8. Jensen JT, Schlaff W, Gordon K. Use of combined hormonal contraceptives for the treatment of endometriosis-related pain: a systematic review of the evidence. Fertil Steril. 2018;110(1):137–152.e1.

9. Benetti-Pinto CL, Mira TAA, Yela DA, Teatin-Juliato CR, Brito LGO. Pharmacological Treatment for Symptomatic Adenomyosis: A Systematic Review. Rev Bras Ginecol Obstet. 2019;41(9):564–574.

10. Grandi G, Barra F, Ferrero S, Sileo FG, Bertucci E, Napolitano A, Facchinetti F. Hormonal contraception in women with endometriosis: a systematic review. Eur J Contracept Reprod Health Care. 2019;24(1):61–70.

11. Samy A, Taher A, Sileem SA, Abdelhakim AM, Fathi M, Haggag H, Ashour K, Ahmed SA, Shareef MA, AlAmodi AA, Keshta NHA, Shatat HBAE, Salah DM, Ali AS, El Kattan EAM, Elsherbini M. Medical therapy options for endometriosis related pain, which is better? A systematic review and network meta-analysis of randomized controlled trials. J Gynecol Obstet Hum Reprod. 2021;50(1):101798.

12. Rathinam KK, Abraham JJ, S HP, S A S, Sen M, George M, A P. Evaluation of pharmacological interventions in the management of adenomyosis: a systematic review. Eur J Clin Pharmacol. 2022;78(4):531–545.

13. Csirzó Á, Kovács DP, Szabó A, Szabó B, Jankó Á, Hegyi P, Nyirády P, Ács N, Valent S. Comparative Analysis of Medical Interventions to Alleviate Endometriosis-Related Pain: A Systematic Review and Network Meta-Analysis. J Clin Med. 2024;13(22):6932.

14. Rosenberger DC, Mennicken E, Schmieg I, Medkour T, Pechard M, Sachau J, Fuchtmann F, Birch J, Schnabel K, Vincent K, Baron R, Bouhassira D, Pogatzki-Zahn EM. A systematic literature review on patient-reported outcome domains and measures in nonsurgical efficacy trials related to chronic pain associated with endometriosis: an urgent call to action. Pain. 2024;165(11):2419–2444.

15. Kou L, Huang C, Xiao D, Liao S, Li Y, Wang Q. Pharmacologic Interventions for Endometriosis-Related Pain: A Systematic Review and Meta-analysis. Obstet Gynecol. 2025;146(2):e23–e35.

16. Campbell M, McKenzie JE, Sowden A, Katikireddi SV, Brennan SE, Ellis S, Hartmann-Boyce J, Ryan R, Shepperd S, Thomas J, Welch V, Thomson H. Synthesis without meta-analysis (SWiM) in systematic reviews: reporting guideline. BMJ. 2020;368:l6890.

17. Sterne JAC, Savovic J, Page MJ, et al. RoB 2: a revised tool for assessing risk of bias in randomised trials. BMJ 2019;366(1):l4898.

18. OBGYN Editors’ Integrity Group OGEIG. Trustworthiness criteria for meta-analyses of randomized controlled studies: OBGYN journal guidelines. Eur J Obstet Gynecol Reprod Biol. 2025;305:416–418. Erratum in: Eur J Obstet Gynecol Reprod Biol. 2025;314:114670.

19. Guyatt GH, Oxman AD, Vist GE, et al. GRADE: an emerging consensus on rating quality of evidence and strength of recommendations. BMJ 2008;336(7650):924–6.

20. Vercellini P, De Giorgi O, Mosconi P, Stellato G, Vicentini S, Crosignani PG. Cyproterone acetate versus a continuous monophasic oral contraceptive in the treatment of recurrent pelvic pain after conservative surgery for symptomatic endometriosis. Fertil Steril. 2002;77(1):52–61.

21. Vercellini P, Pietropaolo G, De Giorgi O, Pasin R, Chiodini A, Crosignani PG. Treatment of symptomatic rectovaginal endometriosis with an estrogen-progestogen combination versus low-dose norethindrone acetate. Fertil Steril. 2005;84(5):1375–87.

22. Razzi S, Luisi S, Ferretti C, Calonaci F, Gabbanini M, Mazzini M, Petraglia F. Use of a progestogen only preparation containing desogestrel in the treatment of recurrent pelvic pain after conservative surgery for endometriosis. Eur J Obstet Gynecol Reprod Biol. 2007;135(2):188–90.

23. Seracchioli R, Mabrouk M, Frascà C, Manuzzi L, Montanari G, Keramyda A, Venturoli S. Long-term cyclic and continuous oral contraceptive therapy and endometrioma recurrence: a randomized controlled trial. Fertil Steril. 2010;93(1):52–6.

24. Seracchioli R, Mabrouk M, Frascà C, Manuzzi L, Savelli L, Venturoli S. Long-term oral contraceptive pills and postoperative pain management after laparoscopic excision of ovarian endometrioma: a randomized controlled trial. Fertil Steril. 2010;94(2):464–71.

25. Muzii L, Maneschi F, Marana R, Porpora MG, Zupi E, Bellati F, Angioli R, Benedetti Panici P. Oral estroprogestins after laparoscopic surgery to excise endometriomas: continuous or cyclic administration? Results of a multicenter randomized study. J Minim Invasive Gynecol. 2011;18(2):173–8. .

26. Cucinella G, Granese R, Calagna G, Svelato A, Saitta S, Tonni G, De Franciscis P, Colacurci N, Perino A. Oral contraceptives in the prevention of endometrioma recurrence: does the different progestins used make a difference? Arch Gynecol Obstet. 2013;288(4):821–7.

27. Harada T, Kosaka S, Elliesen J, Yasuda M, Ito M, Momoeda M. Ethinylestradiol 20 ìg/drospirenone 3 mg in a flexible extended regimen for the management of endometriosis-associated pelvic pain: a randomized controlled trial. Fertil Steril. 2017;108(5):798–805.

28. El Taha L, Abu Musa A, Khalifeh D, Khalil A, Abbasi S, Nassif J. Efficacy of dienogest vs combined oral contraceptive on pain associated with endometriosis: Randomized clinical trial. Eur J Obstet Gynecol Reprod Biol. 2021;267:205–212.

29. Hassanin AI, Youssef AA, Yousef AM, Ali MK. Comparison of dienogest versus combined oral contraceptive pills in the treatment of women with adenomyosis: A randomized clinical trial. Int J Gynaecol Obstet. 2021;154(2):263–269.

30. Mehdizadeh Kashi A, Niakan G, Ebrahimpour M, Allahqoli L, Hassanlouei B, Gitas G, Alkatout I. A randomized, double-blind, placebo-controlled pilot study of the comparative effects of dienogest and the combined oral contraceptive pill in women with endometriosis. Int J Gynaecol Obstet. 2022;156(1):124–132.

31. Vahid-Dastjerdi M, Hosseini R, Rodi H, Rastad H, Hosseini L. Comparison of the effectiveness of Dienogest with medroxyprogesterone acetate in the treatment of pelvic pain and recurrence of endometriosis after laparoscopic surgery. Arch Gynecol Obstet. 2023;308(1):149–155.

32. Amiya, Kaushal J, Singhal SR. A Comparative Study of Dienogest Versus Medroxyprogesterone Acetate in Endometriosis-Associated Dysmenorrhea and Menstrual Irregularities: A Randomized Trial. JK Science [Internet]. 2024;26(4):203–8.

33. De La Hoz F, Neyro JL, Santiago ALO. Evaluation of the efficacy and safety of drospirenone in women with endometriosis. Int J Clin Obstet Gynaecol 2025;9(6):1261–1268.

34. Gurbuz TB, Aslan K, Kasapoglu I, Muzii L, Uncu G. Norethindrone acetate versus dienogest for pain relief in endometriosis related pain: A randomized controlled trial. Eur J Obstet Gynecol Reprod Biol. 2025;310:113940.

35. Vercellini P, Trespidi L, Colombo A, Vendola N, Marchini M, Crosignani PG. A gonadotropin-releasing hormone agonist versus a low-dose oral contraceptive for pelvic pain associated with endometriosis. Fertil Steril. 1993;60(1):75–9.

36. Parazzini F, Di Cintio E, Chatenoud L, Moroni S, Ardovino I, Struzziero E, Falsetti L, Bianchi A, Bracco G, Pellegrini A, Bertulessi C, Romanini C, Zupi E, Massobrio M, Guidetti D, Troiano L, Beretta P, Franchi M. Estroprogestin vs. gonadotrophin agonists plus estroprogestin in the treatment of endometriosis-related pelvic pain: a randomized trial. Gruppo Italiano per lo Studio dell’Endometriosi. Eur J Obstet Gynecol Reprod Biol. 2000;88(1):11–4.

37. Cosson M, Querleu D, Donnez J, Madelenat P, Konincks P, Audebert A, Manhes H. Dienogest is as effective as triptorelin in the treatment of endometriosis after laparoscopic surgery: results of a prospective, multicenter, randomized study. Fertil Steril. 2002;77(4):684–92.

38. Zupi E, Marconi D, Sbracia M, Zullo F, De Vivo B, Exacustos C, Sorrenti G. Add-back therapy in the treatment of endometriosis-associated pain. Fertil Steril. 2004;82(5):1303–8.

39. Petta CA, Ferriani RA, Abrao MS, Hassan D, Rosa E Silva JC, Podgaec S, Bahamondes L. Randomized clinical trial of a levonorgestrel-releasing intrauterine system and a depot GnRH analogue for the treatment of chronic pelvic pain in women with endometriosis. Hum Reprod. 2005;20(7):1993–8.

40. Crosignani PG, Luciano A, Ray A, Bergqvist A. Subcutaneous depot medroxyprogesterone acetate versus leuprolide acetate in the treatment of endometriosis-associated pain. Hum Reprod. 2006;21(1):248–56.

41. Schlaff WD, Carson SA, Luciano A, Ross D, Bergqvist A. Subcutaneous injection of depot medroxyprogesterone acetate compared with leuprolide acetate in the treatment of endometriosis-associated pain. Fertil Steril. 2006;85(2):314–25.

42. Sesti F, Pietropolli A, Capozzolo T, Broccoli P, Pierangeli S, Bollea MR, Piccione E. Hormonal suppression treatment or dietary therapy versus placebo in the control of painful symptoms after conservative surgery for endometriosis stage III-IV. A randomized comparative trial. Fertil Steril. 2007;88(6):1541–7.

43. Sesti F, Capozzolo T, Pietropolli A, Marziali M, Bollea MR, Piccione E. Recurrence rate of endometrioma after laparoscopic cystectomy: a comparative randomized trial between post-operative hormonal suppression treatment or dietary therapy vs. placebo. Eur J Obstet Gynecol Reprod Biol. 2009;147(1):72–7.

44. Strowitzki T, Marr J, Gerlinger C, Faustmann T, Seitz C. Dienogest is as effective as leuprolide acetate in treating the painful symptoms of endometriosis: a 24-week, randomized, multicentre, open-label trial. Hum Reprod. 2010;25(3):633–41.

45. Guzick DS, Huang LS, Broadman BA, Nealon M, Hornstein MD. Randomized trial of leuprolide versus continuous oral contraceptives in the treatment of endometriosis-associated pelvic pain. Fertil Steril. 2011;95(5):1568–73.

46. Bayoglu Tekin Y, Dilbaz B, Altinbas SK, Dilbaz S. Postoperative medical treatment of chronic pelvic pain related to severe endometriosis: levonorgestrel-releasing intrauterine system versus gonadotropin-releasing hormone analogue. Fertil Steril. 2011;95(2):492–6.

47. Carr B, Dmowski WP, O’Brien C, Jiang P, Burke J, Jimenez R, Garner E, Chwalisz K. Elagolix, an oral GnRH antagonist, versus subcutaneous depot medroxyprogesterone acetate for the treatment of endometriosis: effects on bone mineral density. Reprod Sci. 2014;21(11):1341–51.

48. Granese R, Perino A, Calagna G, Saitta S, De Franciscis P, Colacurci N, Triolo O, Cucinella G. Gonadotrophin-releasing hormone analogue or dienogest plus estradiol valerate to prevent pain recurrence after laparoscopic surgery for endometriosis: a multi-center randomized trial. Acta Obstet Gynecol Scand. 2015;94(6):637–45.

49. Takaesu Y, Nishi H, Kojima J, Sasaki T, Nagamitsu Y, Kato R, Isaka K. Dienogest compared with gonadotropin-releasing hormone agonist after conservative surgery for endometriosis. J Obstet Gynaecol Res. 2016;42(9):1152–8.

50. Abdou AM, Ammar IMM, Alnemr AAA, Abdelrhman AA. Dienogest Versus Leuprolide Acetate for Recurrent Pelvic Pain Following Laparoscopic Treatment of Endometriosis. J Obstet Gynaecol India. 2018;68(4):306–313.

51. Ozaki R, Kumakiri J, Jinushi M, Ikuma S, Murakami K, Kawasaki Y, Kitade M. Comparison of effect of preoperative dienogest and gonadotropin-releasing hormone agonist administration on laparoscopic cystectomy for ovarian endometriomas. Arch Gynecol Obstet. 2020;302(4):969–976.

52. Ceccaroni M, Clarizia R, Liverani S, Donati A, Ceccarello M, Manzone M, Roviglione G, Ferrero S. Dienogest vs GnRH agonists as postoperative therapy after laparoscopic eradication of deep infiltrating endometriosis with bowel and parametrial surgery: a randomized controlled trial. Gynecol Endocrinol. 2021;37(10):930–933.

53. Khalifa E, Mohammad H, Abdullah A, Abdel-Rasheed M, Khairy M, Hosni M. Role of suppression of endometriosis with progestins before IVF-ET: a non-inferiority randomized controlled trial. BMC Pregnancy Childbirth. 2021;21(1):264.

54. Osuga Y, Seki Y, Tanimoto M, Kusumoto T, Kudou K, Terakawa N. Relugolix, an oral gonadotropin-releasing hormone (GnRH) receptor antagonist, in women with endometriosis-associated pain: phase 2 safety and efficacy 24-week results. BMC Womens Health. 2021;21(1):250.

55. Tanha FD, Rasti A, Pakniat H, Setudeh SS. The effect of dienogest and gonadotropin-releasing hormone agonist on pelvic pain after laparoscopic surgery for endometriosis: An RCT. Int J Reprod Biomed. 2025;22(12):995–1002.

56. Walch K, Unfried G, Huber J, Kurz C, van Trotsenburg M, Pernicka E, Wenzl R. Implanon versus medroxyprogesterone acetate: effects on pain scores in patients with symptomatic endometriosis--a pilot study. Contraception. 2009;79(1):29–34.

57. Wong AY, Tang LC, Chin RK. Levonorgestrel-releasing intrauterine system (Mirena) and Depot medroxyprogesterone acetate (Depoprovera) as long-term maintenance therapy for patients with moderate and severe endometriosis: a randomised controlled trial. Aust N Z J Obstet Gynaecol. 2010;50(3):273–9.

58. Cheewadhanaraks S, Choksuchat C, Dhanaworavibul K, Liabsuetrakul T. Postoperative depot medroxyprogesterone acetate versus continuous oral contraceptive pills in the treatment of endometriosis-associated pain: a randomized comparative trial. Gynecol Obstet Invest. 2012;74(2):151–6.

59. Shaaban OM, Ali MK, Sabra AM, Abd El Aal DE. Levonorgestrel-releasing intrauterine system versus a low-dose combined oral contraceptive for treatment of adenomyotic uteri: a randomized clinical trial. Contraception. 2015;92(4):301–7.

60. Carvalho N, Margatho D, Cursino K, Benetti-Pinto CL, Bahamondes L. Control of endometriosis-associated pain with etonogestrel-releasing contraceptive implant and 52-mg levonorgestrel-releasing intrauterine system: randomized clinical trial. Fertil Steril. 2018;110(6):1129–1136.

61. Margatho D, Carvalho NM, Bahamondes L. Endometriosis-associated pain scores and biomarkers in users of the etonogestrel-releasing subdermal implant or the 52-mg levonorgestrel-releasing intrauterine system for up to 24 months. Eur J Contracept Reprod Health Care. 2020;25(2):133–140.

62. Ota I, Taniguchi F, Ota Y, Nagata H, Wada I, Nakaso T, Ikebuchi A, Sato E, Azuma Y, Harada T. A controlled clinical trial comparing potent progestins, LNG-IUS and dienogest, for the treatment of women with adenomyosis. Reprod Med Biol. 2021;20(4):427–434.

63. Guo W, Lin Y, Hu S, Shen Y. Compare the Efficacy of Dienogest and the Levonorgestrel Intrauterine System in Women with Adenomyosis. Clin Ther. 2023;45(10):973–976.

64. Choudhury S, Jena SK, Mitra S, Padhy BM, Mohakud S. Comparison of efficacy between levonorgestrel intrauterine system and dienogest in adenomyosis: a randomized clinical trial. Ther Adv Reprod Health. 2024;18:26334941241227401.

65. Cooper KG, Bhattacharya S, Daniels JP, Horne AW, Clark TJ, Saridogan E, Cheed V, Pirie D, Melyda M, Monahan M, Roberts TE, Cox E, Stubbs C, Middleton LJ; PRE-EMPT Collaborative Group. Long acting progestogens versus combined oral contraceptive pill for preventing recurrence of endometriosis related pain: the PRE-EMPT pragmatic, parallel group, open label, randomised controlled trial. BMJ. 2024;385:e079006.

66. da Costa Porto BT, Ribeiro PA, Kuteken F, Ohara F, Abdalla Ribeiro HS. Levonorgestrel intrauterine system versus dienogest effect on quality of life of women with deep endometriosis: a randomized open-label clinical trial. Women Health. 2024;64(7):551–558.

67. Wei A, Tang X, Yang W, Zhou J, Zhu W, Pan S. Efficacy of etonogestrel subcutaneous implants versus the levonorgestrel-releasing intrauterine system in the conservative treatment of adenomyosis. Open Med (Wars). 2024;19(1):20240914.

68. Muzii L, Di Tucci C, Achilli C, Di Donato V, Musella A, Palaia I, Panici PB. Continuous versus cyclic oral contraceptives after laparoscopic excision of ovarian endometriomas: a systematic review and metaanalysis. Am J Obstet Gynecol. 2016;214(2):203–211.

69. Vercellini P, Buggio L, Berlanda N, Barbara G, Somigliana E, Bosari S. Estrogen-progestins and progestins for the management of endometriosis. Fertil Steril. 2016;106(7):1552–1571.e2.

70. Khan Z, Vincent K, Rai T, Dixon S. A lack of sociodemographic participant diversity in endometriosis evidence risks unrepresentative clinical guidance: a structured review of the evidence contributing to a NICE guideline. Br J Gen Pract. 2024;74(suppl 1):bjgp24X737697.

71. Kobayashi T, Hirayama M, Nogami M, et al. Impact of estetrol combined with drospirenone on blood coagulation and fibrinolysis in patients with endometriosis: a multicenter, randomized, open-label, active-controlled, parallel-group study. Clin Appl Thromb Hemost. 2024;30:10760296241286514.

72. Farris M, Bastianelli C, Habiba M, Benagiano G. Hormonal contraception, past, present, and future part 2: optimizing combined oral contraceptives to decrease risks for healthy women. Expert Rev Clin Pharmacol. 2025;18(6):361–372.

73. Harada T, Nogami M, Iizuka M, Meguro K, Manda C, Hirayama M, Kobayashi T. Impact of estetrol and drospirenone combination therapy on alleviation of the pelvic pain of endometriosis: a randomized control trial. F S Rep. 2025;7(1):38–45.

74. Raskin L, Didembourg M, Dogné JM, et al. Venous thromboembolism with combined oral contraceptives based on estrogen and progestin content: a disproportionality analysis of the FDA adverse event reporting system database. Am J Obstet Gynecol. 2026;235(1):78–85.

