## Supplementary appendix 1 for "HORMONAL THERAPIES FOR ENDOMETRIOSIS: A SYSTEMATIC REVIEW AND META-ANALYSIS OF RANDOMISED HEAD-TO-HEAD TRIALS"

**Tables**

**PubMed Search strategy**

(Endometriosis[Mesh] OR Adenomyosis[Mesh] OR endometriosis[tiab] OR adenomyosis[tiab]) AND ("Progestins"[Mesh] OR Contraceptive Agents[Mesh] OR Gonadotropin-Releasing Hormone[Mesh] OR "Intrauterine Devices, Medicated"[Mesh] OR progestin*[tiab] OR POP[tiab] OR desogestrel[tiab] OR norethisterone[tiab] OR NETA[tiab] OR norethindrone[tiab] OR drospirenone[tiab] OR dienogest[tiab] OR DNG[tiab] OR medroxyprogesterone[tiab] OR MPA[tiab] OR combined oral contraceptive*[tiab] OR COC[tiab] OR combined pill*[tiab] OR contraceptive ring[tiab] OR vaginal ring[tiab] OR levonorgestrel intrauterine system[tiab] OR LNG-IUD[tiab] OR hormonal IUD[tiab] OR GnRH-a*[tiab] OR contraceptive patch[tiab] OR transdermal contraceptive patch[tiab] OR etonogestrel implant[tiab] OR subdermal implant[tiab]) AND ((randomized controlled trial[pt] OR randomized[tiab])

Gender/sex were not explicitly included in search terms, but studies typically referred to women, rather than women and those assigned female at birth. Other non-pharmacological and pharmacological interventions outside hormonal therapy were not considered. Similarly, danazol, gestrinone and aromatase inhibitors were not included. The androgenic adverse effects associated with danazol and gestrinone limit their role in contemporary clinical practice, while aromatase inhibitors are only recommended for those with symptoms refractory to other medical/surgical intervention (Becker et al., 2022).

**Study selection and data extraction**

Study selection was independently performed by two reviewers (V.B. and N.S.) through title and abstract screening followed by full-text assessment. Disagreements were resolved by discussion and consensus. Studies were included in the meta-analysis only when sufficient and homogeneous quantitative data for effect size calculation were available. Only those treatment regimens represented by ≥2 study arms were analysed in the forest plots; therefore, regimens for which fewer than two study arms were available (e.g. for desogestrel, drospirenone, and GnRH antagonists) were not pooled and are not shown in the corresponding plots. Data from individual studies were retained in the corresponding supplementary tables. For qualitative comparison, only outcome measures and questionnaire scores reported in at least two studies were retained. When multiple publications reported data from the same study population, data were extracted only from the publication providing the most complete outcome data or the longest follow-up. In cases where different publications from the same cohort reported distinct outcomes, both reports were considered, and each outcome was recorded only once to avoid double counting.

Data from studies evaluating postoperative hormonal treatment were not excluded. Although previous surgery may have exerted a cumulative effect on symptom control, inclusion of these participants was considered more representative of real-world clinical practice. For studies evaluating GnRH antagonists, only treatment arms including approved dosages and regimens were retained for analysis, whereas experimental or non-commercialised regimens were excluded.

Data extracted from each study included country, sample size, participant age, body mass index, type and stage of disease, diagnostic modality, intervention and comparator regimens, treatment duration, follow-up length, and outcome data. Outcome data comprised pain domains (dysmenorrhoea, dyspareunia, and non-menstrual pelvic pain), bleeding patterns, adverse events (AEs), treatment discontinuation, quality of life (QoL), recurrence of lesions or symptoms, treatment satisfaction and change in analgesic use.

- **Pain outcomes (Table S1)**

Pain domains evaluated include overall pelvic pain, non-menstrual pelvic pain (NMPP) dysmenorrhoea and deep dyspareunia. The former represented the broader category and comprised not only studies assessing NMPP but also composite pelvic pain measures (e.g. combined dysmenorrhoea/chronic pelvic pain), and the severest endometriosis-associated pelvic pain (EAPP), as reported by the original studies.

For the purposes of the meta-analysis, the primary analysis assessed the "overall pelvic pain" domain, including studies with the evaluation of EAPP or composite outcomes measures, whereas the sensitivity analyses retained only those studies that measured NMPP as a standalone outcome. Overall pelvic pain and NMPP outcomes were analysed quantitatively, as such outcomes can reflect the global pain burden related to endometriosis, providing the most comprehensive measure of treatment response across hormonal therapies, whereas dysmenorrhoea and deep dyspareunia were summarised only qualitatively.

Only studies reporting baseline and post-treatment overall pelvic pain or NMPP values as means or medians with corresponding standard deviations (SDs), ranges, or interquartile ranges were included in the meta-analyses (i.e. studies reporting only percentage pain reduction or values at study end, without baseline quantitative scores were excluded). For consistency in tabulation and subsequent analyses, overall pelvic pain and NMPP scales reported by the original authors as VAS (Visual Analog Scale), NRS (Numerical Rating Scare), or "linear analogue scales" using a 0–10 scoring system were grouped together. When pain outcomes were reported on a 0–100 VAS, values were rescaled to a 0–10 scale to ensure comparability across studies for the purposes of the meta-analysis. When studies reported medians with interquartile ranges or ranges, these were converted to means and SDs using validated quantile estimation methods implemented in R (Wan et al., 2014; Luo et al., 2018). One study (Cheewadhanara et al., 2012) was excluded from the quantitative synthesis due to instability of the estimates following transformation.

Pain outcomes assessed using the modified Biberoglu and Behrman and modified Andersch and Milsom scales were synthesised qualitatively only because of the limited number of studies using the latter scale and the heterogeneity in reporting among studies using the former (i.e. individual domains, composite scores, and different score systems).

- **Bleeding patterns (Table S2)**

Bleeding patterns were classified into amenorrhoea, spotting and breakthrough bleeding according to the terminology used in the original articles. Reports of "spotting" or "mild bleeding" were assigned to the spotting group, whereas breakthrough bleeding comprised cases described as "breakthrough bleeding", "intermenstrual bleeding", "uterine or vaginal haemorrhage", or "menorrhagia/metrorrhagia". Bleeding outcomes were not recorded when the severity or type of bleeding could not be reliably classified.

When studies reported bleeding outcomes only as percentages, raw event numbers were calculated based on the total number of participants in each treatment arm.

- **Adverse events (Table S3-S4)**

Adverse events were grouped according to individual event type whenever reported. Only the most frequently and consistently reported AEs across studies were tabulated and included in the meta-analysis. When AEs were reported only as percentages, raw event counts were calculated through the total number of participants in the corresponding treatment arm for meta-analysis purposes.

- **Discontinuation of treatments (Table S5)**

Reasons for treatment discontinuation were categorised into four groups: AEs, treatment failure, loss to follow-up, and other reasons (i.e. withdrawal of consent or protocol deviations). A meta-analysis was carried out on the former, whilst for the other three categories only a qualitative description was provided. Whenever available, the specific AEs leading to treatment discontinuation were also recorded.

- **Quality of life (Table S7-S9)**

Regarding QoL outcomes, summary and specific principal domains scores from validated questionnaires were extracted whenever reported. Modular questionnaire components and subdomains were not tabulated. Supplementary tables were organised according to the instrument used, including 36-Item Short Form Health Survey (SF-36), 30-Item Endometriosis Health Profile (EHP-30), and World Health Organization Quality of Life - Brief Version (WHOQOL-BREF).

- **Recurrence of lesions or reappearance of symptoms (Table S10)**

The studies included in this category reported results relating to the recurrence or progression of endometriotic lesions as assessed by imaging scans during treatment or follow-up. Reappearance or worsening of symptoms was extracted when studies reported persistent or recurrent symptoms, treatment failure, lack of efficacy, pain relapse, or treatment withdrawal attributable to symptom persistence.

Given the substantial heterogeneity across studies, recurrence outcomes were summarised descriptively rather than quantitatively pooled. Considerable variability was observed regarding:

(i) the definition and assessment of lesion recurrence, including variability in imaging methods, lesion type, and reporting of baseline disease characteristics;

(ii) the definition of symptom recurrence, which was inconsistently reported across studies and relied on heterogeneous indicators of treatment failure rather than standardised assessments;

(iii) the timing of recurrence assessment for both endometriotic lesions and symptoms, which in some studies occurred several months after treatment discontinuation.

- **Patient satisfaction (Table S11)**

Patient-reported treatment satisfaction was also extracted. Reporting methods were highly heterogeneous across studies and included dichotomous outcomes, ordinal satisfaction scales, and multi-category rating systems.

- **Change in analgesic use during treatment (Table S12)**

Data regarding analgesic use before and during hormonal treatment were also obtained whenever available.

**Table S1 Baseline and post-treatment pain scores across hormonal therapies for endometriosis**

| Study | TX length (months) | Intervention | *n* | Dysmenorrhea | | | *n* | Non-menstrual pelvic pain | | | *n* | Deep dyspareunia | | |
|---|---|---|---|---|---|---|---|---|---|---|---|---|---|---|
| | | | | Baseline | Post-TX | *p* value | | Baseline | Post-TX | *p* value | | Baseline | Post-TX | *p* value |
| **VAS 0-100 mm** | | | | | | | | | | | | | | |
| Vercellini *et al.*, 2002 | 6 | COC continuous | 36 | 74 (59-83) | 0 (0-1) | ns | 20 | 47 (27-72) | 20 (0-30) | ns | 25 | 51 (30-67) | 15 (0-20) | ns |
| | | CPA | 39 | 71 (65-83) | 0 (0-0) | ns | 22 | 54 (41-65) | 14 (0-40) | ns | 23 | 52 (34-70) | 13 (10-30) | ns |
| Vercellini *et al.*, 2005 | 12 | COC continuous | 34 | 72·3 ± 16·6 | 8·7 ± 20·7 | <0·05 | 18 | 52·5 ± 23·7 | 25·0 ± 27·9 | <0·05 | 23 | 46·5 ± 22·1 | 10·8 ± 22·9 | <0·05 |
| | | NETA | 37 | 75·8 ± 18·1 | 3·0 ± 11·3 | <0·05 | 20 | 57·5 ± 24·0 | 14·5 ± 20·9 | <0·05 | 25 | 51·4 ± 24·7 | 13·8 ± 23·0 | <0·05 |
| Strowitzki *et al.*, 2010 | 6 | DNG | NA | NA | NA | NA | 90 | 60·2 ± 24·2 | 12·7 ± 20·3 | NA | NA | NA | NA | NA |
| | | GnRH-agonist | NA | NA | NA | NA | 96 | 57·9 ± 21·0 | 11·9 ± 16·9 | NA | NA | NA | NA | NA |
| Harada *et al.*, 2017* | 6 | COC continuous | NA | NA | NA | NA | 150 | 77·2 ± 16·5 | 40·5 ± 25·1 | NA | NA | NA | NA | NA |
| | | DNG | NA | NA | NA | NA | 53 | 76·3 ± 16·5 | 25·9 ± 23·5 | NA | NA | NA | NA | NA |
| Abdou *et al.*, 2018 | 3 | DNG | NA | NA | NA | NA | 101 | 59·27 ± 11·02 | 30·61 ± 10·65 | <0·001 | 55 | 36·53 ± 3·87 | 16·53 ± 3·10 | <0·001 |
| | | GnRH-a | NA | NA | NA | NA | 96 | 58·73 ± 11·01 | 32·53 ± 8·74 | <0·001 | 62 | 34·98 ± 4·96 | 17·11 ± 2·53 | <0·001 |
| Osuga *et al.*, 2021a | 6 | GnRH-ant | 103 | 30·4 ± 17·0 | [-29·5 (±17·54)] | NA | 103 | 15·3 ± 12·0 | [-11·9 (± 11·26)] | NA | 44 | 9·4 ± 15·4 | [-0·9 (±12·04)] | NA |
| | | GnRH-a | 81 | 27·1 ± 19·8 | [-27·2 (±19·86)] | NA | 81 | 15·2 ± 15·1 | [-12·7 (±12·57)] | NA | 26 | 9·5 ± 10·7 | [-4·6 (±15·09)] | NA |
| **NRS/VAS 0-10** | | | | | | | | | | | | | | |
| Vercellini et al., 1993 | 6 | low dose cyclic OC | 24 | 8·0 ± 1·9 | 3·7 ± 2·1 | <0·01 | 24 | 4·2 ± 3·0 | 1·9 ± 2·5 | <0·01 | 21 | 6·1 ± 3·3 | 3·9 ± 2·9 | <0·01 |
| | | GnRH-a | 26 | 8·1 ± 2·4 | NA | | 26 | 4·4 ± 3·2 | 2·1 ± 2·2 | <0·01 | 22 | 6·4 ± 3·0 | 2·1 ± 2·5 | <0·01 |
| Parazzini et al., 2000 | 12 | cyclic OC | 47 | 6 (0-8) | 6 (2-8) | ns | 44 | 5 (2-7) | 4 (2-5) | ns | NA | NA | NA | NA |
| | | GnRH-a/ cyclic OC | 55 | 6 (1-10) | 4 (1-7) | ns | 55 | 6 (2-9) | 6 (2-8) | ns | NA | NA | NA | NA |
| Zupi et al., 2004 | 12 | GnRHa + add back | 46 | 5·8 ± 1·6 | 0·0 ± 0·0 | 0·01 | 46 | 6·9 ± 1·4 | 0·3 ± 0·1 | 0·01 | 46 | 5·8 ± 1·6 | 1·2 ± 0·6 | 0·01 |
| | | GnRH-a | 44 | 6·1 ± 1·4 | 0·0 ± 0·0 | 0·01 | 44 | 6·7 ± 1·2 | 0·2 ± 0·1 | 0·01 | 44 | 5·9 ± 1·5 | 1·4 ± 0·5 | 0·01 |
| | | COC continuous | 43 | 6·0 ± 1·8 | 0·9 ± 0·5 | 0·01 | 43 | 6·3 ± 1·6 | 0·8 ± 0·5 | 0·01 | 43 | 5·6 ± 1·2 | 1·3 ± 0·6 | 0·01 |
| Petta et al., 2005 | 6 | LNG-IUD | NA | NA | NA | NA | 34 | 7·3 ± 0·3 | [-6 ± (0·3)] | <0·05 | NA | NA | NA | NA |

| | | | | | | | | | | | | | | |
|---|---|---|---|---|---|---|---|---|---|---|---|---|---|---|
| | | GnRH-a | NA | NA | NA | NA | 37 | 7·3 ± 0·3 | [-6 ± (0·2)] | <0·05 | NA | NA | NA | NA |
| Sesti et al., 2007 | 6 | GnRH-a | 39 | 7·7 1·0 | 5·9 0·9 | <0·05 | 39 | 8·4 ± 0·9 | 5·0 ± 1·1 | <0·05 | 39 | 6·9 1·0 | 4·3 1·2 | <0·05 |
| | | COC continuous | 38 | 8·2 1·1 | 5·5 1·2 | <0·05 | 38 | 8·5 ± 0·8 | 5·0 ± 0·8 | <0·05 | 38 | 6·8 1·2 | 4·5 1·3 | <0·05 |
| Guzick et al., 2011† | 12 | COC continuous | NA | NA | NA | NA | 20 | 4·30 ±1·77 | [-2·53 ± (0·59)] | <0·001 | NA | NA | NA | NA |
| | | GnRH-a + add back | NA | NA | NA | NA | 20 | 4·83 ± 1·87 | [-3·56 ± (0·54)] | <0·001 | NA | NA | NA | NA |
| Cheewadhanarak et al., 2012 | 6 | DMPA | 42 | 9 (7–10) | 0 (0–0) | <0·05 | 42 | 2·5 (0–6·8) | 0 (0–0) | <0·05 | 42 | 3 (0–5) | 0 (0–2) | <0·05 |
| | | COC continuous | 42 | 8·2 (7–10) | 0 (0–3) | <0·05 | 42 | 2 (0–6·4) | 0 (0–0·4) | <0·05 | 42 | 4·5 (0–7) | 0 (0–0) | <0·05 |
| Shaaban et al., 2015‡ | 6 | LNG-IUD | NA | NA | NA | NA | 29 | 6·23 ± 0·67 | 1·68 ± 1·25 | <0·001 | NA | NA | NA | NA |
| | | COC cyclic | NA | NA | NA | NA | 28 | 6·55 ± 0·68 | 3·90 ± 0·54 | <0·001 | NA | NA | NA | NA |
| Carvalho et al., 2018 | 6 | ENG | 52 | 7·5 ± 1·7 | 2·2 ± 3·2 | <0·001 | 52 | 7·6 ± 1·7 | 2·0 ± 2·4 | <0·001 | NA | NA | NA | NA |
| | | LNG-IUD | 51 | 7·3 ± 1·7 | 1·9 ± 2·2 | <0·001 | 51 | 7·4 ± 1·7 | 1·9 ± 1·7 | <0·001 | NA | NA | NA | NA |
| El Taha et al., 2021§ | 6 | DNG | NA | NA | NA | NA | 35 | 8·40 ± 1·3 | 2·44 ± 2·1 | <0·001 | NA | NA | NA | NA |
| | | COC continuous | NA | NA | NA | NA | 35 | 7·92 ± 1·5 | 3·38 ± 3·1 | <0·001 | NA | NA | NA | NA |
| Hassanin et al., 2021 | 6 | DNG | 48 | 6·27±1·21 | 3·21±1·18 | <0·001 | NA | NA | NA | NA | NA | NA | NA | NA |
| | | COC cyclic | 49 | 6·11±1·13 | 4·92±1·22 | <0·001 | NA | NA | NA | NA | NA | NA | NA | NA |
| Mehdizadeh Kashi et al., 2022 | 6 | DNG | NA | NA | NA | NA | 30 | 8·59 ± 2·25 | 3·2 ± 1·77 | 0·01 | 30 | 4·81 ± 0·89 | 2·67 ± 1·83 | ns |
| | | COC continuous | NA | NA | NA | NA | 30 | 7·93 ± 3·22 | 2·14 ± 2·3 | 0·01 | 30 | 4·56 ± 1·27 | 1·7 ± 1·41 | 0·04 |
| Vahid Dastjerdi et al., 2023 | 6 | DNG | 48 | 8·58 ± 1·02 | 5·22 ± 1·30 | <0·001 | 48 | 6·72 ± 1·44 | 4·60 ± 1·16 | <0·001 | 48 | 6·67 ± 2·19 | 4·80 ± 1·40 | <0·001 |
| | | oral MPA | 53 | 8·84 ± 0·71 | 5·54 ± 0·74 | <0·001 | 53 | 7·08 ± 1·03 | 5·30 ± 0·93 | <0·001 | 53 | 6·67 ± 2·14 | 4·71 ± 1·39 | <0·001 |
| Amiya et al., 2024 | 3 | DNG | 30 | 5·73 ± 0·38 | 0·23 ± 0·11 | <0·001 | NA | NA | NA | NA | NA | NA | NA | NA |
| | | oral MPA | 30 | 5·3 ± 0·58 | 0·8 ± 0·26 | <0·001 | NA | NA | NA | NA | NA | NA | NA | NA |
| Choudhury et al., 2024 ¶ | 3 | LNG-IUD | NA | NA | NA | NA | 34 | 6·41 ± 1·07 | 3·41 ± 1·04 | <0·001 | NA | NA | NA | NA |
| | | DNG | NA | NA | NA | NA | 34 | 6·41 ± 0·95 | 3·12 ± 1·40 | <0·001 | NA | NA | NA | NA |

| | | | | | | | | | | | | | | |
|---|---|---|---|---|---|---|---|---|---|---|---|---|---|---|
| Coopert et al., 2024 | 36 | IUD-LNG /DMPA | 158 | 7·8 (1·4) | 7·0 (1·7) | NA | 180 | 6·4 (2·0) | 5·3 (2·3) | NA | 150 | 6·4 (2·4) | 5·4 (3·0) | NA |
| | | COC continuous/cyclic | 152 | 7·9 (1·5) | 7·0 (2·1) | NA | 175 | 5·8 (2·1) | 5·4 (2·5) | NA | 159 | 6·4 (2·6) | 5·6 (2·8) | NA |
| Wei et al., 2024 | 12 | ENG | 50 | 7·0 (6·0–7·0) | 2·0 (1·0–2·5) | <0·01 | NA | NA | NA | NA | NA | NA | NA | NA |
| | | LNG-IUD | 58 | 7·0 (5·0–7·0) | 2·0 (1·0–2·0) | <0·01 | NA | NA | NA | NA | NA | NA | NA | NA |
| Gurbuz et al, 2025 | 12 | DNG | 40 | 7·23 ± 2·87 | 0·00 ± 0·00 | <0·05 | 40 | 4·18 ± 3·15 | 0·00 ± 0·00 | <0·05 | 40 | 3·90 ± 3·55 | 0·33 ± 1·41 | <0·05 |
| | | NETA | 30 | 7·43 ± 2·92 | 0·00 ± 0·00 | <0·05 | 30 | 4·97 ± 3·80 | 1·06 ± 2·43 | <0·05 | 30 | 4·00 ± 4·04 | 0·38 ± 1·50 | <0·05 |
| Tanha et al, 2025 | 3 | DNG | 52 | 7·98 ± 2·36 | 3·65 ± 2·46 | <0·001 | NA | NA | NA | NA | 52 | 5·5 (3·25–8·0) | 2·0 (0·0–4·0) | <0·001 |
| | | GnRH-a | 52 | 8·12 ± 1·92 | 3·44 ± 2·46 | <0·001 | NA | NA | NA | NA | 52 | 5·0 (3·0–7·0) | 0·0 (0·0–4·0) | <0·001 |
| **Modified Biberoglu & Behrmanf grading scale** | | | | | | | | | | | | | | |
| Vercellini et al., 1993 | 6 | cyclic OC | NA | NA | NA | NA | NA | NA | NA | NA | 21 | 1·8 ± 1·1 | 1·2 ± 0·7 | <0·01 |
| | | GnRH-a | NA | NA | NA | NA | NA | NA | NA | NA | 22 | 1·7 ± 0·9 | 1·1 ± 1·0 | <0·01 |
| Vercellini et al., 2002 | 6 | COC continuous | 36 | 2 (1-2) | 0 (0-0) | ns | 20 | 1 (0-2) | 0 (0-1) | ns | 25 | 1 (0-2) | 0 (0-1) | ns |
| | | CPA | 39 | 2 (1-2) | 0 (0-0) | ns | 22 | 1 (1-1) | 0 (0-1) | ns | 23 | 1 (0-1) | 0 (0-1) | ns |
| Vercellini et al., 2005 | 12 | COC continuous | 34 | 2·4 ± 0·6 | 0·3 ± 0·7 | <0·05 | 18 | 1·8 ± 0·7 | 0·8 ± 0·9 | <0·05 | 23 | 1·6 ± 0·7 | 0·4 ± 0·8 | <0·05 |
| | | NETA | 37 | 2·5 ± 0·6 | 0·1 ± 0·4 | <0·05 | 20 | 1·8 ± 0·7 | 0·4 ± 0·6 | <0·05 | 25 | 1·7 ± 0·8 | 0·5 ± 0·8 | <0·05 |
| Crosignani et al., 2006‖ | 6 | DMPA | NA | NA | NA | NA | 153 | 9·3 ± 2·4 | 3 ± 2·4 | <0·001 | NA | NA | NA | NA |
| | | GnRH-a | NA | NA | NA | NA | 146 | 9·8 ± 1·8 | 2·5 ± 1·8 | <0·001 | NA | NA | NA | NA |
| Schlaff et al., 2006‖ | 6 | DMPA | NA | NA | NA | NA | 136 | 10·0 ± 1·9 | 3·8 ± 1·9 | <0·001 | NA | NA | NA | NA |
| | | GnRH-a | NA | NA | NA | NA | 138 | 10·3 ± 1·9 | 2·6 ± 1·9 | <0·001 | NA | NA | NA | NA |
| Guzick et al., 2011** | 12 | COC continuous | NA | NA | NA | NA | 20 | 3·57 ±1·47 | [-1·4 ± (0·5)] | <0·01 | NA | NA | NA | NA |
| | | GNRH-a +add back | NA | NA | NA | NA | 20 | 4·35 ± 1·84 | [-2·71 ± (0·55)] | <0·001 | NA | NA | NA | NA |
| Osuga et al., 2021a | 6 | GnRH-ant | 103 | 1·2 ± 0·5†† | [−1·1 (±0·50)] | NA | 103 | 0·65 ± 0·44†† | [−0·4 (±0·45)] | NA | 44 | 0·55 ± 0·48†† | [−0·1 (±0·43)] | NA |
| | | GnRH-a | 81 | 1·2 ± 0·5†† | [−1·2 (±0·47)] | NA | 81 | 0·68 ± 0·55†† | [−0·5 (±0·49)] | NA | 26 | 0·60 ± 0·45†† | [−0·2 (±0·56)] | NA |

| | | | | | | | | | | | | | | |
|---|---|---|---|---|---|---|---|---|---|---|---|---|---|---|
| **Modified version Andersch and Milsom** | | | | | | | | | | | | | | |
| Vercellini et al., 1993 | 6 | cyclic OC | 24 | 5·0 ± 1·1 | 2·4 ± 1·7 | <0·01 | 24 | 2·9 ± 2·1 | 1·6 ± 1·9 | <0·01 | NA | NA | NA | NA |
| | | GnRH-a | 26 | 5·1 ± 1·6 | NA | | 26 | 3·0 ± 1·9 | 1·2 ± 1·3 | <0·01 | NA | NA | NA | NA |
| Parazzini et al., 2000 | 12 | cyclic OC | 47 | 4 (0-6) | 2 (0-5) | ns | 44 | 3 (0-5) | 0 (0-4) | ns | NA | NA | NA | NA |
| | | GnRH-a/ cyclic OC | 55 | 3 (0-6) | 0 (0-5) | ns | 55 | 2 (0-5) | 0 (0-5) | ns | NA | NA | NA | NA |

All pain scores reported as mean ± SD or median (IQR). Post-treatment values can also be expressed as a reduction from baseline (Petta et al., 2005, Guzick et al., 2011, Osuga et al., 2021a). P values are reported as provided in the original studies; 'ns' indicates non-significant results.

*Authors considered together severest endometriosis associated pelvic pain, dysmenorrhea during bleeding, dyspareunia and dyschezia. "Continuous COC" as it involves a non-stop regimen for 120 days followed by a 4-day pause (the only pause in a treatment course lasting a total of 6 months)
†Reported by authors as "global pain" (independent of the menstrual cycle or intercourse)
‡Reported by authors as "pain and/or dysmenorrhea in their last cycle"
§Reported by authors as "endometriosis-associated pelvic pain"
¶Reported by authors as combined pelvic pain (dysmenorrhea/chronic pelvic pain)
||Total score here is 15 considering together 5 domains (dysmenorrhea, dyspareunia, pelvic pain, pelvic tenderness and pelvic induration)
**Total score 0–9 scale, with 0–3 points assigned for each type of pain (dysmenorrhea, dyspareunia, non-cyclic pelvic pain)
††Baseline value taken from the first articles published by the same group (Osuga et al., 2021b)

NRS=numerical rating scale. VAS=visual analog scale. TX=treatment. COC=combined oral contraceptive. CPA=cyproterone acetate. DNG=dienogest. LNG-IUD=levonorgestrel-intrauterine device. GnRH-a=gonadotropin resealing hormone agonist. GnRH-ant=gonadotropin resealing hormone antagonist. DMPA=depot medroxyprogesterone acetate. SD=standard deviation. IQR=interquartile range. NA=not applicable.

### Table S2 Bleeding patterns during hormonal therapies for endometriosis

| Study | TX length (months) | Intervention | *n* | Spotting | Breakthrough bleeding | Amenorrhoea |
|---|---|---|---|---|---|---|
| Vercellini et al., 1993 | 6 | cyclic COC | 28 | 7 | NA | NA |
| | | GnRH-a | 29 | 6 | NA | 29 |
| Cosson et al., 2002 | 12 | DNG | 59 | 61·6% | 1·6% | NA |
| | | GnRH-a | 61 | 25·4% | NA | NA |
| Vercellini et al., 2002 | 6 | continuous COC | 41 | 18 | 4 | 19 |
| | | CPA | 43 | 12 | 3 | 28 |
| Zupi et al., 2004 | 12 | GnRH-a + add back | 46 | 3 (6·5%) | NA | NA |
| | | GnRH-a | 44 | 1 (2·3%) | NA | NA |

| | | | | | | |
|---|---|---|---|---|---|---|
| | | continuous COC | 43 | 7 (16·2%) | NA | NA |
| Petta et al., 2005 | 6 | LNG-IUD | 39 | NA | NA | 70% |
| | | GnRH-a | 43 | NA | NA | 98% |
| Vercellini et al., 2005 | 12 | continuous COC | 38 | 14 | 7 | 17 (45%) |
| | | NETA | 40 | 9 | 2 | 29 (72%) |
| Crosignani et al., 2006 | 6 | DMPA | 153 | NA | 19 (12·5%) | 24% |
| | | GnRH-a | 146 | NA | 1 (0·7%) | 89·9% |
| Schlaff et al., 2006 | 6 | DMPA | 130 | NA | 7 (5·4%) | NA |
| | | GnRH-a | 135 | NA | 1 (0·7%) | NA |
| Razzi et al., 2007 | 6 | desogestrel | 20 | NA | 4 | NA |
| | | continuous COC | 20 | NA | 0 | NA |
| Sesti et al., 2007 | 6 | GnRH-a | 39 | NA | NA | 39 |
| | | continuous COC | 38 | NA | NA | NA |
| Walch et al., 2009 | 12 | ENG | 21* | NA | NA | 3 (14%) |
| | | DMPA | 20* | NA | NA | 3 (15%) |
| Tekin et al., 2011 | 6 | LNG-IUD | 20 | NA | 13 (65%) | 0 |
| | | GnRH-a | 20 | NA | 0 | 6 (30%) |
| Strowitzki et al., 2010 | 6 | DNG | 120 | 16·7% | NA | 38·9% |
| | | GnRH-a | 128 | NA | NA | 75·9% |
| Wong et al., 2010 | 36 | LNG-IUD | 15 | 1 | 0 | NA |
| | | DMPA | 15 | 2 | 1 | NA |
| Muzii et al., 2011 | 6 | continuous COC | 29 | NA | 10 | NA |
| | | cyclic COC | 28 | NA | 2 | NA |
| Cheewadhanarak et al., 2012 | 6 | DMPA | 39 | 28 (71·8%) | 4 (10·3%) | 7 (17·9%) |
| | | continuous COC | 38 | 24 (63·2%) | 11 (28·9%) | 3 (7·9%) |

| | | | | | | |
|---|---|---|---|---|---|---|
| Cucinella et al., 2013 | 24 | continuous COC (desogestrel) | 43 | 25% | NA | NA |
| | | continuous COC (gestodene) | 44 | 32% | NA | NA |
| | | cyclic COC (DNG) | 43 | 15% | NA | NA |
| Carr et al., 2014 | 6 | GnRH-ant | 84 | NA | 0 | NA |
| | | GnRH-ant | 84 | NA | 0 | NA |
| | | DMPA | 83 | NA | 7 | NA |
| Granese et al., 2015 | 9 | continuous COC | 39 | 2 | NA | NA |
| | | GnRH-a | 39 | 0 | NA | NA |
| Takaesu et al., 2016 | 6 | DNG | 54 | 54 (100%) | NA | NA |
| | | GnRH-a | 51 | 3 (6%) | NA | NA |
| Abdou et al., 2018 | 3 | DNG | 121 | NA | 78 (64·5%) | NA |
| | | GnRH-a | 121 | NA | 26 (21·5%) | NA |
| Carvalho et al., 2018 | 6 | ENG | 45 | 15% | 24·4% | 28·8% |
| | | LNG-IUD | 40 | 22·1% | 30·0% | 10% |
| Ceccaroni et al., 2021 | 6 | GnRH-a | 81 | 1 (1·2%) | NA | 77 (95%) |
| | | DNG | 65 | 18 (27·7%) | NA | 52 (80%) |
| El Taha et al., 2021 | 6 | DNG | 35 | 43% | NA | 19% |
| | | continuous COC | 35 | 30% | NA | 14% |
| Hassanin et al., 2021 | 6 | DNG | 48 | 33 (68·8%) | NA | 5 (10·4%) |
| | | cyclic COC | 49 | 2 (4·1%) | NA | NA |
| Mehdizadeh Kashi et al., 2022 | 6 | DNG | 30 | 6 (20%) | NA | 24 (80%) |
| | | continuous COC | 30 | 4 (13·3%) | NA | 26 (86·7%) |
| Osuga et al., 2021a | 6 | GnRH-ant | 103 | NA | 30 (29·1%) | NA |
| | | GnRH-a | 80 | NA | 32 (40·0%) | NA |
| Ota et al., 2021 | 72 | LNG-IUD | 76 | NA | 6 | NA |

| | | | | | | |
|---|---|---|---|---|---|---|
| | | DNG | 81 | NA | 6 | NA |
| Guo et al., 2023 | 36 | LNG-IUD | 48 | NA | 33 (68·8%) | NA |
| | | DNG | 69 | NA | 41 (59·4%) | NA |
| Vahid Dastjerdi et al., 2023 | 6 | DNG | 48 | NA | 12 (25%) | NA |
| | | oral MPA | 53 | NA | 11 (20·7%) | NA |
| Amiya et al., 2024 | 3 | DNG | 30 | NA | 2 (6·7%) | NA |
| | | oral MPA | 30 | NA | 3 (10%) | NA |
| Choudhury et al., 2024 | 3 | LNG-IUD | 34 | 8 (23·5%) | NA | 7 (20·5%) |
| | | DNG | 34 | 13 (38·2%) | NA | 9 (26·4%) |
| Cooper et al., 2024 | 36 | IUD-LNG/DMPA | 101 | NA | 51 (50%) | 50 (50%) |
| | | continuous/cyclic COC | 98 | NA | 62 (63%) | 36 (37%) |
| Wei et al., 2024 | 12 | ENG | 50 | 7 (14%) | NA | 43 (86%)† |
| | | LNG-IUD | 58 | 3 (5·2%) | NA | 55 (94·8%)† |
| De La Hoz et al., 2025 | 6 | DRSP | 94 | 14 (14·9%) | NA | 23 (24·5%) |
| | | cyclic COC | 91 | 16 (15·3%) | NA | 19 (20·9%) |
| Tanha et al., 2025 | 3 | DNG | 52 | 6 (11·5%) | NA | NA |
| | | GnRH-a | 52 | 4 (7·7%) | NA | NA |

Data are reported as number of patients (N), percentage (%), or both (N, %), as provided in the original studies.

*Eighteen patients in the ENG group and 17 in the DMPA group experienced some form of vaginal bleeding or spotting during the study.
†Reported as "ideal vaginal bleeding" (less than 5 episodes of bleeding or dripping within 90 days).

NA=not applicable. COC=combined oral contraceptive. CPA=cyproterone acetate. NETA=norethisterone acetate. DNG=dienogest. LNG-IUD=levonorgestrel-intrauterine device. GnRH-a=gonadotropin resealing hormone agonist. GnRH-ant=gonadotropin resealing hormone antagonist. (D)MPA=(depot) medroxyprogesterone acetate. ENG=etonogestrel. DRSP=drospirenone.

### Table S3 Adverse events across hormonal therapies for endometriosis

| Study | Intervention | *n* | Mood changes | Decreased libido | Weight gain | Headache | Abdominal discomfort/ bloating | Breast tenderness |
|---|---|---|---|---|---|---|---|---|
| Vercellini et al., 1993 | cyclic COC | 28 | 5 | 4 | 4 | 6 | NA | 5 |

| | | | | | | | | |
|---|---|---|---|---|---|---|---|---|
| | GnRH-a | 29 | 5 | 6 | 2 | 4 | NA | 3 |
| Cosson et al., 2002 | DNG | 64 | NA | NA | 0 | 1 (1·6%) | 1 (1·6%) | NA |
| | GnRH-a | 62 | NA | NA | 0 | 1 (1·6%) | NA | NA |
| Vercellini et al., 2002 | continuous COC | 41 | 3* | 2 | 10 | 7 | 15 | NA |
| | CPA | 43 | 8* | 7 | 8 | 2 | 14 | NA |
| Zupi et al., 2004 | GnRH-a + add back | 46 | 5 (10·8%) | NA | NA | NA | NA | NA |
| | GnRH-a | 44 | 16 (36·4%) | NA | NA | NA | NA | NA |
| | continuous COC | 43 | 3 (6·9%) | NA | NA | NA | NA | NA |
| Vercellini et al., 2005 | continuous COC | 45 | 2† | 2 | 7 | 3 | 1 | 1 |
| | NETA | 45 | 3† | 4 | 12 | 2 | 4 | 0 |
| Crosignani et al., 2006 | DMPA | 153 | NA | NA | NA | 5 (3·3%) | NA | 8 (5·3%) |
| | GnRH-a | 146 | NA | NA | NA | 9 (6·3%) | NA | 5 (3·5%) |
| Schlaff et al., 2006 | DMPA | 130 | NA | 3 (2·3%) | NA | 10 (7·7%) | NA | NA |
| | GNRH-a | 135 | NA | 7 (5·2%) | NA | 14 (10·4%) | NA | NA |
| Razzi et al., 2007 | desogestrel | 20 | NA | NA | 0 | NA | NA | NA |
| | continuous COC | 20 | NA | NA | 3 | NA | NA | NA |
| Walch et al., 2009 | ENG | 21 | 0† | 5 (24%) | NA | 3 (14%) | NA | 5 (24%) |
| | DMPA | 20 | 2 (10%)† | 6 (30%) | NA | 4 (20%) | NA | 3 (15%) |
| Strowitzki et al., 2010 | DNG | 120 | 6 (5%)† | 5 (4·2%) | 8 (6·7%) | 15 (12·5%) | NA | NA |
| | GnRH-a | 128 | 11 (8·6%)† | 8 (6·3%) | 5 (3·9%) | 25 (19·5%) | NA | NA |
| Muzii et al., 2011 | continuous COC | 29 | NA | NA | NA | 2 | NA | NA |
| | cyclic COC | 28 | NA | NA | NA | 2 | NA | NA |
| Tekin et al., 2011 | LNG-IUD | 20 | NA | NA | 2 (10%) | NA | NA | NA |
| | GnRH-a | 20 | NA | NA | 1 (5%) | NA | NA | NA |
| Cheewadhanarak et al., 2012 | DMPA | 39 | 12 (30·8%)‡ | NA | NA | NA | NA | NA |
| | continuous COC | 42 | NA | NA | NA | NA | NA | 21 (50%) |
| Carr et al., 2014 | GnRH-ant | 84 | 7 (8·3%) | NA | NA | 22 (26·2%) | 2 (2·4%) | NA |
| | GnRH-ant | 84 | 6 (7·1%) | NA | NA | 23 (27·4%) | 0 | NA |

| | | | | | | | | |
|---|---|---|---|---|---|---|---|---|
| | DMPA | 83 | 10 (11·9%) | NA | NA | 15 (17·9%) | 6 (7·1%) | NA |
| Granese et al., 2015 | continuous COC | 39 | NA | 12 | 2 | 7 | NA | NA |
| | GnRH-a | 39 | NA | 4 | 1 | 1 | NA | NA |
| Takaesu et al., 2016 | DNG | 54 | NA | NA | NA | 5 (9%) | NA | NA |
| | GNRH-a | 51 | NA | NA | NA | 2 (4%) | NA | NA |
| Harada et al., 2017 | continuous COC | 150 | NA | NA | NA | 22 (16·9%) | 3 (2·3%) | 3 (2·3%) |
| | DNG | 53 | NA | NA | NA | NA | NA | NA |
| Abdou et al., 2018 | DNG | 121 | NA | NA | 13 (10·8%) | 17 (14%) | NA | NA |
| | GnRH-a | 121 | NA | NA | 4 (3·3%) | 26 (21·5%) | NA | NA |
| Ozaki et al., 2020 | DNG | 35 | NA | 10 | 6 | NA | NA | NA |
| | GnRH-a | 35 | NA | 10 | 10 | NA | NA | NA |
| Ceccaroni et al., 2021 | GnRH-a | 81 | 35 (43%) | 10 (16%) | NA | 12 (14·8%) | 20 (24·7%) | 2 (2·4%) |
| | DNG | 65 | 23 (35·4%) | 7 (10·8%) | NA | 15 (23%) | 21 (32%) | 5 (7·7%) |
| El Taha et al., 2021 | DNG | 31 | 14 (45·2%) | 1 (3·2%) | 3 (9·7%) | 10 (32·3%) | 5 (16·1%) | 6 (19·4%) |
| | continuous COC | 32 | 24 (75%) | 3 (9·4%) | 11 (34·4%) | 19 (59·4%) | 12 (37·5%) | 15 (46·9%) |
| Hassanin et al., 2021 | DNG | 48 | NA | NA | 8 (16·7%) | 6 (12·5%) | NA | 8 (16·7%) |
| | cyclic COC | 49 | NA | NA | 5 (10.2%) | 7 (14·3%) | NA | 4 (8·2%) |
| Khalifa et al., 2021 | GnRH-a | 67 | 8§ | NA | NA | 0 | NA | 0 |
| | DNG | 67 | 0 | NA | NA | 13 | NA | 7 |
| Osuga et al., 2021a | GnRH-ant | 103 | NA | NA | NA | 11 (10·7%) | NA | NA |
| | GnRH-a | 80 | NA | NA | NA | 11 (13·8%) | NA | NA |
| Ota et al., 2021 | LNG-IUD | 76 | NA | 0 | NA | NA | 3¶ | NA |
| | DNG | 81 | NA | 4 | NA | NA | 4¶ | NA |
| Guo et al., 2023 | LNG-IUD | 48 | 5 (10·4%)† | NA | 18 (37·5%) | NA | NA | 6 (12·5%) |
| | DNG | 69 | 6 (8·7%)† | NA | 29 (42%) | NA | NA | 9 (13·0%) |
| Vahid Dastjerdi et al., 2023 | DNG | 48 | 7 (14·6%) | NA | 5 (10·6%) | 21 (43·7%) | NA | 1 (2·1%) |
| | oral MPA | 53 | 13 (25%) | NA | 20 (37.7%) | 12 (22·7%) | NA | 4 (7·6%) |
| Amiya et al., 2024 | DNG | 30 | 6 (20%)‖ | 1 (4%) | 9 (30%) | 5 (16·7%) | 2 (6.7%) | 4 (13·3%) |
| | oral MPA | 30 | 6 (20%)‖ | 7 (25%) | 11 (36·7%) | 1 (3·3%) | 7 (23.3%) | 8 (26.7%) |
| Choudhury et al., 2024 | LNG-IUD | 34 | NA | NA | NA | NA | NA | 3 (8·8%) |
| | DNG | 34 | NA | NA | NA | NA | NA | 4 (11·8%) |

| | | | | | | | | |
|---|---|---|---|---|---|---|---|---|
| Wei et al., 2024 | ENG | 50 | NA | NA | 16 (32%) | NA | NA | NA |
| | LNG-IUD | 58 | NA | NA | 5 (9·1%) | NA | NA | NA |
| De La Hoz et al., 2025 | DRSP | 94 | 3 (3·2%) | NA | 4 (4·3%) | 7 (7·4%) | NA | 11 (11·7%) |
| | cyclic COC | 91 | 4 (4·4%) | NA | 6 (6·6%) | 8 (8·8%) | NA | 15 (16·5%) |
| Tanha et al., 2025 | DNG | 52 | NA | 24 (46·2%) | NA | NA | NA | NA |
| | GNRH-a | 52 | NA | 12 (23·1%) | NA | NA | NA | NA |

Data are reported as number of patients (N), percentage (%), or both (N, %), as provided in the original studies.

*Depression and irritability
†Depression
‡Irritability
§Low mood
¶Reported as severe irritable bowel syndrome symptoms
||Depression and aggression

NA=not applicable. COC=combined oral contraceptive. CPA=cyproterone acetate. NETA=norethisterone acetate. DNG=dienogest. LNG-IUD=levonorgestrel-intrauterine device. GnRH-a=gonadotropin resealing hormone agonist. GnRH-ant=gonadotropin resealing hormone antagonist. (D)MPA=(depot) medroxyprogesterone acetate. ENG=etonogestrel. DRSP=drospirenone.

### Table S4 Adverse events across hormonal therapies for endometriosis

| Study | Intervention | *n* | Hot flushes/ night sweats | Vaginal dryness | Nausea | Hair loss | Acne/ oily skin | Insomnia |
|---|---|---|---|---|---|---|---|---|
| Vercellini et al., 1993 | cyclic COC | 28 | 1 | 0 | NA | NA | NA | 0 |
| | GnRH-a | 29 | 24 | 5 | NA | NA | NA | 7 |
| Cosson et al., 2002 | DNG | 64 | 9·6% | NA | NA | 0 | 0 | NA |
| | GnRH-a | 62 | 61·2% | NA | NA | 0 | 0 | NA |
| Vercellini et al., 2002 | continuous COC | 41 | 0 | 0 | 4 | NA | NA | NA |
| | CPA | 43 | 3 | 2 | 0 | NA | NA | NA |
| Zupi et al., 2004 | GnRH-a + add back | 46 | 12 (26·1%) | NA | NA | NA | NA | NA |
| | GnRH-a | 44 | 34 (77·3%) | NA | NA | NA | NA | NA |
| | continuous COC | 43 | 0 | NA | NA | NA | NA | NA |
| Vercellini et al., 2005 | continuous COC | 45 | NA | NA | 3 | NA | 1 | NA |

| | | | | | | | | |
|---|---|---|---|---|---|---|---|---|
| | NETA | 45 | NA | NA | 0 | NA | 2 | NA |
| Crosignani et al., 2006 | DMPA | 153 | 9 (5·9%) | NA | 17 (11·2%) | NA | NA | NA |
| | GnRH-a | 146 | 24 (16·8%) | NA | 10 (7·0%) | NA | NA | NA |
| Schlaff et al., 2006 | DMPA | 130 | 3 (2·3%) | 0 | NA | NA | NA | 3 (2·3%) |
| | GNRH-a | 135 | 15 (11·1%) | 5 (3·7%) | NA | NA | NA | 7 (5·2%) |
| Razzi et al., 2007 | desogestrel | 20 | NA | NA | NA | NA | NA | NA |
| | continuous COC | 20 | NA | NA | NA | NA | NA | NA |
| Walch et al., 2009 | ENG | 21 | 1 (5%) | NA | NA | 1 (5%) | 0 | NA |
| | DMPA | 20 | 2 (10%) | NA | NA | 2 (10%) | 1 (5%) | NA |
| Strowitzki et al., 2010 | DNG | 120 | 0 | 2 (1·7%) | NA | 4 (3·3%) | 5 (4·1%) | 2 (1·7%) |
| | GnRH-a | 128 | 9 (7%) | 9 (7%) | NA | 7 (5·5%) | 6 (4·7%) | 10 (7·8%) |
| Muzii et al., 2011 | continuous COC | 29 | NA | NA | NA | NA | NA | NA |
| | cyclic COC | 28 | NA | NA | NA | NA | NA | NA |
| Tekin et al., 2011 | LNG-IUD | 20 | 0 | NA | NA | NA | NA | NA |
| | GnRH-a | 20 | 10 (50%) | NA | NA | NA | NA | NA |
| Cheewadhanarak et al., 2012 | DMPA | 39 | NA | NA | NA | NA | 15 (38·5%) | NA |
| | continuous COC | 42 | NA | NA | 15 (25·7%) | NA | NA | NA |
| Carr et al., 2014 | GnRH-ant | 84 | 60 (71%) | NA | 16 (19%) | NA | 7 (8·3%) | 4 (4·8%) |
| | GnRH-ant | 84 | 69 (82%) | NA | 13 (15·5%) | NA | 2 (2·4%) | 7 (8·3%) |
| | DMPA | 83 | 63 (76%) | NA | 13 (15.5%) | NA | 7 (8·3%) | 4 (4·8%) |
| Granese et al., 2015 | continuous COC | 39 | 0 | 8 | NA | NA | NA | NA |
| | GnRH-a | 39 | 1 | 1 | NA | NA | NA | NA |
| Takaesu et al., 2016 | DNG | 54 | 6 (11%) | NA | NA | 0 | NA | NA |
| | GNRH-a | 51 | 48 (94%) | NA | NA | 1 (2%) | NA | NA |
| Harada et al., 2017 | continuous COC | 150 | NA | NA | 15 (11·5%) | NA | NA | NA |
| | DNG | 53 | NA | NA | NA | NA | NA | NA |
| Abdou et al., 2018 | DNG | 121 | 19 (15·7%) | 4 (3·3%) | NA | NA | NA | NA |
| | GnRH-a | 121 | 56 (46·3%) | 19 (15·7%) | NA | NA | NA | NA |
| Ozaki et al., 2020 | DNG | 35 | NA | 3 | NA | 5 | 5 | NA |

| | | | | | | | | |
|---|---|---|---|---|---|---|---|---|
| | GnRH-a | 35 | NA | 6 | NA | 7 | 6 | NA |
| Ceccaroni et al., 2021 | GnRH-a | 81 | 70 (86·4%) | 13 (16%) | NA | 17 (21%) | NA | NA |
| | DNG | 65 | 8 (12%) | 13 (20%) | NA | 10 (15%) | NA | NA |
| El Taha et al., 2021 | DNG | 31 | NA | NA | 5 (16·1%) | NA | NA | 3 (9·7%) |
| | continuous COC | 32 | NA | NA | 17 (53·1%) | NA | NA | 9 (28·1%) |
| Hassanin et al., 2021 | DNG | 48 | 11 (22·9%) | NA | 8 (16·7%) | NA | NA | NA |
| | cyclic COC | 49 | 0 | NA | 2 (4·1%) | NA | NA | NA |
| Khalifa et al., 2021 | GnRH-a | 67 | 11 | 7 | NA | NA | NA | 8 |
| | DNG | 67 | 0 | 0 | NA | NA | NA | 0 |
| Osuga et al., 2021a | GnRH-ant | 103 | 55 (53·4%) | NA | NA | NA | NA | NA |
| | GnRH-a | 80 | 37 (46·3%) | NA | NA | NA | NA | NA |
| Ota et al., 2021 | LNG-IUD | 76 | NA | NA | NA | NA | NA | NA |
| | DNG | 81 | NA | NA | NA | NA | NA | NA |
| Guo et al., 2023 | LNG-IUD | 48 | NA | NA | NA | NA | NA | 5 (10·4%) |
| | DNG | 69 | NA | NA | NA | NA | NA | 7 (10·1%) |
| Vahid Dastjerdi et al., 2023 | DNG | 48 | NA | NA | NA | 8 (16·6%) | NA | NA |
| | oral MPA | 53 | NA | NA | NA | 0 | NA | NA |
| Amiya et al., 2024 | DNG | 30 | 0 | 1 (3·3%) | 1 (3·3%) | NA | 1 (3·3%) | 0 |
| | oral MPA | 30 | 4 (13·3%) | 8 (26·7%) | 3 (10%) | NA | 4 (13·3%) | 0 |
| Choudhury et al., 2024 | LNG-IUD | 34 | 0 | NA | NA | NA | NA | NA |
| | DNG | 34 | 3 (8·8%) | NA | NA | NA | NA | NA |
| Wei et al., 2024 | ENG | 50 | NA | NA | NA | NA | 10 (18%) | NA |
| | LNG-IUD | 58 | NA | NA | NA | NA | 5 (8·6%) | NA |
| De La Hoz et al., 2025 | DRSP | 94 | NA | NA | NA | 5 (5·3%) | 10 (10·6%) | NA |
| | cyclic COC | 91 | NA | NA | NA | 3 (3·3%) | 8 (8·8%) | NA |
| Tanha et al., 2025 | DNG | 52 | 16 (30·8%) | 13 (25%) | NA | NA | NA | NA |
| | GNRH-a | 52 | 18 (34·6%) | 24 (46·2%) | NA | NA | NA | NA |

Data are reported as number of patients (N), percentage (%), or both (N, %), as provided in the original studies.

NA=not applicable. COC=combined oral contraceptive. CPA=cyproterone acetate. NETA=norethisterone acetate. DNG=dienogest. LNG-IUD=levonorgestrel-intrauterine device. GnRH-a=gonadotropin resealing hormone agonist. GnRH-ant=gonadotropin resealing hormone antagonist. (D)MPA=(depot) medroxyprogesterone acetate. ENG=etonogestrel. DRSP=drospirenone.

**Table S5 Treatment discontinuation for any reasons across hormonal therapies for endometriosis**

| | | | Drop out reasons | | | | |
|---|---|---|---|---|---|---|---|
| **Study** | **Intervention** | ***n*** | **Adverse events related to treatment** | **Treatment inefficacy/ failure** | **Lost to follow up** | **Other (withdrawal of consent, protocol deviation)** | **Specify adverse events** |
| Vercellini et al., 1993 | cyclic COC | 28 | 1 | NA | 1 | NA | Severe headache (1) |
| | GNRH-a | 29 | 0 | NA | 1 | NA | NA |
| Parazzini et al., 2000 | cyclic COC | 47 | NA | NA | NA | 2 | NA |
| | GnRH-a/cyclic COC | 55 | NA | NA | NA | 1 | NA |
| Cosson et al., 2002 | DNG | 64 | 4 | NA | NA | 1 | Diabetes (1), metrorrhagia (1), migraine (1), gastric pain (1) |
| | GnRH-a | 62 | 1 | NA | NA | NA | Headache and hot flushes (1) |
| Vercellini et al., 2002 | continuous COC | 45 | 5 | 2 | 2 | NA | Weight gain (2), headache (1), and nausea (1) |
| | CPA | 45 | 4 | 2 | 0 | NA | Abdominal bloating (1), decreased libido (1), depression (1), headache (1) |
| Petta et al., 2005 | LNG-IUD | 39 | 0 | NA | NA | 5 | NA |
| | GnRH-a | 43 | 0 | NA | NA | 6 | NA |
| Vercellini et al., 2005 | continuous COC | 45 | 3 | 3 | 1 | NA | Depression (1), headache (1), decreased libido (1) |
| | NETA | 45 | 2 | 3 | 0 | NA | Erythematous reaction (1), decreased libido (1) |
| Crosignani et al., 2006 | DMPA | 153 | 3 | NA | 1 | 11 | Not specified |
| | GnRH-a | 146 | 2 | NA | 3 | 5 | Not specified |
| Schlaff et al., 2006 | DMPA | 136 | 9 | 7 | 14 | 18 | Not specified (cited only blending/spotting) |
| | GnRH-a | 138 | 9 | 1 | 11 | 15 | |
| Sesti et al., 2009 | GnRH-a | 65 | 7 | NA | NA | NA | hot flushes, vaginal dryness, reduced libido |
| | continuous COC | 64 | 4 | NA | NA | NA | Not specified (cited only breakthrough bleeding, headache, breast tension, nausea, and weight gain) |
| Walch et al., 2009 | ENG | 21 | 3 | 1 | 0 | NA | Recurrent hot flushes (1), unbearable spotting-bleeding episodes (2) |
| | DMPA | 20 | 4 | 1 | 1 | 1 | Severe depression (1), hot flushes (1), weight gain (1), loss of hair combined with weight gain (1) |
| Seracchioli et al., 2010b | cyclic COC | 103 | 8 | NA | NA | 3 | Not specified |
| | continuous COC | 104 | 6 | NA | NA | 3 | Not specified |
| Strowitzki et al., 2010 | DNA | 120 | 6 | NA | NA | 4 | Hypertension (1), tinnitus (1), ovarian cyst (1), nausea (1), depression (2) |
| | GnRH-a | 128 | 5 | NA | NA | 3 | Hot flushes (1), arthritis (2), depression (1), allergic reaction (1), sleep disorder (1) |
| Wong et al., 2010 | LNG-IUD | 15 | 1 | NA | 0 | 1 | Prolonged vaginal spotting (1) |
| | depot MPA | 15 | 5 | NA | 0 | 3 | Prolonged vaginal spotting (2), increase pain and heavy bleeding (1), significant bone loss (2) |
| Muzii et al., 2011 | continuous COC | 29 | 12 | NA | NA | NA | Bleeding related (10), headache (2) |
| | cyclic COC | 28 | 4 | NA | NA | NA | Bleeding related (2), headache (2) |
| Cheewadhanarak et al., 2012 | depot MPA | 42 | 2 | 1 | 0 | 0 | Weight gain (2) |
| | continuous COC | 42 | 1 | 3 | 0 | 0 | Bleeding (1) |

| | | | | | | | |
|---|---|---|---|---|---|---|---|
| Cucinella et al., 2013 | continuous COC (desogestrel) | 43 | 11 | NA | NA | 14 | Not specified (reported only headache, decreased libido, spotting, water retention, vaginal dryness, depression, acne and insomnia) |
| | continuous COC (gestodene) | 44 | | | | | |
| | cyclic COC (DNG) | 43 | | | | | |
| Carr et al., 2014 | GnRH-ant | 84 | 4 | 4 | 5 | 15 | Not specified (reported only BP increase, dehydration, headache, mood changes, dizziness, fall, hot flushes, weight decreased, bleeding, abdominal distension, nausea and scotoma) |
| | GnRH-ant | 84 | 7 | 2 | 4 | 9 | |
| | DMPA | 84 | 14 | 3 | 5 | 11 | |
| Granese et al., 2015 | continuous COC | 39 | 1 | NA | 3 | 2 | Not specified |
| | GnRH-a | 39 | 0 | NA | 5 | 5 | NA |
| Shaaban et al., 2015 | LNG-IUD | 31 | 1 | 1 | 0 | NA | Spontaneous expulsion (1) |
| | cyclic COC | 31 | 1 | 0 | 2 | NA | Not specified |
| Takaesu et al., 2016 | DNG | 54 | 1 | NA | 2 | 1 | Not specified |
| | GnRH-a | 51 | 3 | NA | 4 | 1 | Not specified |
| Harada et al., 2017 | continuous COC | 150 | 12 | NA | 0 | 14 | Not specified (cited only naso-pharyngitis, genital haemorrhage, and headache) |
| | DNG | 53 | 0 | NA | 0 | 8 | NA |
| Abdou et al., 2018 | DNG | 130 | 0 | NA | 9 | NA | NA |
| | GnRH-a | 131 | 0 | NA | 10 | NA | NA |
| Margatho et al., 2020 | ENG | 52 | 1 | 1* | 20 | 6 | Irritability and weight gain (1) |
| | LNG-IUD | 51 | 5 | 0* | 21 | 0 | Device expulsion (5) |
| El Taha et al., 2021 | DNG | 35 | 3 | 2 | 2 | 3 | Irritability and weight gain (2), irritability and spotting (1) |
| | continuous COC | 35 | 3 | 2 | 1 | 3 | Prolonged bleeding (1), irritability and weight gain (2) |
| Hassanin et al., 2021 | DNG | 55 | 0 | 3† | 4 | 1 | NA |
| | cyclic COC | 55 | 0 | 3† | 3 | 0 | NA |
| Osuga et al., 2021a | GnRH-ant | 103 | 2 | 0 | 0 | 1 | Not specified (most common reported were headache, bleeding and hot flushes) |
| | GnRH-a | 80 | 9 | 0 | 0 | 8 | |
| Ota et al., 2021 | LNG-IUD | 76 | 14 | 9 | NA | NA | Severe IBS symptoms (3), heavy uterine bleeding (6), expulsion of IUD (7) |
| | DNG | 81 | 14 | 0 | NA | NA | Decreased libido (4), severe IBS symptoms (4), heavy uterine bleeding (6) |
| Mehdizadeh Kashi et al., 2022 | DNG | 30 | 1 | NA | 5 | NA | Not specified (most common reported were spotting, hair loss, headache, hot flushes, and nausea) |
| | continuous COC | 30 | 1 | NA | 5 | NA | |
| Guo et al., 2023 | LNG-IUD | 48 | 8 | NA | NA | NA | Expulsion of IUD (8) |
| | DNG | 69 | 3 | NA | NA | NA | Excessive vaginal bleeding (3) |
| Cooper et al., 2024 | IUD-LNG/ DMPA | 205 | NA | 50‡ | NA | NA | NA |
| | continuous/ cyclic COC | 200 | NA | 61‡ | NA | NA | NA |
| da Costa Porto et al., 2024 | DNG | 20 | 1 | NA | NA | NA | Not specified |
| | LNG-IUD | 20 | 1 | NA | NA | NA | Expulsion of IUD (1) |
| Gurbuz et al., 2025 | DNG | 40 | 11 | 2 | 0 | 2 | Not specified, reported primarily AUB (66.7 %) and mood changes (8.3 %) |
| | NETA | 30 | 19 | 2 | 0 | 1 | |

Data are reported as number of patients (N), as provided in the original studies.

* In addition, five patients in the ENG group and two in the IUD group required surgery during treatment, but no reason was given. One other patient in the ENG group and four in the IUD group changed their treatment during the trial, switching to other hormonal therapies.

† Treatment failure included the initiation of alternative medical or surgical therapy, or pregnancy.

‡ The necessity for further surgery (laparoscopy to investigate recurrent pain, to treat endometriosis, or hysterectomy) or the use of GnRH analogues was used as an indicator of treatment failure.

NA=not applicable. COC=combined oral contraceptive. CPA=cyproterone acetate. NETA=norethisterone acetate. DNG=dienogest. LNG-IUD=levonorgestrel-intrauterine device. GnRH-a=gonadotropin resealing hormone agonist. GnRH-ant=gonadotropin resealing hormone antagonist. (D)MPA=(depot) medroxyprogesterone acetate. ENG=etonogestrel. DRSP=drospirenone. BP=blood pressure. IBS=irritable bowel syndrome. AUB=abnormal uterine bleeding**.**

**Table S6 'Absolute' RCT trustworthiness criteria**

| **Authors, year** | **Retraction** (the RCT has not been retracted) | **Registration** (the RCT was pre- registered in a publicly available international clinical trials registry prior to randomization) | **Ethics** (the RCT was approved by an ethics committee or an institutional review board) | **CONSORT** (statement that CONSORT guidance was followed) | **Outcome variation** (the primary outcome of the RCT is consistent with the stated primary outcome in the RCT registration) | **Does the article meet criteria?** |
|---|---|---|---|---|---|---|
| Vercellini et al., 1993 | Not retracted | NA | Yes | NA | NA | Yes* |
| Parazzini et al., 2000 | Not retracted | NA | Yes | NA | NA | Yes* |
| Cosson et al., 2002 | Not retracted | NA | Yes | NA | NA | Yes* |
| Vercellini et al., 2002 | Not retracted | NA | Yes | NA | NA | Yes* |
| Zupi et al., 2004 | Not retracted | NA | Yes | Yes | NA | Yes* |
| Petta et al., 2005 | Not retracted | NA | Yes | NA | NA | Yes* |
| Vercellini et al., 2005 | Not retracted | NA | Yes | NA | NA | Yes* |
| Crosignani et al., 2006 | Not retracted | NA | Yes | NA | NA | Yes* |
| Schlaff et al., 2006 | Not retracted | NA | Yes | NA | NA | Yes* |
| Razzi et al., 2007 | Not retracted | NA | Yes | NA | NA | Yes* |
| Sesti et al., 2007 | Not retracted | NA | Yes | Yes | NA | Yes* |
| Sesti et al., 2009 | Not retracted | NA | Yes | Yes | NA | Yes* |
| Walch et al., 2009 | Not retracted | NA | Yes | NA | NA | Yes* |

| | | | | | | |
|---|---|---|---|---|---|---|
| Seracchioli et al., 2010a | Not retracted | NA | Yes | NA | NA | Yes* |
| Seracchioli et al., 2010b | Not retracted | NA | Yes | NA | NA | Yes* |
| Strowitzki et al., 2010 | Not retracted | NA | Yes | NA | NA | Yes* |
| Wong et al., 2010 | Not retracted | NA | Yes | NA | NA | Yes* |
| Muzii et al., 2011 | Not retracted | NA | Yes | NA | NA | Yes* |
| Guzick et al., 2011 | Not retracted | NA | Yes | NA | NA | Yes* |
| Tekin et al., 2011 | Not retracted | NA | Yes | NA | NA | Yes* |
| Cheewadhanarak et al., 2012 | Not retracted | NA | Yes | NA | NA | Yes* |
| Cucinella et al., 2013 | Not retracted | NA | Yes | NA | NA | Yes* |
| Carr et al., 2014 | Not retracted | NA | No | NA | NA | No |
| Granese et al., 2015 | Not retracted | NA | Yes | Yes | NA | Yes* |
| Shaaban et al., 2015 | Not retracted | Yes | Yes | No | Yes | Yes, with concerns |
| Takaesu et al., 2016 | Not retracted | NA | Yes | NA | NA | Yes* |
| Harada et al., 2017 | Not retracted | Yes | Yes | No | Yes | Yes, with concerns |
| Abdou et al., 2018 | Not retracted | No | Yes | No | NA | No |
| Carvalho et al., 2018 | Not retracted | Yes | Yes | No | Yes | Yes, with concerns |
| Ozaki et al., 2020 | Not retracted | Yes | Yes | No | Yes | Yes, with concerns |
| Margatho et al., 2020 | Not retracted | Yes | Yes | Yes | Yes | Yes |

| | | | | | | |
|---|---|---|---|---|---|---|
| Ceccaroni et al., 2021 | Not retracted | No | Yes | No | NA | No |
| El Taha et al., 2021 | Not retracted | Yes | Yes | No | Yes | Yes, with concerns |
| Hassanin et al., 2021 | Not retracted | Yes | Yes | No | Yes | Yes, with concerns |
| Khalifa et al., 2021 | Not retracted | Yes | Yes | Yes | Yes | Yes |
| Osuga et al., 2021a | Not retracted | Yes | Yes | No | Yes | Yes, with concerns |
| Ota et al., 2021 | Not retracted | No | Yes | No | NA | No |
| Mehdizadeh Kashi et al., 2022 | Not retracted | Yes | Yes | No | Yes | Yes, with concerns |
| Guo et al., 2023 | Not retracted | No | Yes | No | NA | No |
| Vahid Dastjerdi et al., 2023 | Not retracted | No | Yes | No | NA | No |
| Amiya et al., 2024 | Not retracted | No | Yes | No | NA | No |
| Cooper et al., 2024 | Not retracted | Yes | Yes | Yes | Yes | Yes |
| da Costa Porto et al., 2024 | Not retracted | Yes | Yes | No | Yes | Yes, with concerns |
| Choudhury et al., 2024 | Not retracted | Yes | Yes | Yes | Yes | Yes |
| Wei et al., 2024 | Not retracted | No | Yes | No | NA | No |
| De La Hoz et al., 2025 | Not retracted | No | Yes | No | NA | No |
| Gurbuz et al., 2025 | Not retracted | Yes | Yes | No | Yes | Yes, with concerns |
| Tanha et al., 2025 | Not retracted | Yes | Yes | No | Yes | Yes, with concerns |

* Met all applicable absolute trustworthiness criteria. Registration and CONSORT requirements were not considered mandatory for trials initiated before 2010.

NA=not applicable. RCT=randomized controlled trial. CONSORT=CONsolidated Standards of Reporting Trials.

**Table S7 Baseline and post-treatment quality of life scores across hormonal therapies for endometriosis, assessed by EHP-30 questionnaire**

| Study | Intervention | *n* | Summery scale score | | | Pain | | | Control and Powerlessness | | | Emotional Well-Being | | | Social Support | | | Self-image | | |
|---|---|---|---|---|---|---|---|---|---|---|---|---|---|---|---|---|---|---|---|---|
| | | | Baseline | Post-TX | *p value* | Baseline | Post-TX | *p value* | Baseline | Post-TX | *p value* | Baseline | Post-TX | *p value* | Baseline | Post-TX | *p value* | Baseline | Post-TX | *p value* |
| Carr et al., 2014* | GnRH-ant | 84 | NA | NA | NA | 47·0±2·4 | 15·4±2·6 | NA | 56·3±2·3 | 15·8± 3·1 | NA | 59·8±2·3 | 34·6±3·7 | NA | 67·0±2·9 | 21·5±3·6 | NA | 47·9±3·1 | 21·1±3·7 | NA |
| | GnRH-ant | 84 | NA | NA | NA | 50·6±2·2 | 21·5±2·8 | NA | 61·3±2·8 | 26·2± 3·6 | NA | 61·3±2·4 | 37·7±3·0 | NA | 69·9±2·7 | 28·8±3·8 | NA | 52·7±3·3 | 21·9±3·6 | NA |
| | DMPA | 83 | NA | NA | NA | 48·5±2·4 | 22·1±3·5 | NA | 63·0±2·4 | 27·5±4·3 | NA | 61·4±2·3 | 36·8±3·9 | NA | 68·4±2·8 | 34·8±4·8 | NA | 48·8±3·0 | 24·0±4·1 | NA |
| Granese et al., 2015† | continuous COC | 39 | 4·2 ± 1·9 | 8·6 ± 2·0 | NA | NA | NA | NA | NA | NA | NA | NA | NA | NA | NA | NA | NA | NA | NA | NA |
| | GnRH-a | 39 | 4·9 ± 2·0 | 9·1 ± 1·8 | NA | NA | NA | NA | NA | NA | NA | NA | NA | NA | NA | NA | NA | NA | NA | NA |
| Carvalho et al., 2018‡ | ENG | 52 | NA | NA | NA | 68·7±13·2 | 38·2±19·3 | <0·01 | 74·7±16·3 | 42·2±22·1 | <0·01 | 71·6±18·1 | 46·9±20·0 | <0·01 | 64·5±23·9 | 42·9±24·5 | <0·01 | 62·8±24·4 | 41·9±23·2 | <0·01 |
| | LNG-IUD | 51 | NA | NA | NA | 65·2±21·7 | 37·5±17·7 | <0·01 | 66·9±23·6 | 35·9±19·1 | <0·01 | 58·1±21·3 | 43·1±17·2 | <0·01 | 55·1±24·6 | 44·5±22·2 | <0·05 | 50·3±27·5 | 41·0±26·2 | <0·05 |
| El Taha et al., 2021‡ | DNG | 35 | NA | NA | NA | 69·7±22·8 | 19·6±18·0 | <0·01 | 69·0±27·7 | 18·5±19·8 | <0·01 | 55·7±27·2 | 32·1±30·6 | <0·01 | 48·5±31·8 | 22·7±26·1 | <0·01 | 40·7±31·0 | 17·9±18·1 | <0·01 |
| | continuous COC | 35 | NA | NA | NA | 61·6±22·5 | 29·0±30·0 | <0·01 | 74·0±29·7 | 36·2±31·4 | <0·01 | 66·3±31·4 | 52·2±35·1 | ns | 65·6±29·9 | 48·8±36·2 | <0·05 | 53·2±31·4 | 31·7±31·3 | <0·01 |
| Osuga et al., 2021a | GnRH-ant | 103 | NA | NA | NA | 28·9±20·1 | [-25·9 (19·90)] | NA | 25·9±21·2 | [-20·9 (21·68)] | NA | 20·4±17·5 | [-13·3 (16·32)] | NA | 15·7±18·7 | [-10·3 (17·11)] | NA | 15·0±18·7 | [-9·7 (17·74)] | NA |
| | GnRH-a | 81 | NA | NA | NA | 26·5±19·6 | [-26·4 (20·34)] | NA | 27·8±22·9 | [-24·8 (23·84)] | NA | 21·2±19·1 | [-12·4 (18·33)] | NA | 17·1±20·3 | [-10·5 (17·92)] | NA | 16·3±21·9 | [-9·4 (15·55)] | NA |
| Cooper et al., 2024‡ | IUD-LNG/ DMPA | 197 | NA | NA | NA | 56·6±17·3 | 32·9±25·0 | NA | 69·1±19·7 | 40·9±28·5 | NA | 53·0±20·3 | 35·6±26·6 | NA | 56·8±23·5 | 40·7±31·5 | NA | 54·3±28·4 | 43·7±34·4 | NA |
| | continuous/cyclic COC | 192 | NA | NA | NA | 55·8±19·9 | 32·9±27·6 | NA | 66·6±23·4 | 45·4±34·2 | NA | 52·4±23·2 | 38·6±31·1 | NA | 56·5 (26·5) | 48·4±36·1 | NA | 52·6±29·0 | 48·1±36·7 | NA |
| da Costa Porto et al., 2024‡ | DNG | 20 | 67·3±23·1 | 29·6±22·6 | <0·01 | NA | NA | NA | NA | NA | NA | NA | NA | NA | NA | NA | NA | NA | NA | NA |
| | LNG-IUD | 20 | 75·7±20·1 | 25·7±24·3 | <0·01 | NA | NA | NA | NA | NA | NA | NA | NA | NA | NA | NA | NA | NA | NA | NA |

All quality-of-life scores reported as mean ± SD, as provided in the original studies. Post-treatment values can also be expressed as a reduction from baseline (Osuga et al., 2021a). P values are reported as provided in the original studies; 'ns' indicates non-significant results.

*Questionnaire EHP-5: 5 core items and 6 modular items (0-4), final score converted in 0-100.
†Questionnaire EHP-5 with 0-10 score.
‡Only the results for the main domains are shown here, please refer to the original article for the results of the modular questionnaires.

NA=not applicable. EHP=endometriosis health profile. TX=treatment. COC=combined oral contraceptive. DNG=dienogest. LNG-IUD=levonorgestrel-intrauterine device. GnRH-a=gonadotropin resealing hormone agonist. GnRH-ant=gonadotropin resealing hormone antagonist. DMPA=depot medroxyprogesterone acetate. ENG=etonogestrel.

**Table S8 Baseline and post-treatment quality of life scores across hormonal therapies for endometriosis, assessed by SF-36 questionnaire**

| Study | Intervention | *n* | Physical functioning | | Role limitation (physical) | | Pain | | General health | | Vitality | | Social functioning | | Role limitation (emotional) | | Mental health | |
|---|---|---|---|---|---|---|---|---|---|---|---|---|---|---|---|---|---|---|
| | | | Baseline | Post-TX | Baseline | Post-TX | Baseline | Post-TX | Baseline | Post-TX | Baseline | Post-TX | Baseline | Post-TX | Baseline | Post-TX | Baseline | Post-TX |
| Vercellini et al., 2002 | continuous COC | 45 | 79·9 ± 20·6 | 85·1 ± 14·2* | 42·8 ± 40·9 | 79·2 ± 25·8 | 46·6 ± 20·7 | 69·8 ± 20·9* | 55·0 ± 23·5 | 60·6 ± 13·1 | 47·5 ± 18·1 | 52·3 ± 17·5 | 56·4 ± 24·3 | 67·3 ± 25·4 | 41·9 ± 40·7 | 81·9 ± 27·2* | 52·7 ± 19·5 | 61·3 ± 13·9* |
| | CPA | 45 | 81·3 ± 19·9 | 93·6 ± 10·1* | 50·4 ± 35·9 | 89·5 ± 21·4* | 44·7 ± 22·2 | 81·3 ± 19·4* | 52·5 ± 18·6 | 65·8 ± 15·6* | 49·2 ± 15·0 | 63·3 ± 14·6* | 58·9 ± 20·5 | 77·0 ± 19·8* | 58·8 ± 38·3 | 80·6 ± 34·1* | 55·7 ± 15·6 | 66·1 ± 14·7* |
| Zupi et al., 2004 | GnRH-a + add back | 46 | 52·6 ± 14·4 | 66·4 ± 15·1* | 58·3 ± 13·0 | 57·3 ± 14·8 | 47·1 ± 19·2 | 63·6 ± 17·0* | 47·9 ± 12·7 | 59·6 ± 13·7* | 52·7 ± 11·0 | 68·0± 12·8* | 56·4 ± 11·0 | 58·3 ± 12·7 | 60·8 ± 12·0 | 60·0 ± 14·4 | 58·1 ± 12·3 | 60·5 ± 14·8 |
| | GnRH-a | 44 | 51·6 ± 13·2 | 57·6 ± 14 | 59·2 ± 15·4 | 60·1 ± 13·9 | 46·4 ± 18·5 | 62·1 ± 14·0* | 49·4 ± 14·2 | 54·9 ± 12·7 | 53·4 ± 10·3 | 57·8 ± 11·3 | 55·6 ± 9·7 | 54·5 ± 11·5 | 60·5 ± 11·9 | 62·3 ± 15·2 | 59·8 ± 12·9 | 60·2 ± 13·6 |
| | continuous COC | 43 | 52·8 ± 10·9 | 55·6 ± 13·2 | 57·1 ± 13·9 | 58·8 ± 12·0 | 50·1 ± 14·0 | 58·3 ± 14·2 | 48·1 ± 12·1 | 51·2 ± 14·2 | 52·3 ± 11·3 | 56·1 ± 19·2 | 58·5 ± 11·5 | 56·7 ± 11·0 | 60·1 ± 15·2 | 58·1 ± 12·3 | 60·2 ± 13·6 | 59·4 ± 11·0 |
| Strowitzki et al., 2010 | DNG | 90 | 41·4 ± 8·5† | 51·6 ± 6·7† | NA | NA | NA | NA | NA | NA | NA | NA | NA | NA | NA | NA | 42·1 ± 11·5‡ | 45·4 ± 10·9‡ |
| | GnRH-a | 96 | 44·2 ± 8·0† | 51·2 ± 7·1† | NA | NA | NA | NA | NA | NA | NA | NA | NA | NA | NA | NA | 44·0 ± 11·6‡ | 45·9 ± 11·7‡ |
| da Costa Porto et al., 2024 | DNG | 20 | 37·0 ± 26·3 | 80·9 ± 23·5* | 13·8 ± 28·6 | 82·5 ± 36·4* | 20·0 ± 19·1 | 58·4 ± 19·3* | 18·9 ± 19·7 | 55·5 ± 28·5* | 25·5 ± 20·8 | 63·0 ± 28·2* | 37·5 ± 26·3 | 75·8 ± 23·6* | 12·3 ± 31·8 | 80·7 ± 39·0* | 28·8 ± 23·3 | 67·8 ± 27·8* |
| | LNG-IUD | 20 | 37·1 ± 22·8 | 80·4 ± 25* | 5·3 ± 18·5 | 83·5 ± 36·6* | 21·4 ± 16·3 | 59·7 ± 26·1* | 27·6 ± 24·9 | 57·9 ± 24·2* | 21·3 ± 18·4 | 59·5 ± 30·5* | 30·4 ± 20·7 | 77·5 ± 28·6* | 8·8 ± 26·9 | 84·2 ± 37·5* | 26·0 ± 23·3 | 63·8 ± 30·9* |

All quality-of-life scores reported as mean ± SD.

*Statistically significant reduction (*p* value<0·05).
†Physical health summary scale score
‡Mental health summary scale score

NA=not applicable. SF-36=36-item short form survey. TX=treatment. COC=combined oral contraceptives. CPA=cyproterone acetate. DNG=dienogest. LNG-IUD=levonorgestrel-intrauterine device. GnRH-a=gonadotropin resealing hormone agonist. SD=standard deviation.

**Table S9 Baseline and post-treatment quality of life scores across hormonal therapies for endometriosis, assessed by WHOQOL-BREF questionnaire**

| Study | Intervention | *n* | Physical health | | | Psychological health | | | Social relationships | | | Environmental health | | |
|---|---|---|---|---|---|---|---|---|---|---|---|---|---|---|
| | | | Baseline | Post-TX | *p* value | Baseline | Post-TX | *p* value | Baseline | Post-TX | *p* value | Baseline | Post-TX | *p* value |
| Mehdizadeh Kashi et al., 2022 | DNG | 30 | 21·56 ± 3·4 | 26·07 ± 4·9 | ns | 17·52 ± 2·24 | 23·19 ± 3·74 | ns | 8·93 ± 1·99 | 10·89 ± 4·3 | ns | 21·44 ± 3·76 | 31·3 ± 11·26 | 0·03 |
| | continuous COC | 30 | 20·56 ± 3·01 | 26·59 ± 3·73 | ns | 18·53 ± 2·04 | 23·48 ± 5·13 | ns | 8·9 ± 1·79 | 11·15 ± 6·55 | ns | 21·93 ± 2·97 | 31·78 ± 13·13 | ns |
| Choudhury et al., 2024 | LNG-IUD | 34 | 59·97 ± 16·12 | 67·09 ± 17·78 | <0·001 | 55·79 ± 12·67 | 66·06 ± 11·60 | <0·001 | 56·29 ± 20·57 | 62·68 ± 21·29 | <0·001 | 58·94 ± 14·49 | 63·94 ± 12·02 | <0·001 |
| | DNG | 34 | 54·35 ± 19·67 | 67·94 ± 13·83 | <0·001 | 59·26 ± 15·67 | 71·15 ± 15·37 | <0·001 | 65·26 ± 19·77 | 75·97 ± 13·63 | <0·001 | 62·09 ± 19·81 | 74·18 ± 16·41 | <0·001 |

All quality-of-life scores are reported as mean ± SD, as provided in the original studies. P values are reported as provided in the original studies; ‘ns’ indicates non-significant results.

NA=not applicable. WHOQOL-BREF=world health organization quality of life-abbreviated version. TX=treatment. COC=combined oral contraceptive. DNG=dienogest. LNG-IUD=levonorgestrel-intrauterine device

**Table S10 Recurrence of lesions and reappearance or worsening of symptoms during and after hormonal therapies for endometriosis**

| Study | TX length (months) | Intervention | *n* | Recurrence of lesions (or progression) | Reappearance or worsening of pain symptoms | Criteria used to define recurrence of lesions or symptom worsening |
|---|---|---|---|---|---|---|
| Vercellini et al., 2002 | 6 | continuous COC | 45 | NA | 2 | Withdraw for treatment inefficacy |
| | | CPA | 45 | NA | 2 | |
| Vercellini et al., 2005 | 12 | continuous COC | 45 | NA | 3 | Withdraw for treatment inefficacy |
| | | NETA | 45 | NA | 3 | |
| Schlaff et al., 2006 | 6 (follow up 18 months) | DMPA | 136 | NA | 7 | Withdraw for perceived lack of efficacy, most after only one dose of study drug. Treatment duration 6 months; follow-up 18 months (12 months off therapy). |
| | | GnRH-a | 138 | NA | 1 | |
| Sesti et al., 2009 | 6 (follow up 18 months) | GnRH-a | 58 | 6 | NA | Recurrence of ovarian endometriomas. Treatment duration 6 months; follow-up 18 months (12 months off therapy). |
| | | continuous COC | 60 | 9 | NA | |
| Walch et al., 2009 | 12 | ENG | 21 | NA | 1 | Withdraw for therapy-resistant pelvic pain |
| | | DMPA | 20 | NA | 1 | |
| Seracchioli et al., 2010a | 24 | cyclic OC | 75 | 11 (14·7%) | NA | Recurrence of ovarian endometriomas |
| | | continuous COC | 73 | 6 (8·2%) | NA | |
| Wong et al., 2010 | 36 | LNG-IUD | 15 | 0 | NA | NA |
| | | DMPA | 15 | 0 | NA | |
| Muzii et al., 2011 | 6 (minimum follow-up of 12 months, mean 22 months) | continuous COC | 29 | 0 | 5 | Recurrence of ovarian endometriomas and recurrence of pain. Treatment duration 6 months, minimum follow-up of 12 months (at least 6 months off therapy) |
| | | COC cyclic | 28 | 1 | 9 | |
| Cheewadhanarak et al., 2012 | 6 | DMPA | 42 | NA | 1 | Withdraw for persistent pain |
| | | continuous COC | 42 | NA | 3 | |
| Cucinella et al., 2013 | 24 | continuous COC (desogestrel) | 43 | 4 | NA | Recurrence of ovarian endometriomas |

| | | | | | | |
|---|---|---|---|---|---|---|
| | | continuous COC (gestodene) | 44 | 2 | NA | |
| | | cyclic COC (DNG) | 43 | 1 | NA | |
| Carr et al., 2014 | 6 | GnRH-ant | 84 | NA | 4 | Withdraw for pain symptoms |
| | | GnRH-ant | 84 | NA | 2 | |
| | | DMPA | 84 | NA | 3 | |
| Shaaban et al., 2015 | 6 | LNG-IUD | 31 | NA | 1 | Patient request removal of IUD because no improvement was observed |
| | | cyclic COC | 31 | NA | 0 | |
| Granese et al., 2015 | 9 for COC, 6 for GnRH-a | continuous COC | 39 | 3 | NA | Recurrence of ovarian endometriomas at 9 mo of follow up for COC and at 6 mo of follow up for GnRH-a. |
| | | GnRH-a | 39 | 1 | NA | |
| Takaesu et al., 2016 | 6 (follow up 24 months) | DNG | 54 | 4 (7·4%) | NA | Recurrence of endometriosis based on MRI. Treatment duration 6 months; follow-up 24 months (18 months off therapy). |
| | | GnRH-a | 51 | 8 (15·7%) | NA | |
| Margatho et al., 2020 | 24 | ENG | 52 | NA | 1 | Withdraw for pain relief not achieved |
| | | LNG-IUD | 51 | NA | 0 | |
| Ceccaroni et al., 2021 | 6 (follow up 30 months) | GnRH-a | 81 | 7·4% | 18·5% | Recurrence of ovarian endometriomas or DIE and symptoms relapse at 30 mo. Treatment duration 6 months; median follow-up 30±6 months (at least 18 months off therapy). |
| | | DNG | 65 | 10·7% | 29·2% | |
| El Taha et al., 2021 | 6 | DNG | 35 | NA | 2 | Withdraw for pain relief not achieved |
| | | continuous COC | 35 | NA | 2 | |
| Osuga et al., 2021a | 6 | GnRH-ant | 103 | NA | 0 | Lack of efficacy |
| | | GnRH-a | 80 | NA | 0 | |
| Ota et al., 2021 | 72 | LNG-IUD | 76 | 9 | NA | Five cases of adenomyosis progression and 4 cases of endometrioma progression |
| | | DNG | 81 | 0 | NA | |
| Mehdizadeh Kashi et al., 2022 | 6 | DNG | 30 | NA | 2 (6·6%) | Pain recurrence |
| | | continuous COC | 30 | NA | 2 (6·6%) | |
| Vahid Dastjerdi et al., 2023 | 6 | DNG | 48 | 7 (14·6%) | NA | Recurrence of ovarian endometriomas after surgery |
| | | oral MPA | 53 | 13 (24·5%) | NA | |
| Amiya et al., 2024 | 3 | DNG | 30 | NA | 2 (6·7%) | |

| | | | | | | |
|---|---|---|---|---|---|---|
| | | oral MPA | 30 | NA | 8 (26·7%) | Persistence of dysmenorrhea during the treatment period |
| Cooper et al., 2024 | 36 | IUD-LNG/DMPA | 205 | NA | 24 | Patients withdrew because needed second-line medical treatment (GnRH-a) |
| | | continuous/cyclic COC | 200 | NA | 25 | |
| Wei et al., 2024 | 12 | ENG | 50 | 8 (16%) | NA | New ovarian cysts (not specified the nature) |
| | | LNG-IUD | 58 | 8 (14·5%) | NA | |
| Gurbuz et al., 2025 | 12 | DNG | 40 | NA | 2 | Withdraw because resistance to treatment |
| | | NETA | 30 | NA | 2 | |

Data are reported as number of patients (N), percentage (%), or both (N, %), as provided in the original studies.

NA=not applicable. COC=combined oral contraceptive. CPA=cyproterone acetate. NETA=norethisterone acetate. DNG=dienogest. LNG-IUD=levonorgestrel-intrauterine device. GnRH-a=gonadotropin resealing hormone agonist. GnRH-ant=gonadotropin resealing hormone antagonist. (D)MPA=(depot) medroxyprogesterone acetate. ENG=etonogestrel. MRI=magnetic resonance imaging.

### Table S11 Treatment satisfaction across hormonal therapies for endometriosis, reported as frequencies according to different satisfaction levels

| Study | Interventions (*n*) | Level of satisfaction | Treatment 1 frequency | Treatment 2 frequency |
|---|---|---|---|---|
| Cosson et al., 2002 | DNG (59)<br>GnRH-a (61) | Very satisfied | 34·4% | 30% |
| | | Satisfied | 51·7% | 50% |
| Vercellini et al., 2002 | continuous COC (45) vs CPA (45) | Very satisfied | 5 (11%) | 6 (13%) |
| | | Satisfied | 25 (56%) | 27 (60%) |
| | | Uncertain | 2 (4%) | 2 (4%) |
| | | Dissatisfied | 13 (29%) | 9 (20%) |
| | | Very dissatisfied | 1 (2%) | 1 (2%) |
| Vercellini et al., 2005 | COC continuous (45) vs NETA (45) | Very satisfied | 6 (13%) | 11 (24%) |
| | | Satisfied | 22 (49%) | 22 (49%) |
| | | Uncertain | 8 (18%) | 8 (18%) |
| | | Dissatisfied | 7 (16%) | 3 (7%) |

| | | | | |
|---|---|---|---|---|
| | | Very dissatisfied | 2 (4%) | 1 (2%) |
| Walch et al., 2009 | ENG (21) vs DMPA (20) | Very satisfied | 24% | 26% |
| | | Satisfied | 33% | 32% |
| Wong et al·, 2010 | LNG-IUD (15) vs DMPA (15) | Satisfied* | 13 (86.7%) | 7 (46·7%) |
| Muzii et al., 2011 | continuous COC (29) vs cyclic COC (28) | Satisfied or very satisfied | 29 (100%) | 28 (100%) |
| Cheewadhanarak et al., 2012 | DMPA (42) vs continuous COC (42) | Satisfied | 39 (92·9%) | 37 (88·1%) |
| Shaaban et al., 2015 | LNG-IUD (31) vs cyclic COC (31) | Satisfied | 25 | 18 |
| De La Hoz et al., 2025 | DRSP (94) vs cyclic COC (91) | Satisfied | 71 (75·53%) | 65 (71·42%) |
| | | Not satisfied | 7 (7·44%) | 11 (12·08%) |

Data are reported as number of patients (N), percentage (%), or both (N, %), as provided in the original studies.

*Reported as compliance and patients' acceptance/desire to continue therapy beyond 3 years.

TX=treatment. COC=combined oral contraceptive. CPA=cyproterone acetate. NETA=norethisterone acetate. DNG=dienogest. LNG-IUD=levonorgestrel-intrauterine device. GnRH-a=gonadotropin resealing hormone agonist. DMPA=depot medroxyprogesterone acetate. ENG=etonogestrel. DRSP=drospirenone.

### Table S12 Analgesic use prior to and during hormonal therapies for endometriosis

| Study | TX length (months) | Intervention | *n* | Use of analgesic | | Comment (analgesic type and outcome measure) |
|---|---|---|---|---|---|---|
| | | | | prior to hormonal treatment | during hormonal treatment | |
| Parazzini et al., 2000 | 12 | cyclic COC | 47 | NA | 15 (31·9%) | NSAIDs<br>*n* (%) |
| | | GnRH-a/cyclic COC | 55 | NA | 16 (29·1%) | |
| Walch et al., 2009 | 12 | ENG | 21 | 85·7% | 41·2% | *n* (%) |
| | | DMPA | 20 | 100% | 38·5% | |
| Cheewadhanarak et al., 2012 | 6 | DMPA | 42 | NA | 8 (19%) | Acetaminophen<br>*n* (%) |
| | | continuous COC | 42 | NA | 12 (28·6%) | |
| Carr et al., 2014 | 6 | GnRH-ant | 84 | 21·4% | 23·8% | Opioids |

| | | | | | | |
|---|---|---|---|---|---|---|
| | | GnRH-ant | 84 | 19·0% | 25·0% | % |
| | | DMPA | 84 | 28·9% | 33·7% | |
| Ozaki et al., 2020 | 4 | DNG | 35 | 3·1 ± 4·0 | 1·0 ± 3·0 | NSAIDs<br>Mean ± SD of users |
| | | GnRH-a | 35 | 3·1 ± 4·3 | 1·4 ± 3·8 | |
| De La Hoz et al., 2025 | 6 | DRSP | 94 | NA | 10 (10·6%) | NSAIDs<br>*n* (%) |
| | | cyclic COC | 91 | NA | 5 (5·5%) | |
| Osuga et al., 2021a | 6 | GnRH-ant | 103 | 12 (14·5) | [-10·0 (14·21)] | NSAIDs<br>% of days with use (mean ± SD), change from baseline |
| | | GnRH-a | 81 | 11·6 (13·8) | [-10·2 (13·10)] | |

Outcome measures are reported as provided in the original studies.

TX=treatment. COC=combined oral contraceptive. ENG=etonogesterel. DNG=dienogest. GnRH-a=gonadotropin resealing hormone agonist. GnRH-ant=gonadotropin resealing hormone antagonist. DMPA=depot medroxyprogesterone acetate. DRSP=drospirenone. NSAIDs=non-steroidal anti-inflammatory drugs. SD=standard deviation. NA=not applicable.

### Figure S1 Mean difference in overall pelvic pain with combined oral contraceptives

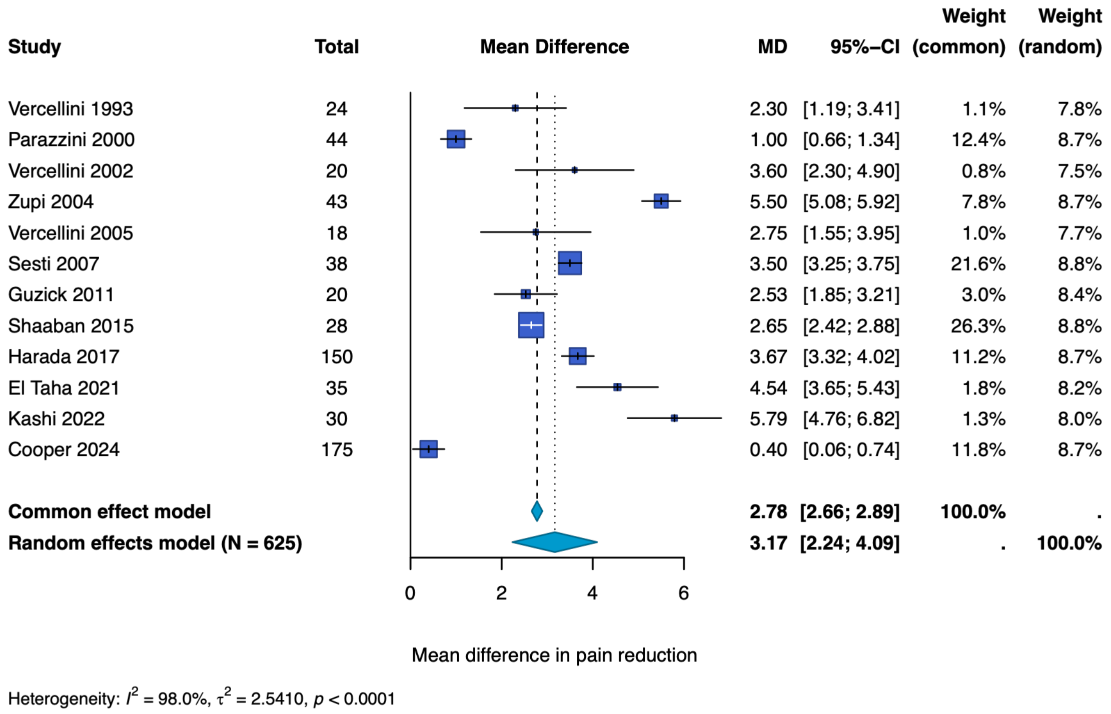

### Figure S2 Mean difference in overall pelvic pain with oral progestogens

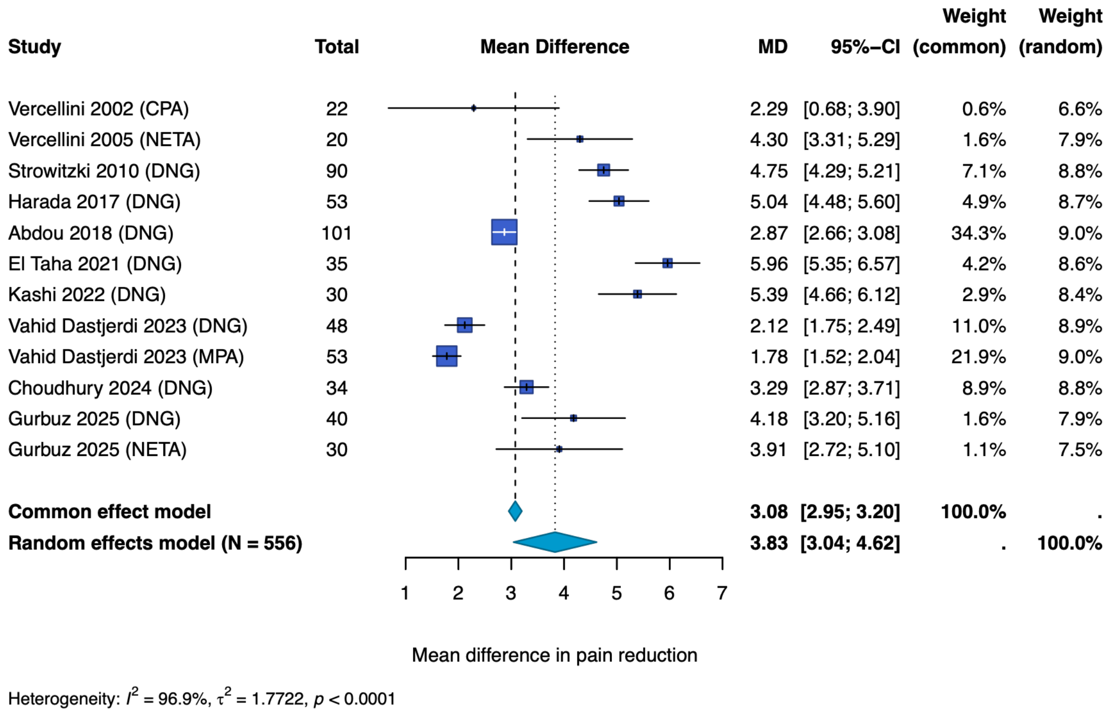

**Figure S3 Mean difference in overall pelvic pain with long-acting progestogens**

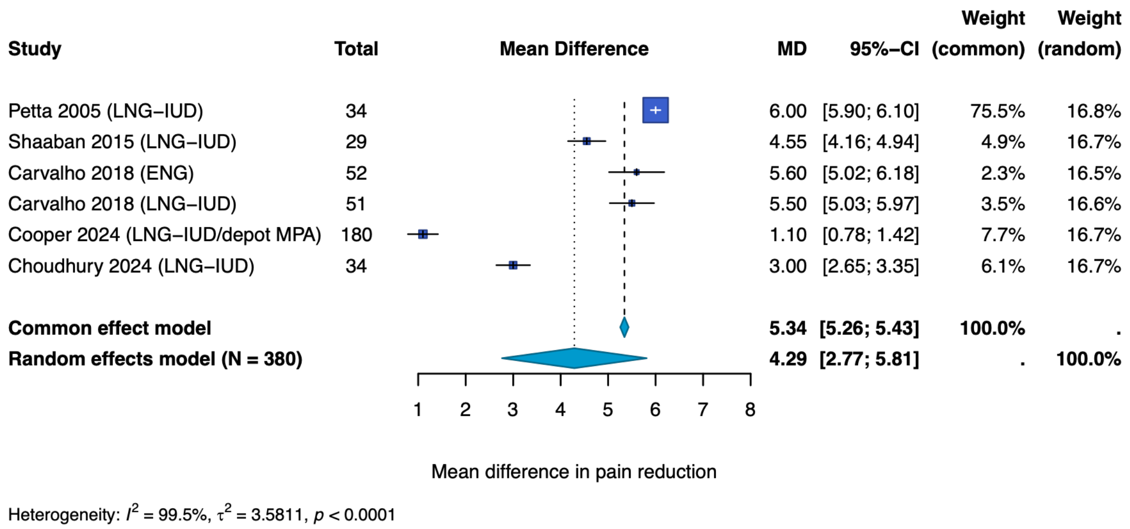

**Figure S4 Mean difference in non-menstrual pelvic pain with GnRH analogues**

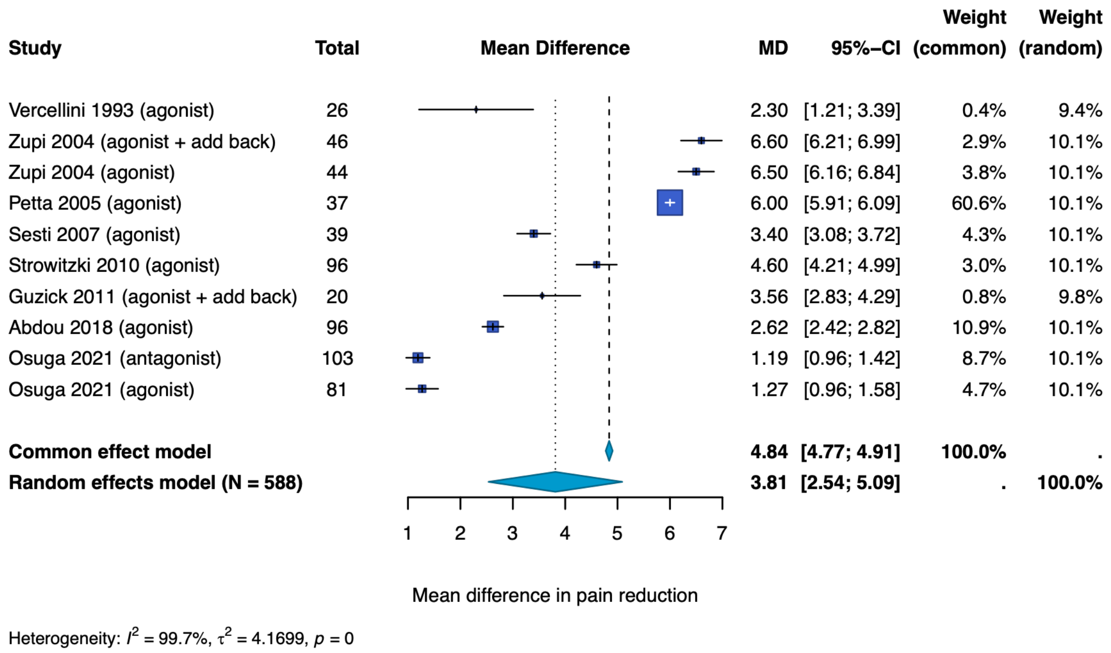

**Figure S5 Pooled proportion of breakthrough bleeding across hormonal treatments**

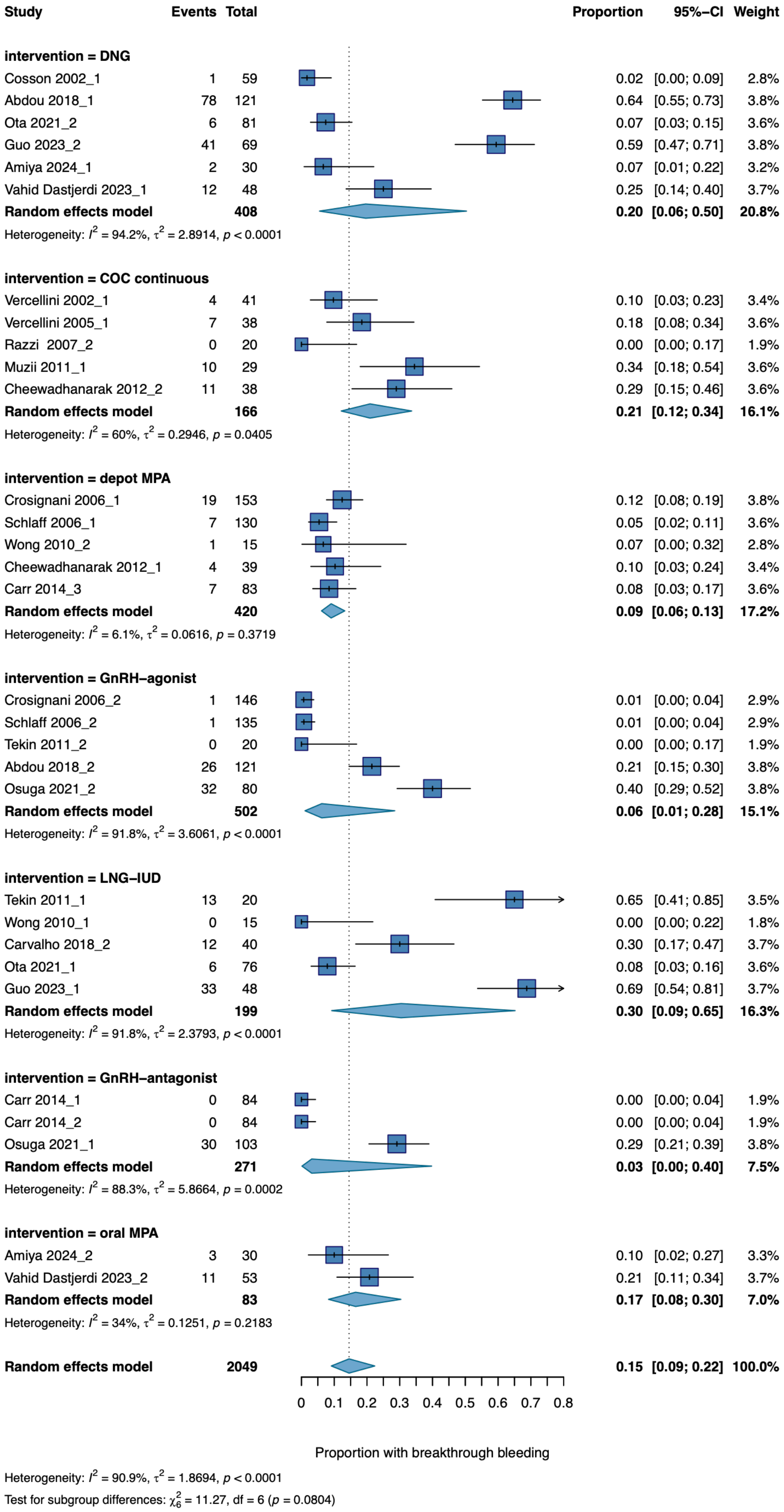

**Figure S6 Pooled proportion of amenorrhoea across hormonal treatments**

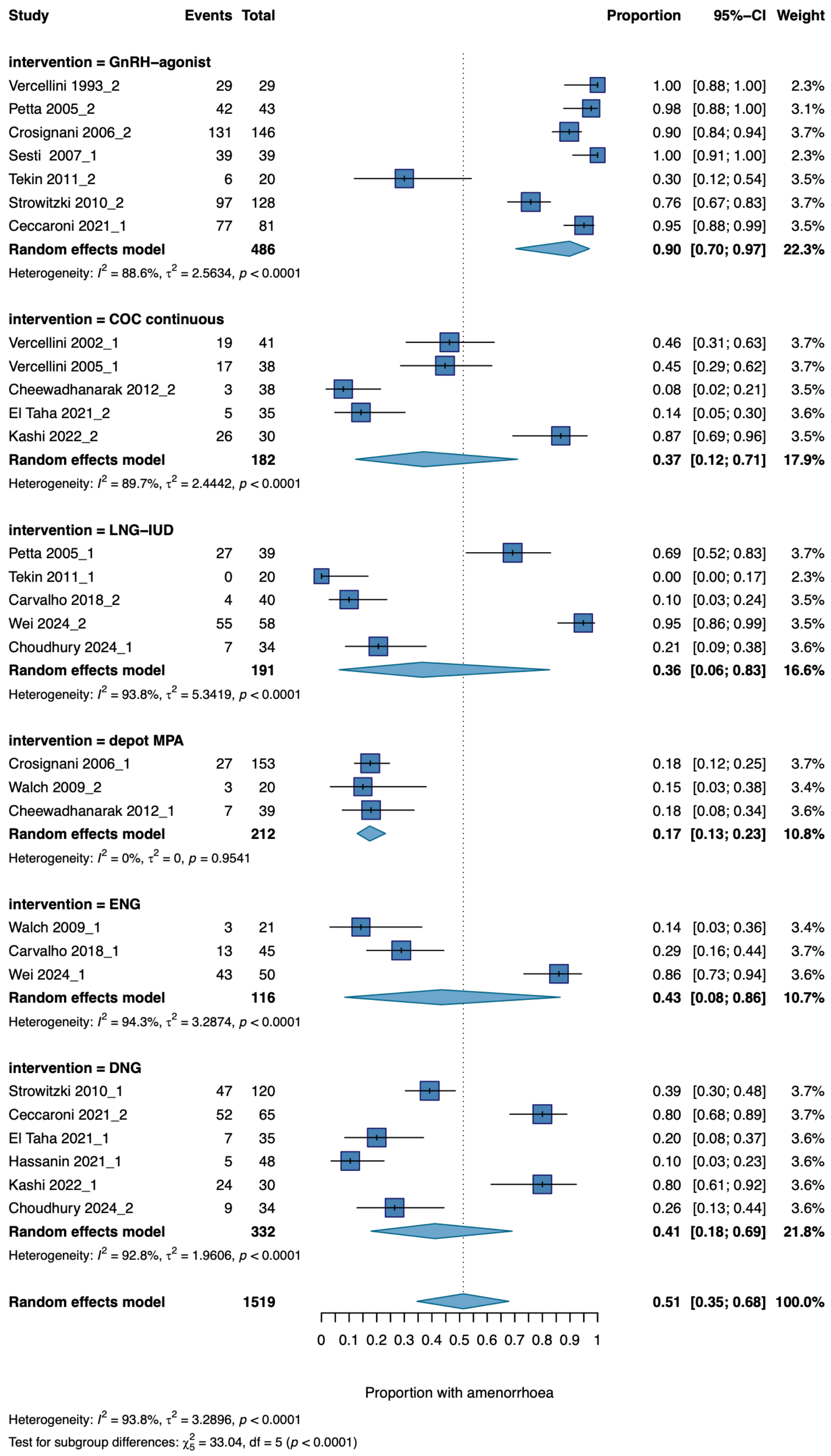

**Figure S7 Pooled proportion of nausea across hormonal treatments**

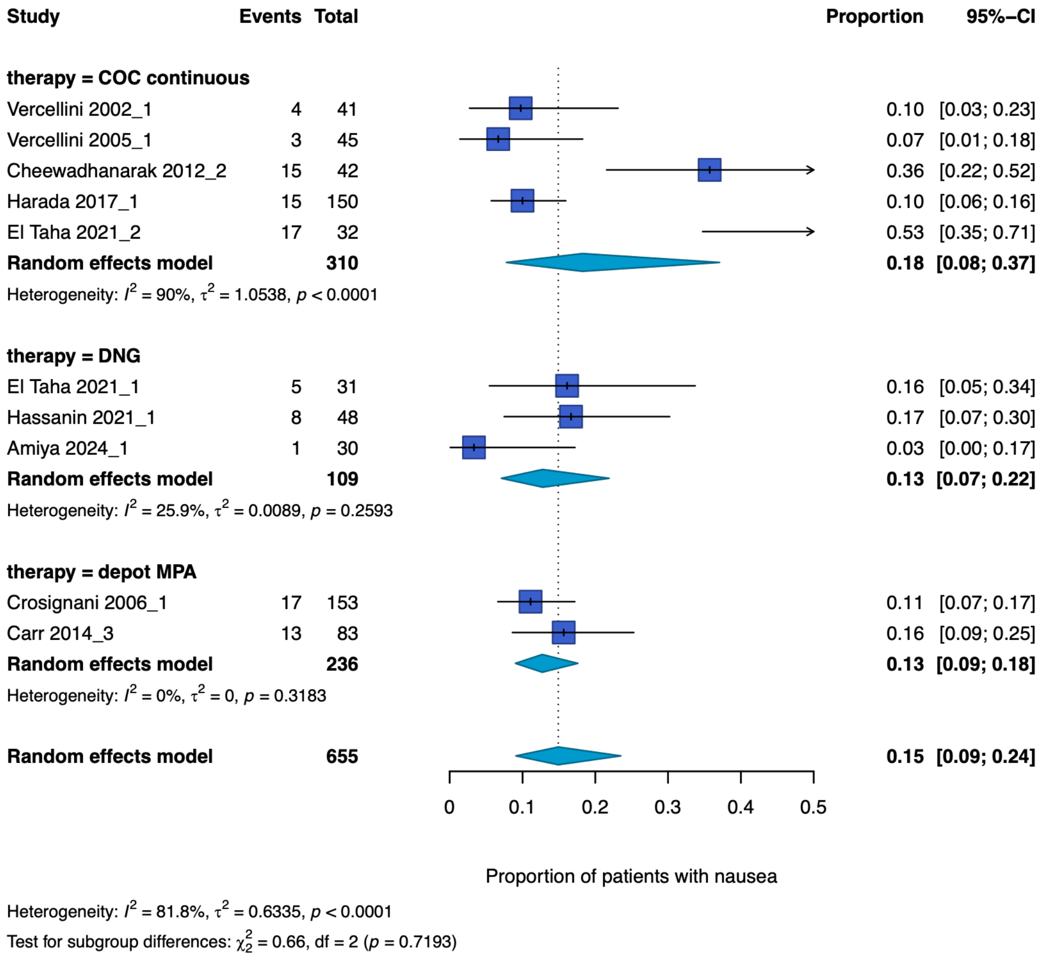

**Figure S8 Pooled proportion of headache across hormonal treatments**

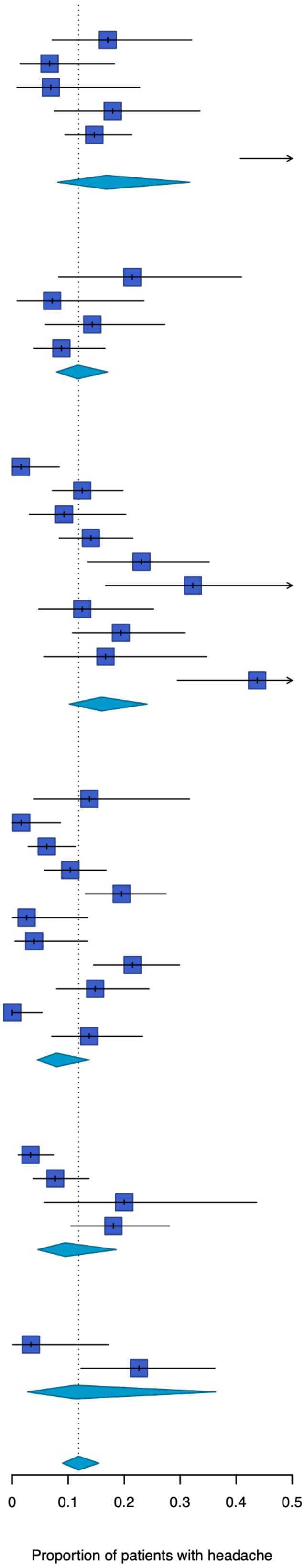

| Study | Events | Total | Proportion | 95%–CI |
|---|---|---|---|---|
| **therapy = COC continuous** | | | | |
| Vercellini 2002_1 | 7 | 41 | 0.17 | [0.07; 0.32] |
| Vercellini 2005_1 | 3 | 45 | 0.07 | [0.01; 0.18] |
| Muzii 2011_1 | 2 | 29 | 0.07 | [0.01; 0.23] |
| Granese 2015_1 | 7 | 39 | 0.18 | [0.08; 0.34] |
| Harada 2017_1 | 22 | 150 | 0.15 | [0.09; 0.21] |
| El Taha 2021_2 | 19 | 32 | 0.59 | [0.41; 0.76] |
| **Random effects model** | | **336** | **0.17** | **[0.08; 0.32]** |
| Heterogeneity: $I^2 = 85.4\%$, $\tau^2 = 0.8671$, $p < 0.0001$ | | | | |
| **therapy = COC cyclic** | | | | |
| Vercellini 1993_1 | 6 | 28 | 0.21 | [0.08; 0.41] |
| Muzii 2011_2 | 2 | 28 | 0.07 | [0.01; 0.24] |
| Hassanin 2021_2 | 7 | 49 | 0.14 | [0.06; 0.27] |
| De La Hoz 2025_2 | 8 | 91 | 0.09 | [0.04; 0.17] |
| **Random effects model** | | **196** | **0.12** | **[0.08; 0.17]** |
| Heterogeneity: $I^2 = 24.1\%$, $\tau^2 = 0$, $p = 0.2664$ | | | | |
| **therapy = DNG** | | | | |
| Cosson 2002_1 | 1 | 64 | 0.02 | [0.00; 0.08] |
| Strowitzki 2010_1 | 15 | 120 | 0.12 | [0.07; 0.20] |
| Takaesu 2016_1 | 5 | 54 | 0.09 | [0.03; 0.20] |
| Abdou 2018_1 | 17 | 121 | 0.14 | [0.08; 0.22] |
| Ceccaroni 2021_2 | 15 | 65 | 0.23 | [0.14; 0.35] |
| El Taha 2021_1 | 10 | 31 | 0.32 | [0.17; 0.51] |
| Hassanin 2021_1 | 6 | 48 | 0.12 | [0.05; 0.25] |
| Khalifa 2021_2 | 13 | 67 | 0.19 | [0.11; 0.31] |
| Amiya 2024_1 | 5 | 30 | 0.17 | [0.06; 0.35] |
| Vahid Dastjerdi 2023_1 | 21 | 48 | 0.44 | [0.29; 0.59] |
| **Random effects model** | | **648** | **0.16** | **[0.10; 0.24]** |
| Heterogeneity: $I^2 = 76.2\%$, $\tau^2 = 0.5339$, $p < 0.0001$ | | | | |
| **therapy = GnRH–agonist** | | | | |
| Vercellini 1993_2 | 4 | 29 | 0.14 | [0.04; 0.32] |
| Cosson 2002_2 | 1 | 62 | 0.02 | [0.00; 0.09] |
| Crosignani 2006_2 | 9 | 146 | 0.06 | [0.03; 0.11] |
| Schlaff 2006_2 | 14 | 135 | 0.10 | [0.06; 0.17] |
| Strowitzki 2010_2 | 25 | 128 | 0.20 | [0.13; 0.27] |
| Granese 2015_2 | 1 | 39 | 0.03 | [0.00; 0.13] |
| Takaesu 2016_2 | 2 | 51 | 0.04 | [0.00; 0.13] |
| Abdou 2018_2 | 26 | 121 | 0.21 | [0.15; 0.30] |
| Ceccaroni 2021_1 | 12 | 81 | 0.15 | [0.08; 0.24] |
| Khalifa 2021_1 | 0 | 67 | 0.00 | [0.00; 0.05] |
| Osuga 2021_2 | 11 | 80 | 0.14 | [0.07; 0.23] |
| **Random effects model** | | **939** | **0.08** | **[0.04; 0.14]** |
| Heterogeneity: $I^2 = 65.8\%$, $\tau^2 = 0.7543$, $p = 0.0011$ | | | | |
| **therapy = depot MPA** | | | | |
| Crosignani 2006_1 | 5 | 153 | 0.03 | [0.01; 0.07] |
| Schlaff 2006_1 | 10 | 130 | 0.08 | [0.04; 0.14] |
| Walch 2009_2 | 4 | 20 | 0.20 | [0.06; 0.44] |
| Carr 2014_3 | 15 | 83 | 0.18 | [0.10; 0.28] |
| **Random effects model** | | **386** | **0.09** | **[0.05; 0.19]** |
| Heterogeneity: $I^2 = 80.4\%$, $\tau^2 = 0.4650$, $p = 0.0016$ | | | | |
| **therapy = oral MPA** | | | | |
| Amiya 2024_2 | 1 | 30 | 0.03 | [0.00; 0.17] |
| Vahid Dastjerdi 2023_2 | 12 | 53 | 0.23 | [0.12; 0.36] |
| **Random effects model** | | **83** | **0.11** | **[0.03; 0.36]** |
| Heterogeneity: $I^2 = 75\%$, $\tau^2 = 0.6980$, $p = 0.0454$ | | | | |
| **Random effects model** | | **2588** | **0.12** | **[0.09; 0.16]** |

Heterogeneity: $I^2 = 74.5\%$, $\tau^2 = 0.7024$, $p < 0.0001$

Test for subgroup differences: $\chi^2_5 = 5.05$, df = 5 ($p = 0.4103$)

**Figure S9 Pooled proportion of weight gain across hormonal treatments**

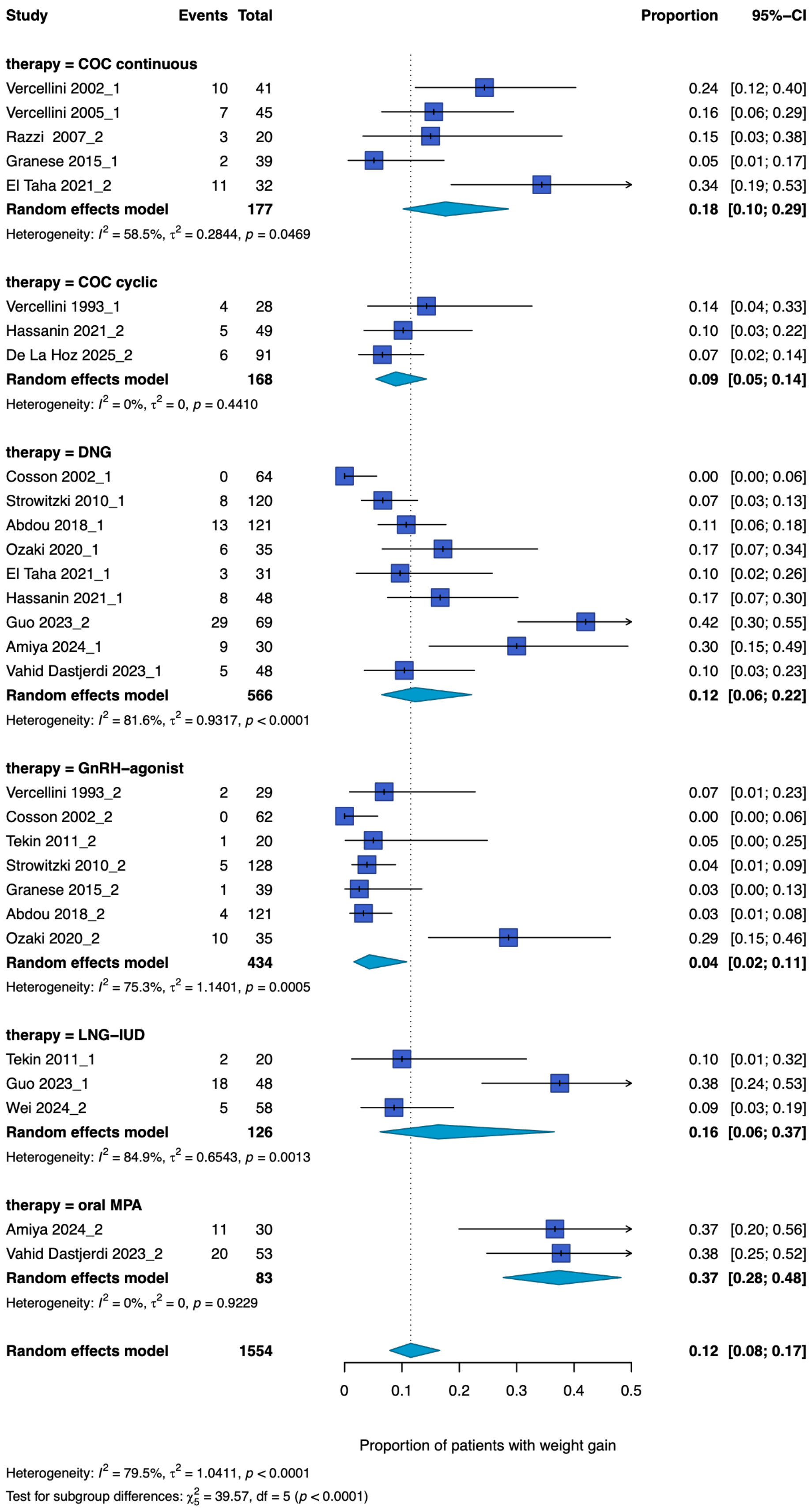

**Figure S10 Pooled proportion of decreased libido across hormonal treatments**

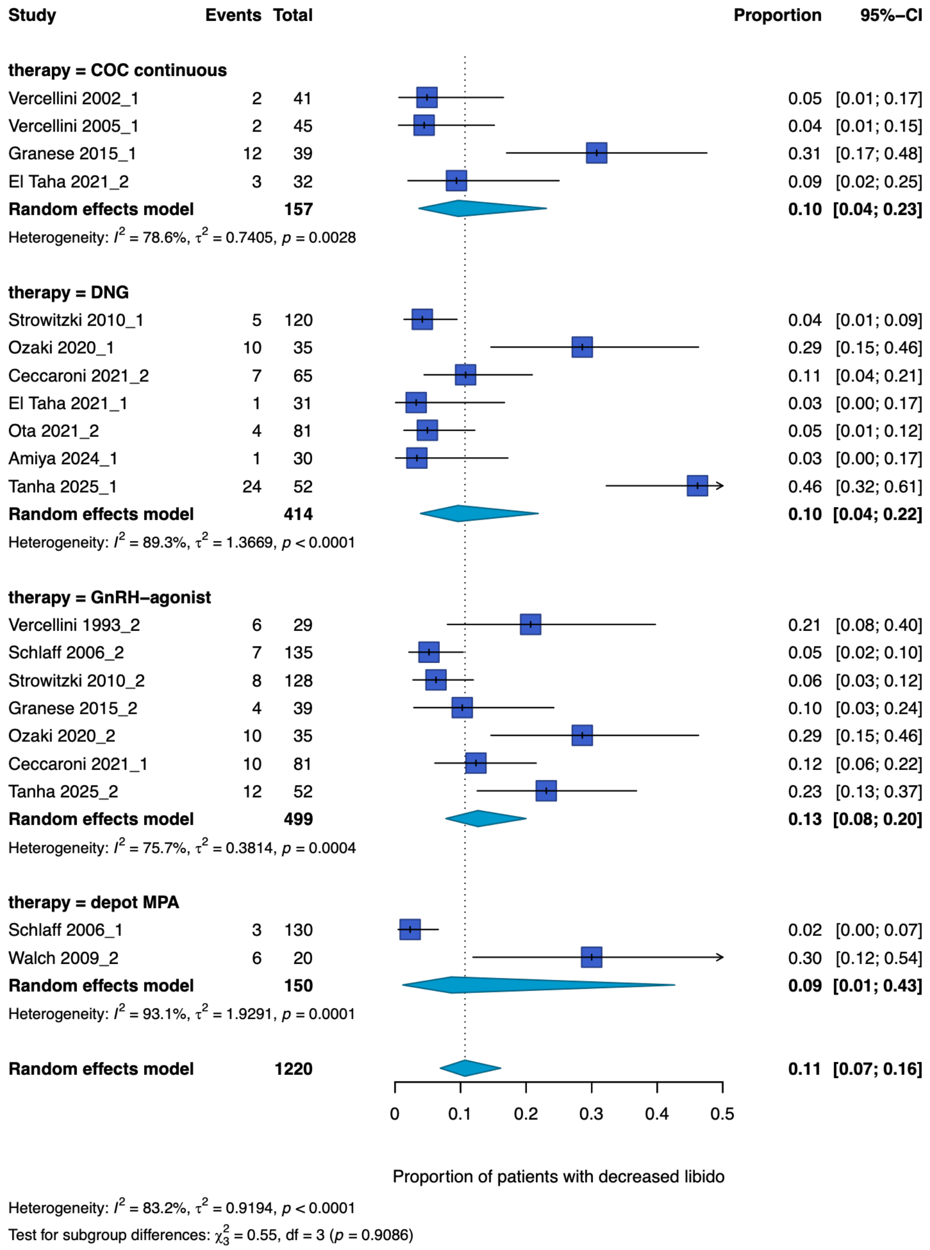

**Figure S11 Pooled proportion of hot flushes/night sweats across hormonal treatments**

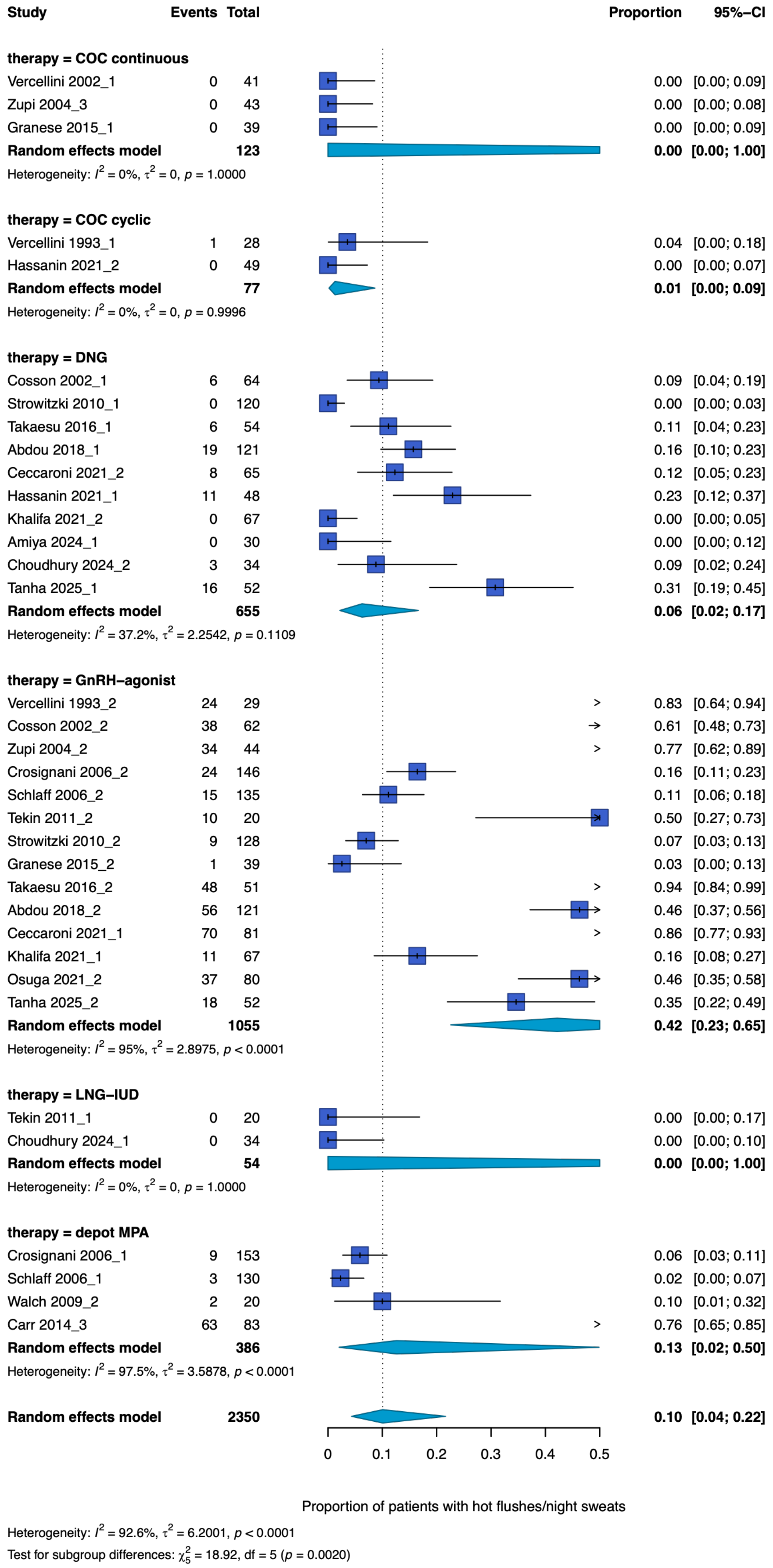

**Figure S12 Pooled proportion of mood breast tenderness across hormonal treatments**

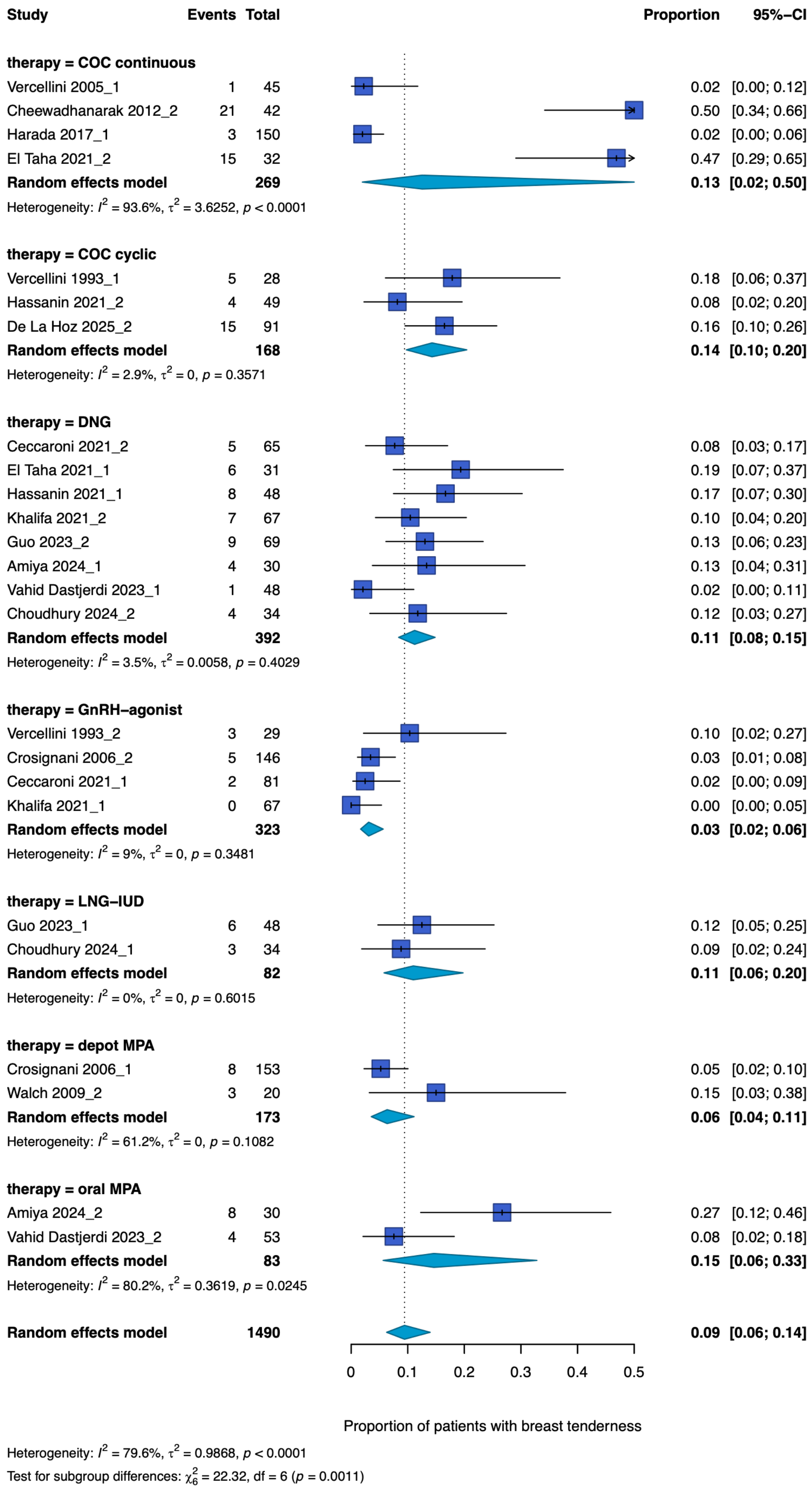

**Figure S13 Pooled proportion of abdominal discomfort/bloating across hormonal treatments**

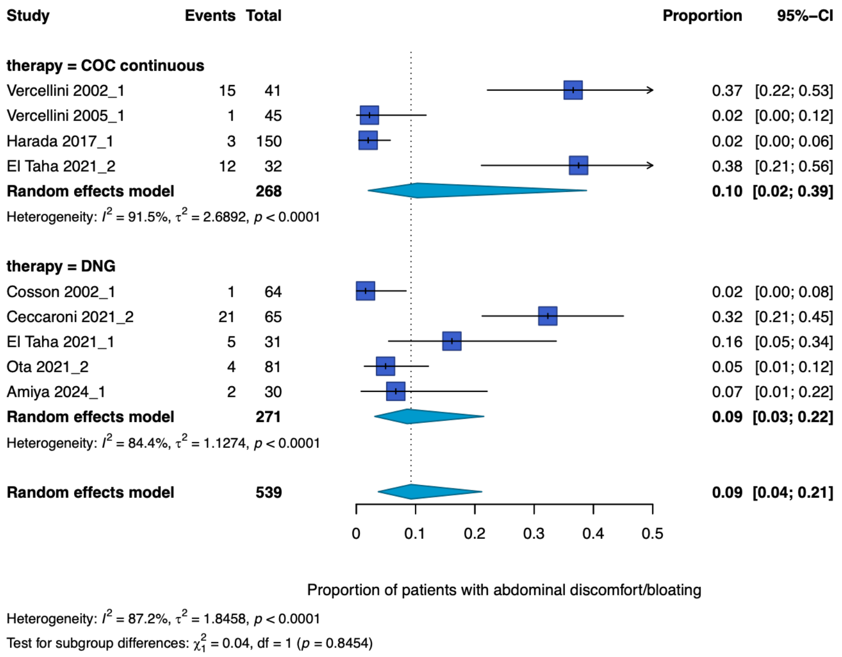

**Figure S14 Pooled proportion of vaginal dryness across hormonal treatments**

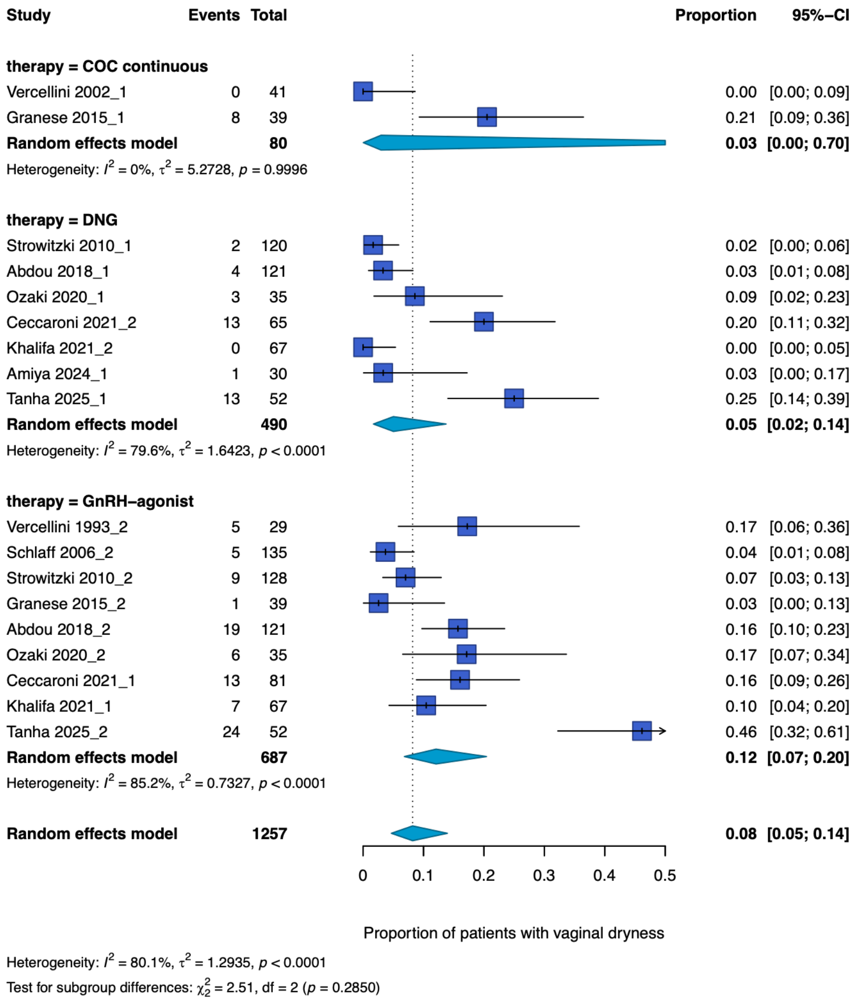

### Figure S15 Pooled proportion of acne/oily skin across hormonal treatments

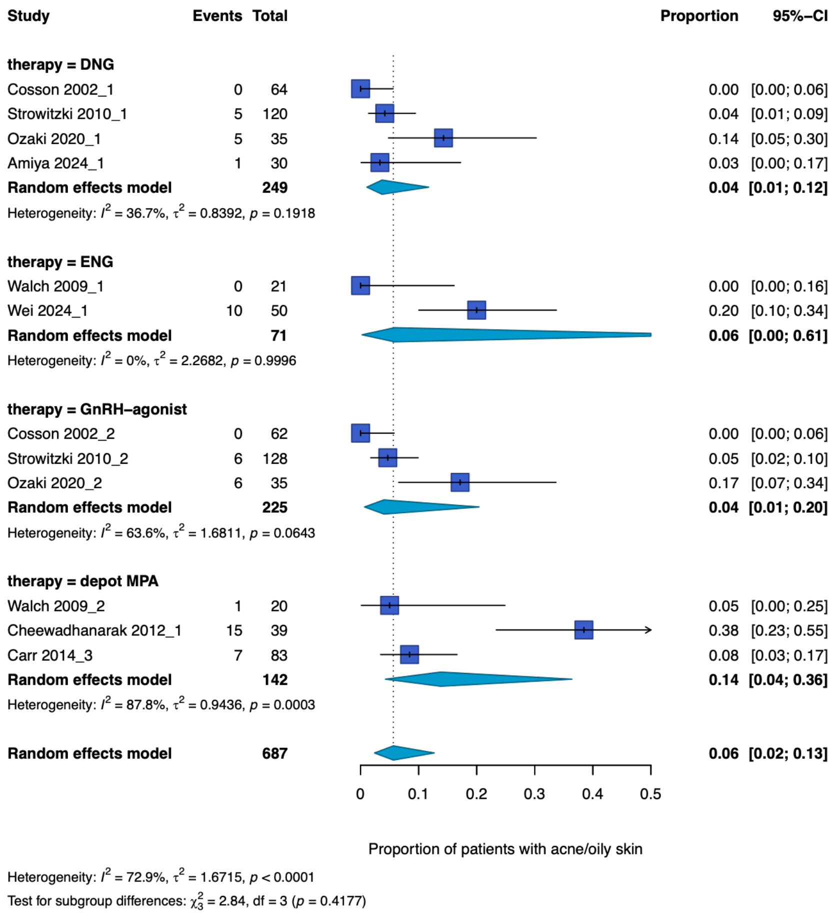

### Figure S16 Pooled proportion of insomnia hormonal treatments

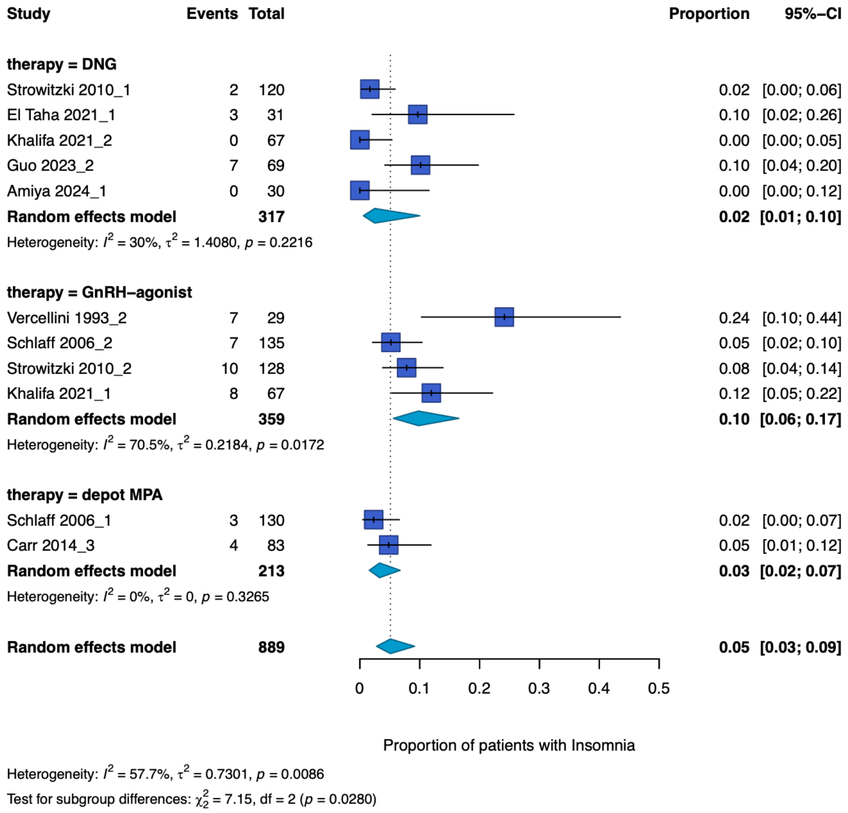

### Figure S17 Pooled proportion of hair loss hormonal treatments

| Study | Events | Total | Proportion | 95%–CI |
|---|---|---|---|---|
| **therapy = DNG** | | | | |
| Cosson 2002_1 | 0 | 64 | 0.00 | [0.00; 0.06] |
| Strowitzki 2010_1 | 4 | 120 | 0.03 | [0.01; 0.08] |
| Takaesu 2016_1 | 0 | 54 | 0.00 | [0.00; 0.07] |
| Ozaki 2020_1 | 5 | 35 | 0.14 | [0.05; 0.30] |
| Ceccaroni 2021_2 | 10 | 65 | 0.15 | [0.08; 0.26] |
| Vahid Dastjerdi 2023_1 | 8 | 48 | 0.17 | [0.07; 0.30] |
| **Random effects model** | | **386** | **0.04** | **[0.01; 0.16]** |
| Heterogeneity: $I^2 = 45.3\%$, $\tau^2 = 2.0287$, $p = 0.1033$ | | | | |
| **therapy = GnRH–agonist** | | | | |
| Cosson 2002_2 | 0 | 62 | 0.00 | [0.00; 0.06] |
| Strowitzki 2010_2 | 7 | 128 | 0.05 | [0.02; 0.11] |
| Takaesu 2016_2 | 1 | 51 | 0.02 | [0.00; 0.10] |
| Ozaki 2020_2 | 7 | 35 | 0.20 | [0.08; 0.37] |
| Ceccaroni 2021_1 | 17 | 81 | 0.21 | [0.13; 0.31] |
| **Random effects model** | | **357** | **0.06** | **[0.01; 0.19]** |
| Heterogeneity: $I^2 = 74.4\%$, $\tau^2 = 1.7733$, $p = 0.0035$ | | | | |
| **Random effects model** | | **743** | **0.05** | **[0.02; 0.12]** |

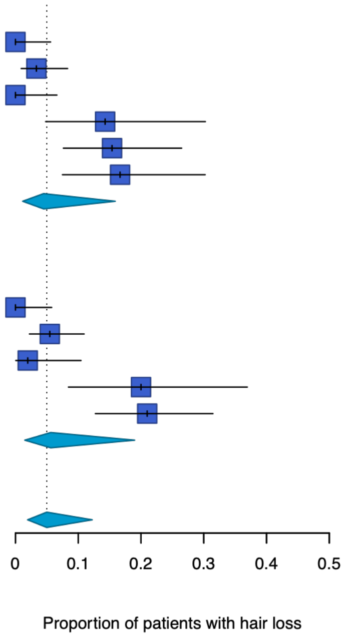

Heterogeneity: $I^2 = 60.2\%$, $\tau^2 = 1.9046$, $p = 0.0052$

Test for subgroup differences: $\chi^2_1 = 0.06$, df = 1 ($p = 0.8116$)

**Figure S18 Risk of bias assessment using the RoB 2 tool**

| Study ID | D1 | D2 | D3 | D4 | D5 | Overall |
|---|---|---|---|---|---|---|
| Vercellini et al., 1993 | + | + | + | − | + | − |
| Parazzini et al., 2000 | + | ? | + | ? | ? | ? |
| Cosson et al., 2002 | ? | + | + | + | + | ? |
| Vercellini et al., 2002 | + | + | + | − | + | − |
| Zupi et al., 2004 | + | + | + | + | + | + |
| Petta et al., 2005 | + | + | + | + | + | + |
| Vercellini et al., 2005 | + | + | + | − | + | − |
| Crosignani et al., 2006 | ? | − | + | + | + | − |
| Schlaff et al., 2006 | + | + | + | + | − | − |
| Razzi et al., 2007 | + | + | + | + | + | + |
| Sesti et al., 2007 | + | + | + | + | + | + |
| Sesti et al., 2009 | + | + | + | + | + | + |
| Walch et al., 2009 | + | + | + | − | + | − |
| Seracchioli et al., 2010a | + | + | + | + | + | + |
| Seracchioli et al., 2010b | + | + | + | + | + | + |
| Strowitzki et al., 2010 | + | + | + | − | + | − |
| Wong et al., 2010 | ? | − | + | + | + | − |
| Muzii et al., 2011 | + | ? | + | + | ? | ? |
| Guzick et al., 2011 | ? | + | + | + | ? | ? |
| Tekin et al., 2011 | + | + | + | − | + | − |
| Cheewadhanarak et al., 2012 | ? | + | + | − | ? | − |
| Cucinella et al., 2013 | − | ? | ? | + | ? | − |
| Carr et al., 2014 | + | − | + | + | + | − |
| Granese et al., 2015 | ? | ? | ? | − | ? | − |
| Shaaban et al., 2015 | + | ? | + | − | + | − |
| Takaesu et al., 2016 | ? | + | + | − | + | − |
| Harada et al., 2017 | ? | + | + | + | + | ? |
| Abdou et al., 2018 | + | + | + | − | + | − |
| Carvalho et al., 2018 | + | + | + | − | + | − |
| Ozaki et al., 2020 | ? | + | + | + | ? | ? |
| Margatho et al., 2020 | + | + | + | − | + | − |
| Ceccaroni et al., 2021 | − | + | + | + | ? | − |
| El Taha et al., 2021 | + | + | + | + | + | + |
| Hassanin et al., 2021 | + | ? | + | ? | ? | ? |
| Khalifa et al., 2021 | + | ? | + | + | + | + |
| Osuga et al., 2021 | + | + | + | + | + | + |
| Ota et al., 2021 | + | ? | + | ? | + | ? |
| Kashi et al., 2022 | + | + | + | + | + | + |
| Guo et al., 2023 | − | ? | ? | ? | ? | − |
| Vahid Dastjerdi et al., 2023 | ? | + | + | + | + | ? |
| Amiya et al., 2024 | ? | − | + | − | ? | − |
| Cooper et al., 2024 | + | ? | + | ? | + | ? |
| da Costa Porto et al., 2024 | + | + | + | + | + | + |
| Choudhury et al., 2024 | ? | ? | + | ? | + | ? |
| Wei et al., 2024 | + | ? | + | + | + | ? |
| De La Hoz et al., 2025 | ? | ? | + | ? | ? | ? |
| Gurbuz et al., 2025 | ? | ? | − | − | + | − |
| Tanha et al., 2025 | ? | ? | + | ? | ? | ? |

Green (+) indicates low risk of bias, yellow (?) indicates some concerns, and red (−) indicates high risk of bias. D1: bias arising from the randomisation process; D2: bias due to deviations from intended interventions; D3: bias due to missing outcome data; D4: bias in measurement of the outcome; D5: bias in selection of the reported result.

**References**

1. Becker CM, Bokor A, Heikinheimo O, Horne A, Jansen F, Kiesel L, King K, Kvaskoff M, Nap A, Petersen K, Saridogan E, Tomassetti C, van Hanegem N, Vulliemoz N, Vermeulen N; ESHRE Endometriosis Guideline Group. ESHRE guideline: endometriosis. Hum Reprod Open. 2022;2022:hoac009.
2. Wan X, Wang W, Liu J, Tong T. Estimating the sample mean and standard deviation from the sample size, median, range and/or interquartile range. BMC Med Res Methodol. 2014;14:135.
3. Luo D, Wan X, Liu J, Tong T. Optimally estimating the sample mean from the sample size, median, mid-range, and/or mid-quartile range. Stat Methods Med Res. 2018;27:1785-1805.
